# The Impact of Mask-Wearing in Mitigating the Spread of COVID-19 During the Early Phases of the Pandemic

**DOI:** 10.1101/2020.09.11.20192971

**Authors:** Ashwin Aravindakshan, Jörn Boehnke, Ehsan Gholami, Ashutosh Nayak

**Author notes:** Correspondence to Ashwin Aravindakshan,.

## Abstract

Masks have been widely recommended as a precaution against COVID-19 transmission. Several studies have shown the efficacy of masks at reducing droplet dispersion in lab settings. However, during the early phases of the pandemic, the usage of masks varied widely across countries. Using individual response data from the Imperial College London — YouGov personal measures survey, this study investigates the effect of mask use within a country on the spread of COVID-19. The survey shows that mask-wearing exhibits substantial variations across countries and over time during the pandemic’s early phase. We use a reduced form econometric model to relate population-wide variation in mask-wearing to the growth rate of confirmed COVID-19 cases. The results indicate that mask-wearing plays an important role in mitigating the spread of COVID-19. Widespread mask-wearing within a country associates with an expected 7% (95% CI: 3.94% — 9.99%) decline in the growth rate of daily active cases of COVID-19 in the country. This daily decline equates to an expected 88.5% drop in daily active cases over a 30-day period when compared to zero percent mask-wearing, all else held equal. The decline in daily growth rate due to the combined effect of mask-wearing, reduced outdoor mobility, and non-pharmaceutical interventions averages 28.1% (95% CI: 24.2%-32%).

## 1. Introduction

In response to the COVID-19 pandemic, multiple countries curbed the spread of the disease by enforcing strict policy measures such as lockdowns and shelter-in-place orders [1]. The non-pharmaceutical interventions (NPIs) included closures of schools, restaurants, bars, retail outlets, and other non-essential businesses, as well as shelter-in-place policies and the prohibition of large gatherings (e.g., limited to 10 people) [2]. These institutional measures aimed to reduce the exposure of susceptible individuals to symptomatic and asymptomatic infected individuals by decreasing outdoor mobility (e.g., going out to movies, concerts, and restaurants, assembling in large groups) and encouraging social distancing. (e.g., 1m-2m physical distancing) [3,4].

Unlike the widespread and proactive implementation of lockdowns and physical distancing measures, the usage of masks varied widely across countries. Some countries quickly adopted guidelines for mask usage (e.g., Malaysia, Singapore, Taiwan, and Thailand) while others did not recommend using face masks unless sick [5,6,7]. Indeed, the World Health Organization updated its mask-wearing guidelines only on June 5, 2020 [8], to recommend that “The general public should wear non-medical masks where there is widespread transmission and when physical distancing is difficult, such as on public transport, in shops or in other confined or crowded environments.”. Due to these changing guidelines and uneven implementations, mask-wearing varied dramatically across countries and over time during the early phases of the pandemic [9].

Multiple studies have investigated the impact of various governmental NPIs [3,10,11,12,13], that encourage physical distancing and other restrictions. In each case, the studies find that NPIs and physical distancing reduce the transmission of COVID-19. Studies on the effectiveness of face masks [14,15,16] also show that face masks could contribute to the mitigation of COVID-19. However, a recent study [17] uses a randomized control trial to investigate the effect of masks. The authors find that infection with SARS-CoV-2 occurred in 1.8% of the participants in the treated group (recommended masks for 3 hours per day) versus 2.1% of the participants in the control group. This result, a difference of about 17% over a 60 day period, appears to be statistically insignificant. Despite this conclusion, as noted by [18], the trial in [17] points to “a likely benefit of mask-wearing to the wearer—it did not examine the wider potential benefit of the reduced spread of infection to others—and this even in a population where mask-wearing isn’t mandatory and prevalence of infection is low.” In addition, the interventions, government policies,, individual measures and exposures to infection due to outdoor mobility seldom act in isolation. Treating these measures in isolation could lead to under- *or* over-estimation of their effectiveness at reducing the spread of the disease, biasing the assessments of the measure’s impact. In this study, we investigate the association of population-wide mask-wearing with the number of COVID-19 cases, concurrent with other individual and institutional measures.

In sum, because mask-wearing varied dramatically in early 2020, we restrict this study to examine the mitigating role that mask-wearing played during the early phases of the pandemic. Specifically, we expand on the current stream of research by simultaneously considering the effects of NPIs and outdoor mobility in combination with a population’s reported usage of face masks in public places in a reduced-form econometric model (see examples in [3] and [10]). Using data from 24 countries, we identify the effect of each measure by exploiting the country-wise differences in the (1) percentage of the population who report wearing a face mask in public places (YouGov Survey Data [9]), (2) outdoor mobility across multiple categories such as Parks and Transit Locations (using Google Mobility Reports [19]) and (3) the NPI implementations (using CoronaNet-Project [1]). The results re-affirm the importance of mask-wearing in combating the spread of COVID-19.

## 2. Methods

This study is a cross-sectional analysis of the effects of personal and governmental measures across 24 countries on mitigating COVID-19 disease spread. The data used in this study were collected from February 21, 2020, to July 8, 2020, representing 139 days of data for each country. All analysis presented in this paper uses publicly available data. Subsequently, we first present the data on the three measures, namely, mask-wearing, outdoor mobility and NPIs, and then discuss the model-based analysis.

### Key Variables of Interest

#### Mask-Wearing

We study the impact of mask-wearing behavior using survey data released by the Institute of Global Health Innovation (IGHI) at Imperial College London and YouGov [9] provide reported mask-wearing across multiple countries. The survey covers 26 countries (as of July 8, 2020), with around 21,000 people interviewed each week. Further details about the survey design can be found in Supplement S2.1. We restrict our analysis to 24 countries because two countries – China and Hong Kong – do not have publicly available data on outdoor mobility which we control for in this study. The data present global insights on people’s reported behavior in response to COVID-19. The dataset provides the percentage of population in each country who report wearing a mask in public places. Because these surveys were conducted at an interval of several days, we interpolate (linearly) to estimate the percentage of the population that would wear masks in public spaces for days when the data were unavailable (Figure 1). We use the significant variation of mask-wearing across countries to measure the association of people reporting mask-wearing and the spread of COVID-19.

**Figure 1.**
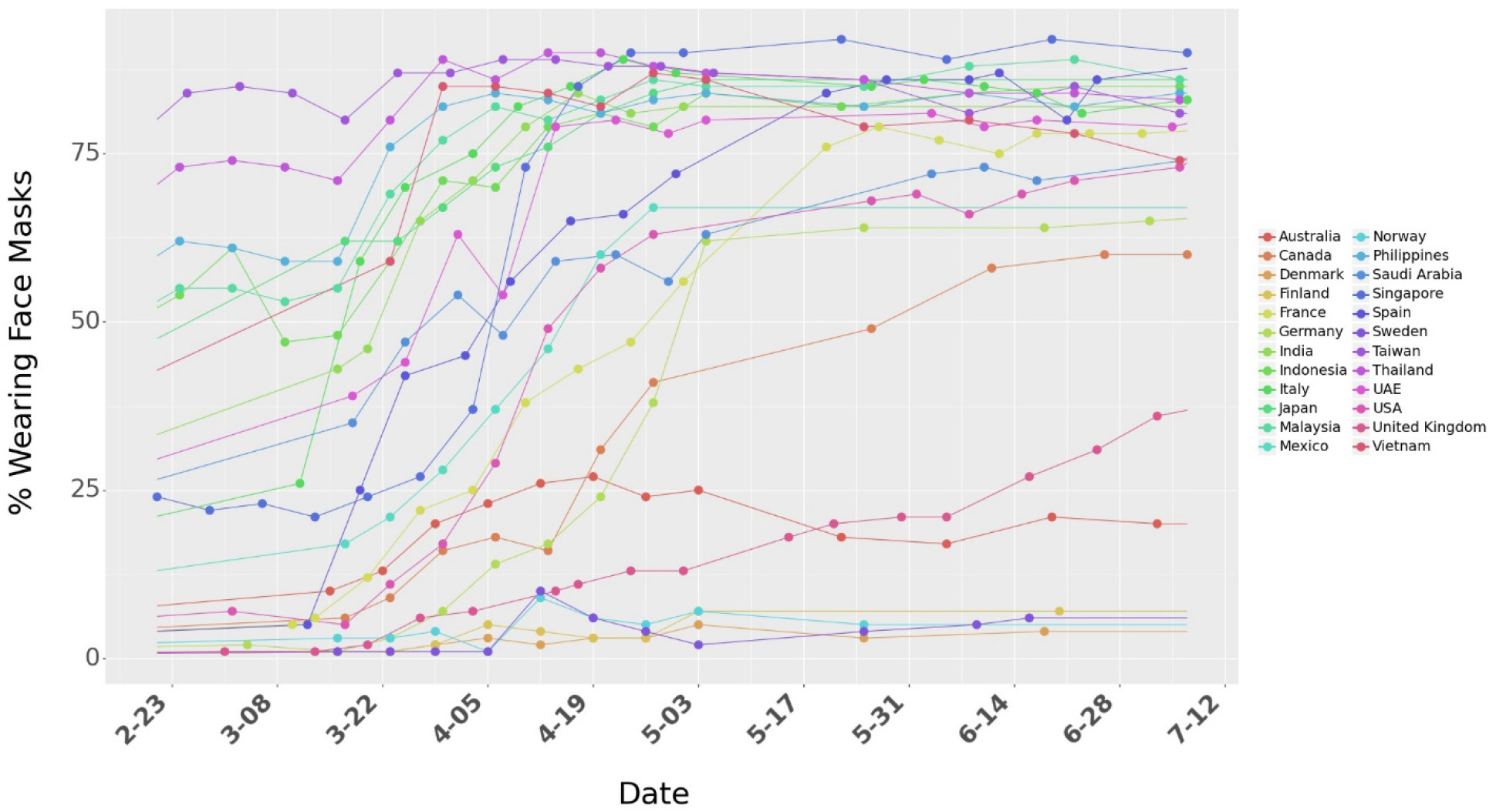
Percentage of people who say they are wearing a face mask in public spaces

#### Outdoor mobility

Google Community Mobility Reports provide data on relative mobility changes with respect to an internal baseline across multiple categories namely, retail and recreation, groceries and pharmacies, parks, transit stations, workplaces, and residential (Figure 2). A summary of the community mobility is shown in Table S2 in the Supplement. Apart from the Google Mobility reports, we also utilize mobility data from Apple to test the robustness of the model to different measures of mobility. We note that neither Google nor Apple provides absolute measures of mobility, but rather present relative changes with respect to benchmarks they use internally. Finally, drops in mobility could be driven by both individual actions (e.g., cautious behavior) as well as institutional actions due to NPIs enacted by governments. To control for mobility declines due to institutional actions, we also include country-specific interventions enacted both nationally and provincially.

**Figure 2.**
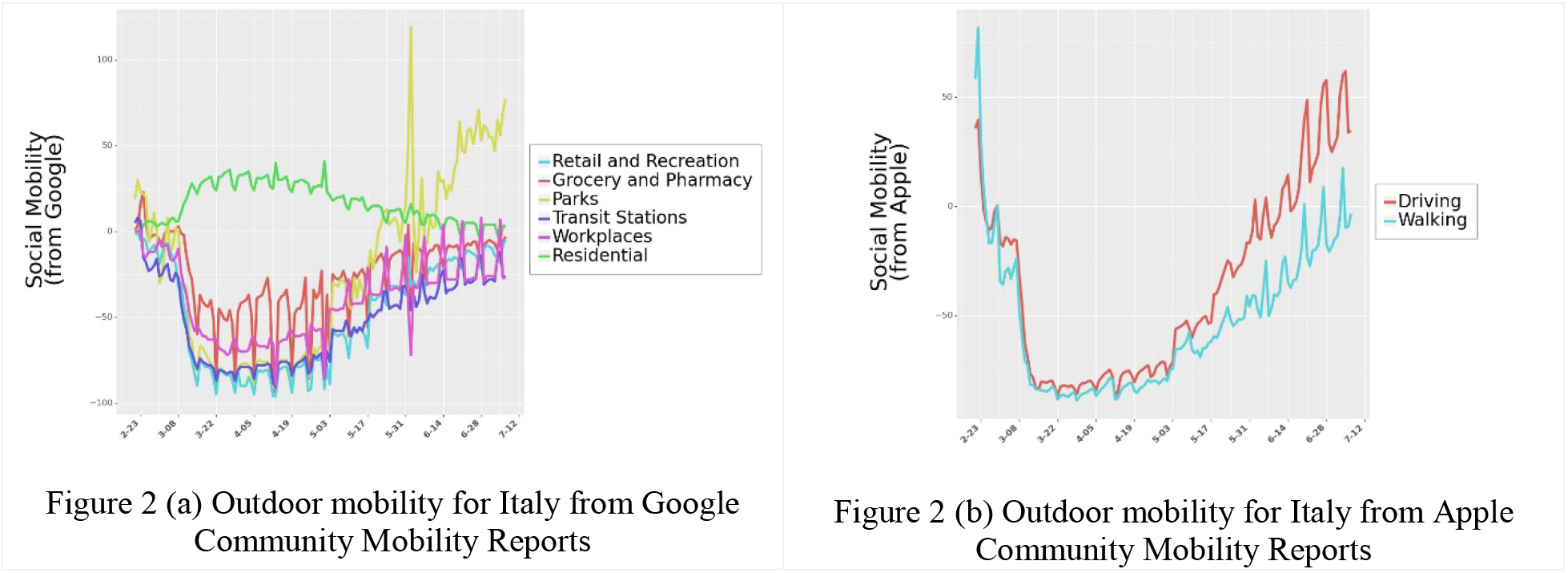
Outdoor mobility from Google Mobility Reports and Apple Mobility Reports for Italy. Outdoor mobility for all 24 countries is shown in Figure S8 in Supplementary.

#### Non-Pharmaceutical Interventions

Governments across the 24 countries enforced different policies to control the spread of COVID-19. Prior research has shown that these policies played a significant role in reducing human to human physical contacts and led to a slowdown in the spread of the disease. However, these policies were implemented at different levels, some nationally, some provincially. We use data from COVID-19 Government Response Event Dataset [1] to control for government policies in estimating the effect of masks. Figure S10 lists the types and counts of national and provincial government policies implemented across the 24 countries we consider in this study. The dataset contains 5,816 entries on policies at the National and Provincial levels. Finally, the inclusion of these interventions helps control for some of the observed drops in mobility that are not necessarily associated with individual actions but by the presence of institutional policies. Detailed information about the interventions are included in the Supplement (Section S2.5).

### Covariates

Because the data span multiple countries and weeks, we include time and country fixed effects in the model. The model controls for country-level heterogeneity using fixed-effects, where the variable for a country assumes a value of one if the data considered are specific to that country, and zero otherwise. This allows for control of country-level characteristics that are not in the model and helps reduce the errors due to omitted variables in our analysis. In addition to country-level differences, we also control for time-based differences (e.g., people are more aware and cautious over time) by incorporating time-fixed effects, where the variable *Week*_*t*_ takes a value of 1 if the data are from week ‘*t*’ (where *t* =1 represents the first week for a given country in the data). In addition, we control for each country’s testing capability (Figure 3a) by accounting for the total number of daily tests in the country. Finally, we also control for actions people take to educate themselves by including the Google Trends (Figure 3b) data for the search term ‘coronavirus’.

**Figure 3.**
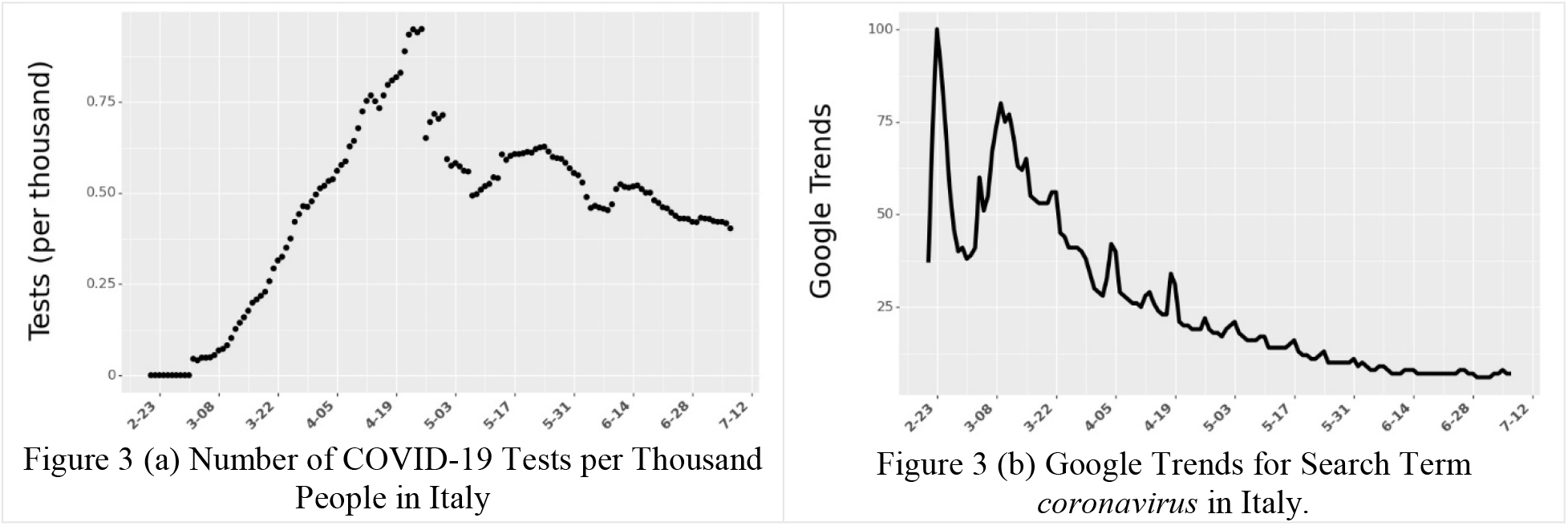
Number of COVID-19 Tests and Google Trends (per thousand people) for Italy. Data for all the 24 countries is shown in Figure S13 and Figure S14 respectively in Supplementary.

### Outcome Variable

Data for the number of active daily cases in each country were obtained from the Johns Hopkins University School of Public Health [20]. We use a seven-day moving average of cumulative confirmed cases and cumulative recovered cases to compute daily active cases and daily growth rate. The dataset aggregates this information across multiple national, state, and local health departments within each country. The daily growth rate is then related, through a reduced-form econometric model, to the independent variables described earlier. We describe the derivation in the Supplement (Section S1.1). We illustrate the daily cases and growth rate for one country, Italy, in Figure 4.

**Figure 4.**
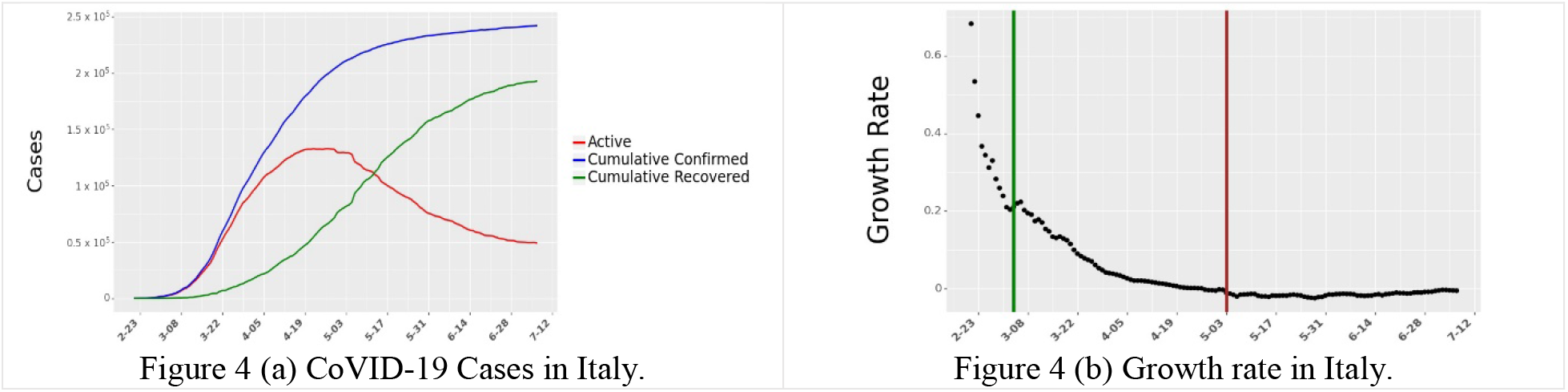
Daily Active cases and growth rate in Active cases for Italy. The vertical green line in (b) shows the start of data collection for Italy. The vertical red line in (b) shows the end of 60 days of data collection for Italy. Data for all 24 countries are shown in Figure S6 and Figure S7 respectively in the Supplement.

### Analysis

A reduced form econometrics model was used to relate the growth rate of daily active infections to the independent variables described earlier. Similar models have been used by [3] to determine the effect of anti-contagion policies on the spread of COVID-19. In brief, the model assumes that the daily growth rate (ratio of active infections today to active infections the day before) is affected by institutional measures such as NPIs as well as individual measures such as outdoor mobility and mask-wearing. The covariates listed above help control for other factors that could affect growth over time. Because the epidemiological parameters for new diseases such as COVID-19 might not be well understood, reduced form techniques allow for the estimation of the impact of governmental and personal measures to help contain the spread of the virus. To filter out the high variation in growth rates when the number of cases is very low at the beginning of the pandemic, our model initializes when a country reaches 20% of peak new cases as observed by July 8, 2020,. For robustness, we also test other starting times in the Supplement and find results in line with the ones presented here. The Supplement also provides further details about the methodological approach and model formulation used in this paper. We provide some brief notes on the operationalization of the independent variables and the model initialization below:

1. Responses to the survey about mask-wearing are subject to biases. For example, individuals might overestimate the efficacy of their mask or their wearing pattern. To alleviate some of these concerns, we compute the natural log of the mask-wearing variable to discount its impact on the growth rate of daily active cases. This transformation yields a curve that grows at a slower rate as the values of mask-wearing increase, thereby diminishing the impact of higher levels of mask-wearing. We also test for other functional forms (square-root and linear) and present those results in the Supplement (Table S7).
2. Due to the high correlation across the different mobility data categories obtained from Google, we only include *Mobility: Parks* and *Mobility: Transit Stations* in the model. Because we are interested in determining the impact of mobility in general, the two mobility variables suffice in capturing the individual’s movement patterns during this time. In Supplement S3.4, we present results including other types of mobility and also run the model with Apple Mobility data in the place of Google Mobility Reports.
3. The CoronaNet dataset from [1] collected information on all the government policies introduced by different countries across the world. They categorized the policies into 19 different policy-types. We use their categorization in the model. From February 21, 2020 to July 8, 2020, we check if a policy *p* was implemented in a country *j* on day *t*. If the policy was implemented, we assign a value of 1 to *s*_*j,t,p*_, where *s* represents the level of policy coverage. If the policy was introduced at a provincial level, we normalize *s*_*j,t,p*_ by the population of the state. Because several policies were introduced at the same time or close together, they too suffered from collinearity issues. To minimize multicollinearity issues, we choose only a specific set of policies to include in the analysis. The Supplement (Section S2.5) discusses this selection mechanism.
4. Due to uncertainty of the lag in COVID-19 incidences and the difficulties in detection during the early days of the disease [21], similar to prior research we tested the focal model across multiple lag periods (*shift*) from zero to 14 days and for different initialization thresholds (*th*) for each country (zero percent to 20% of a country’s peak daily cases by July 08, 2020). We chose the best *shift* and *th* values using a k-fold cross-validation process (k=5). The chosen model had the highest maximum likelihood estimate of the data as well as the lowest prediction error. We discuss this procedure in the Supplement (Section S4.1). The results presented in the next section correspond to a model with a shift of nine days and a *th* of 20% of peak new cases by July 12, 2020. Finally, the model was estimated on 1,422 observations across 24 countries. We restrict our analysis to the first 60 days after model initialization based on *th*. However, we test the robustness of the findings for other lengths of data. This allows for greater variation in mask usage within the data.

In the next section, we describe our results and their policy implications.

## 3. Results

The results indicate that individual measures such as mask-wearing and outdoor mobility combined with institutional measures (NPIs) play a role in mitigating the spread of COVID-19. The estimates from the focal reduced form model for these measures and their corresponding confidence intervals are shown in Figure 5. The full table of results, along with results for all robustness checks are provided in Supplement (Section 3). We first list the results of the key measures we consider and then discuss their implications.

**Figure 5.**
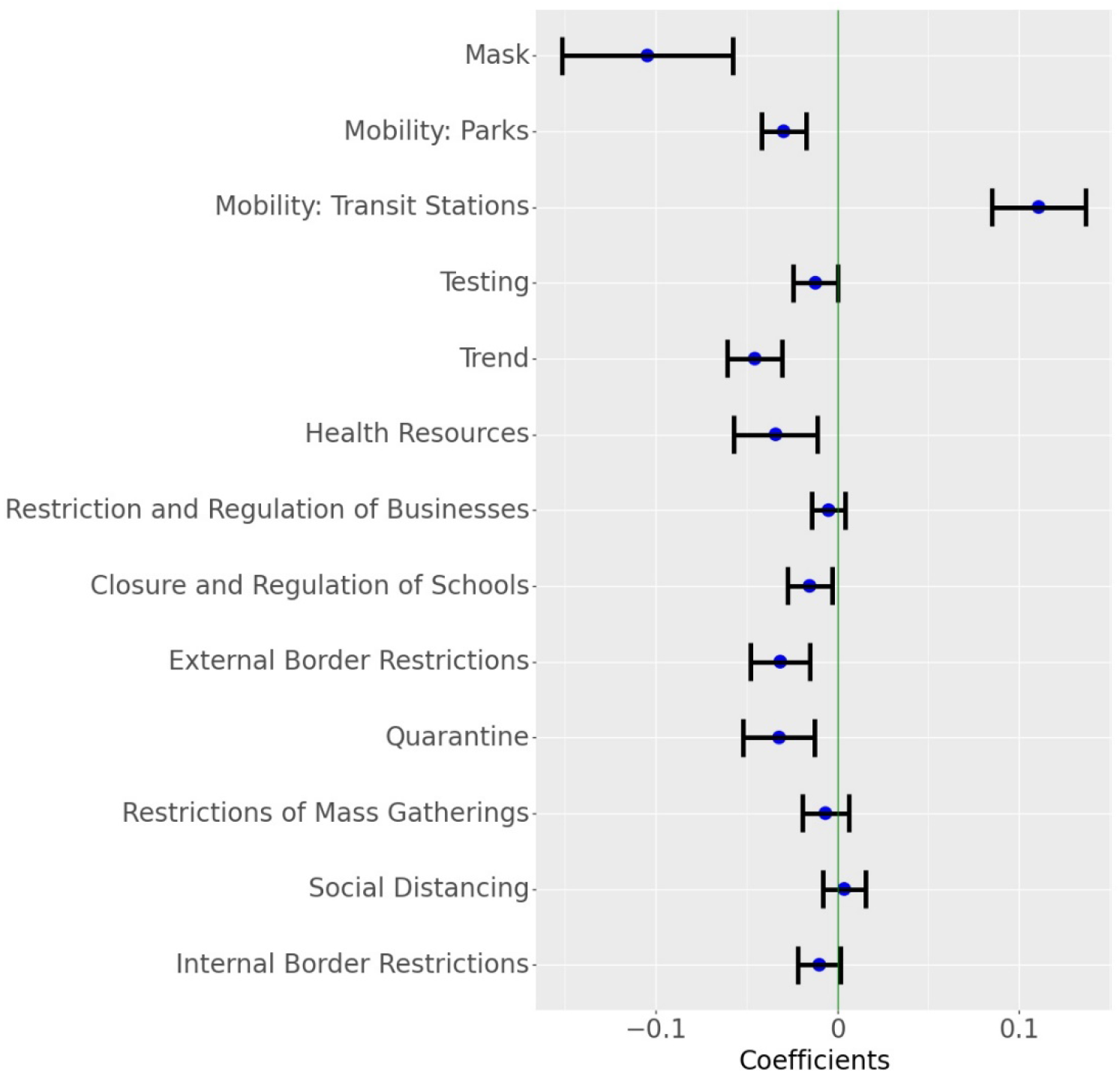
Parameter estimates for the Growth rate model. The blue dot indicates the point estimates and the horizontal lines indicate 95% confidence interval around the estimate (Masks transformed as ln(1+mask)). Results for different transformations of Mask and other covariates are shown in Figure S16.

### Mask-Wearing

The model finds that a reported mask wearing of 100% associates with an average 7% (95% CI: 3.94% — 9.99%) drop in the daily growth rate of COVID-19 cases. While this daily effect appears small, 100% reported mask-wearing leads to approximately 88.5% (95% CI: 68.7% — 89.2%) decline in active cases over a 30 day period when compared to the situation where 0% of the people report wearing masks (all else remaining the same across the two scenarios). Modifying the functional form of the mask variable did not appreciably change the association. For example, in the linear model, masks are associated with an average 8.69% (95% CI: 5.63% — 11.66%) drop in daily growth rate and for the square root model the expected daily drop in growth rate was 7.89% (95% CI: 4.81% — 10.87%). The stability of the results indicates that mask-wearing plays a significant role in mitigating the spread of the disease. Figure 5 also illustrates that widespread mask-wearing, as an intervention by itself, has the largest (by magnitude) association with the growth rate of active COVID-19 cases. Figure 6 plots the ratio of active cases under different proportions of respondents who claim to wear masks as against no mask wearing and for various time periods.

**Figure 6.**
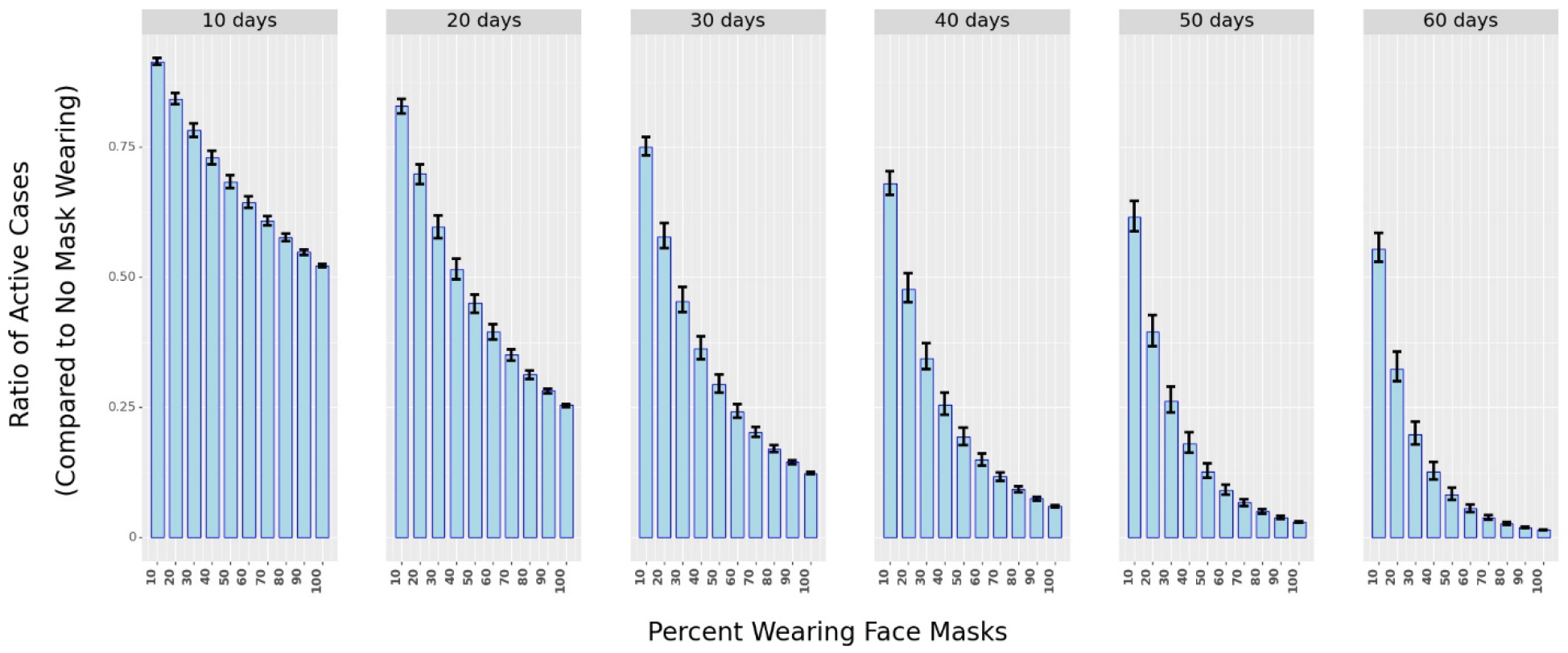
The ratio of Active cases under different percentages of mask-wearing in public spaces as compared to 0% mask-wearing over different periods (in days). The shaded bars represent in ratio while the black vertical lines represent the 25^th^ and 75^th^ percentile of the ratio (from simulations using Krinsky-Robb method)

### Mobility and NPIs

As expected, the model finds that a rise in mobility links with a rise in the number of cases. Specifically, the selected mobility variables associate with a *combined* 8.1% (95% CI: 5.6% - 10.6%) drop in daily case numbers. Similarly, we find that the implementation of NPIs are also associated with a drop in daily growth rates across countries. After accounting for mobility declines, the NPI measures ‘Quarantine’, ‘External Border Restrictions’, and ‘Closure and Regulation of Schools’ link with the highest declines in the growth rate of daily active cases. Overall, all NPIs included in the model led to a decrease in the growth rate of COVID-19. This finding confirms multiple studies that investigated the effects of NPIs at limiting the spread of COVID-19 [10,13,22]. Overall, we find that if the NPIs were enacted uniformly across the whole country, then the *combined* association of the NPIs with the decline of growth in daily cases of COVID-19 would average 13% (95% CI: 9.2% - 16.2%). We determine the combined effect using the Krinsky-Robb method, a Monte Carlo simulation used to draw samples from a multivariate normal distribution. Supplement S3.1 provides more details on this method.

#### Controlling for Endogeneity using Control Functions

Due to nearly concurrent enactments and blanket coverage of policies and precautionary behaviors within countries, the individual (e.g., masks, limiting mobility) and institutional (NPIs) measures correlate in time. This precludes the causal identification of each measure’s effect on disease mitigation. In other words, because mask-wearing, mobility reductions, and NPIs occur at similar times, their effects are intertwined and difficult to determine separately. For some variables such as mobility and NPIs, we lack the necessary data to fully control for these issues. In the case of mask-wearing, even though we cannot eliminate all the possible endogeneity issues, we attempt to alleviate some of the concerns of confounding variables by employing control functions [23]. As noted in [24], control functions make the intervention exogenous in a regression equation. To create a control function, we use country mortality data for *prior outbreaks* (in the country) of SARS, H1N1, and MERS in each country as instrumental variables to predict the proportion of mask-wearing in each country (see Supplement S4.5.2 for more details). We posit that exposure to prior pandemics would have resulted in a more aware populace that could be amenable to precautionary behaviors such as mask-wearing. Next, we compute the control function by determining the predicted mask-wearing residuals (computed via determining “Predicted Mask-Wearing minus Reported Mask-Wearing”), allowing for better identification of the effect of reported mask-wearing on COVID-19 case numbers.

Using this procedure, we find that if 100% of the population claimed to wear masks, then mask-wearing relates with an average 4.95% (95% CI: 2.26% — 7.53%) drop in daily growth rate of COVID-19, when compared to zero percent reported mask-wearing. Over 30 days, this translates to a 70.4% (95% CI: 62.3% — 72.7%) drop in new COVID-19 cases. While we are careful to note that this estimate could still be affected by confounding variables, this result lends stronger support to the magnitude of the disease mitigation that mask-wearing in the general population provides. In summary, widespread mask-wearing leads to a significant decline in the spread of COVID-19.

#### Robustness Checks

To help determine the accuracy and stability of the results we run several robustness checks (see Supplement S4):

1. We vary the lag period (*shift*) from 0 to 14 days. The results show that the estimates of the individual and institutional measures are relatively stable.
2. We also vary the length of time we consider in the analysis. The model considered 60 days of data for each country. We vary this to estimate the model on 35, 45, 55, 65, 75, and 85 days of data. We find that the results remain stable to these variations.
3. We replace Google mobility data with Apple mobility data. The model estimates remain robust to this change.
4. We vary the functional form of how mask-wearing relates to the spread of COVID-19. The results are not statistically different in these cases.
5. We also test the robustness of the analysis by modifying the data using exponential smoothing. Specifically, for any day *t*, the focal model in Equation (1) ignores the value of the independent variables from days *t-shift+1* to t (discussed in Figure S1). In the model we use for the robustness check, we do not ignore values between *t-shift* and *t* and use exponential smoothing to average the intervening data. Finally, we also modify the interpolation method from linear (current) to quadratic. We find that the results are stable to all these modifications.

The Supplement details all the robustness checks and simulations as well as their results.

## 4. Discussion

Over the past few months, several studies have investigated the efficacy of masks at minimizing droplet dispersion [25,26] and the potential consequences of their use [14,27] in the general population. Although a randomized control trial on the efficacy of face mask usage appears to indicate inconclusive results in the general population [17], [15,16] provide evidence for the benefits of face mask usage through a systematic review of the multiple observational studies and the evidence thus far. While the type of face mask as well as the timing and length of use can affect its efficacy, its use as a precautionary principle has been strongly advised [28]. Despite the abundant scholarly and some anecdotal evidence [29], face mask use in some countries like Sweden and the United States remains controversial [30,31,32]. Additionally, as observed in the data, even in countries where masks do not face similar headwinds and as support for mask usage gathers further evidence, face mask use is not as commonplace (e.g., Denmark, Norway, Sweden, Finland), even as a precautionary principle.

This study links the growth rate in active cases of COVID-19 in a country, to a population’s reported wearing of face masks in public places over time. The model also includes other measures that could simultaneously impact the spread of the disease as face mask usage changes over time. After accounting for these measures and controlling for other covariates, the results indicate that reported face mask use is associated with a decline in the growth of COVID-19. More precisely, if 100% of the population claimed to wear masks, then mask-wearing is associated with an average 7% decline in the growth of daily active cases of COVID-19. This association persists across multiple robustness checks and model formulations. A decline of 7% corresponds to an 88.5% drop in the number of active cases 30 days later. Taken together with the other measures (mobility changes, NPIs), the combined association of individual and institutional measures on the decline in the growth rate of daily active cases of COVID-19 is 28.1% (95% CI: 24.2%-32%).

### Limitations

Countries enacted multiple NPIs simultaneously. This precludes us from identifying the effectiveness of NPIs separately. Second, the mobility data provided by Google and Apple are only indicative of the relative changes from a benchmark, so their association with disease spread should be interpreted with precaution. Third, we rely on the accuracy of data collected by third parties. Inconsistencies in testing, reporting, and recording of the data could lead to errors in the results obtained. Additionally, mask-types and mask-wearing patterns could vary across countries, individuals, and over time. This limitation affects all observational COVID-19 population-based studies.

## 5. Conclusions

The population-wide usage of face masks as a preventative measure against the transmission of COVID-19 varies widely across countries. Using data from 24 countries, this study finds that face mask usage associates with a decline in the growth rate of daily active cases of COVID-19. Over a 30-day period, mask-wearing associates with an 88.5% decline in number of daily active cases. This result re-affirms the prominent importance of masks in combating the spread of COVID-19.

## Supporting information

Supplementary Analysis

## Data Availability

All codes have been written in python 3.7 programming language. All data and codes are available at open sourced Github repository at https://github.com/ashutoshnayakIE/COVID-masks. Data is stored in pythons numpy format. However, the raw data can be procured from the sources mentioned in the references below (link for raw data set is also provided in the Github repository).

