## Supplementary Analysis for "The Impact of Mask-Wearing in Mitigating the Spread of COVID-19 During the Early Phases of the Pandemic"

**Supplementary Material for**
**Mask-Wearing During the COVID-19 Pandemic**

Ashwin Aravindakshan[1*]
Jörn Boehnke[*]
Ehsan Gholami[*]
Ashutosh Nayak[*]

[*]University of California, Davis

### S1. Method

In this Section, we provide a detailed description of the reduced form econometrics model considered in our analysis. The model is derived from Susceptible-Infectious-Recovered (SIR) epidemiology models.

#### S1.1 SIR Growth Rate Model

Similar to the work by Hsiang et al [1], we employ a reduced form econometrics technique that relates the growth rate of active COVID-19 cases to the individual and institutional measures such as masks, social mobility and governmental Non-Pharmaceutical Interventions (NPIs). Growth rate in econometrics is defined as the first difference in log of economic outputs in different time periods. The growth rate model is a well-established method in econometrics where growth rates of economic output can be affected by different factors e.g. policy. Similar to economic output, we model the growth rate of daily active cases and estimate how it is affected by wearing masks in public spaces, social mobility and NPIs. The method also has roots in epidemiology models – SIR (Susceptible, Infection, Recover). We do not consider deaths and reinfection in our analysis.

Equation S1-S4 describe the SIR model where $S_{j,t}, I_{j,t}, R_{j,t}$ show the active susceptible, infectious and recovered population at time $t$ in country $j$. $\beta_j$ is the rate of transmission and $\gamma_j$ is the rate of recovery in country $j$. Since we do not consider reinfection and deaths, $\gamma_j$ can be considered as the rate of removal from infectious population. $N_j$ is the total population of the country $j$. Equation S1 shows how infections spread from the infectious individuals to susceptible individuals. Equation 2 shows how infectious population changes over time as some susceptible individuals contract the disease while some already infectious individuals recover from the disease and test negative. Equation S3 shows how the number of recovered individuals increase over time as individuals recover after testing negative for the virus. Equation S4 is a feasibility constraint which ensures that the total population is accounted for in the model. Addition of Equations S1 – S3 yields Equation S4.

$$\frac{dS_{j,t}}{dt} = \frac{\beta_j I_{j,t} S_{j,t}}{N_j} \qquad \text{(S1)}$$

$$\frac{dI_{j,t}}{dt} = \frac{\beta_j I_{j,t} S_{j,t}}{N_j} - \gamma I_{j,t} \qquad \text{(S2)}$$

$$\frac{dR_{j,t}}{dt} = \gamma_j I_{j,t} \qquad \text{(S3)}$$

$$\frac{dS_{j,t}}{dt} + \frac{dI_{j,t}}{dt} + \frac{dR_{j,t}}{dt} = 0 \qquad \text{(S4)}$$

Since we model only the growth rate in the total confirmed cases, we consider Equation S2 in our analysis. Assuming $S_{j,t} \approx N_j$, we can rewrite Equation S2 as shown in Equation S5. It can be solved by integration as shown in Equation S6. If we consider daily growth rate ($t_2 - t_1 = 1$), Equation S6 can be simplified as shown in Equation S7, where $g_j$ is the growth rate and it is given by $\beta_j - \gamma_j$.

$$\frac{dI_{j,t}}{dt} = (\beta_j - \gamma_j)\, I_{j,t} \qquad \text{(S5)}$$

$$\int_{t_1}^{t_2} \frac{dI_{j,t}}{dt} = \log\left(I_{j,t_2}\right) - \log\left(I_{j,t_1}\right) = \left(\beta_j - \gamma_j\right)(t_2 - t_1) \qquad \text{(S6)}$$

$$\log\left(I_{j,t}\right) - \log\left(I_{j,t}\right) = \; g_j \qquad \text{(S7)}$$

Wearing face masks, reducing social mobility and implementation of NPIs can alter the growth rate by changing $g_j$. We consider a common $\beta \; \forall j$ instead of solving Equation S8 individually for countries. We include country fixed effects ($countries_j$) to account for country specific heterogeneity in $g_j$. Growth rate can be modeled as shown in Equation S8 ($P$ is set of policies; $M$ is the set of indicators of social mobility, $W$ is set of weeks during the period of our analysis and $J$ is the countries in our analysis). $mobility_{j,t,m}$ is the $m^{th}$indicator for social mobility, $week_{j,t,w} = 1$ if day $t$ in country $j$ is in week $w$ after the initialization point for country $j$. To account for other factors, we consider several control variables (e.g. testing, Google Trends, fixed effect for week) as we discuss in the next section.

$$g_{j,t+shift} = \theta_0 + \theta_m.mask_{j,t} + \sum_{p\in P} \theta_p policy_{j,t,p} + \sum_{m\in M} \theta_m mobility_{j,t,m} + \theta_e.testing_{j,t} + \sum_{w\in W} \theta_w week_{j,t,w} + \theta_r.trend_{j,t} + \sum_{j\in J} \theta_j countries_j + \epsilon_t \qquad \text{(S8)}$$

We use government announcements on health resources, health monitoring, health testing and tests conducted (per thousand individuals) to account for increased awareness and testing over time. We also use Google Trends on a keyword *coronavirus* to account for public self-awareness. We discuss NPIs, Testing and Google Trends later.

The econometrics approach of using the growth rate to estimate the effects of masks, social mobility and NPIs has several advantages. The model can estimate the effect of change in the exogenous independent variables on the dependent outcome variable (growth rate). Since right hand side of Equation S5 can be empirically calculated, it does not explicitly require the knowledge of relationship between exogenous variables and $I_{j,t}$. Thus, the model does not need to know the link between masks, NPIs and social mobility on daily active cases (or cumulative conformed cases) but can still estimate their effect on the growth rate of infectious cases. Using the growth rate, $I_{j,t}$ can be estimated by integrating it from time period 0 (or using previous integration up to day $t - 1$). Thus, this model is a forward-looking model.

The model is also able to handle underreporting in COVID-19. In COVID-19 pandemic, data for an individual is recorded only when they are tested. Total confirmed cases (deaths and recovered cases) in publicly available datasets only provides information on the individuals who got themselves tested. Due to various reasons e.g. lack of testing, lack of motivation to get tested or lack of visible symptoms in symptomatic cases, it is being estimated that there is a massive underreporting in total confirmed positive cases. However, the growth rate model is agnostic to underreporting as it models the first difference in the log of confirmed cases. If the underreporting remains constant, $I_{j,t}$ and $I_{j,t-1}$ can be multiplied with a constant over the time period considered and it would not affect our estimation of growth rate (Equation S5).

Multiple studies have reported delays between the association of policies with COVID-19 spread [1]. This delay could be due to several reasons. One of the most commonly noted reasons is the incubation period (time between getting infected and onset of symptoms/knowing that individual is confirmed for COVID-19). During the incubation period, an individual may be asymptomatic. Incubation period is estimated to be 4 days to 14 days [2]. Another reason could be the testing time – time it takes to get the confirmation of results. To model this delay, we use a lag variable. We use cross validation method to find the lag with the best fit for the data. We test and observe that the model performs best at a lag of 9 days.

#### S1.2 Robustness and Control Function Models

*Robustness Model*: As a robustness check to the growth rate model discussed above, we also use an exponential smoothing model to estimate the effect of masks, social mobility and NPIs to validate the results from the base model in Equation S9. In this model, we use exponentially smoothed data for right hand side of Equation S8 for the past $w$ days, without using the $lag$ variable. It can be written as shown in Equation S10 where $< x >_w$ is exponentially smoothed over the last $w$ days. Smoothing function is shown in Equation S11. This method considers data in the recent future of day $t$ instead of considering all the data leading up to day $t$ or data observed $lag$ days before as in the growth rate model in Equation S9. This model is analogous to counting the number of days in which a policy was active in the past $w$ days. We use exponential smoothing to include the lag effect of masks, NPIs and mobility on the growth rate.

$$\overline{g_{J,t+shift}} = \theta_0 + < \theta_m . mask_{j,t} >_w + \sum_{p \in P} \theta_p < policy_{j,t,p>_w} + \sum_{m \in M} \theta_m < mobility_{j,t,m} >_w$$
$$+ \theta_e < testing_{j,t} >_m + \sum_{w \in W} \theta_w week_{j,t,w} + \theta_r < trend_{j,t} >_w + \sum_{j \in J} \theta_j countries_j + \epsilon_t \quad \text{(S9)}$$

$$< x >_w = \frac{\sum_{l=1}^{w} 0.8^{w-l} x_{t-w}}{\sum_{l=1}^{w} 0.8^{w-l}} \quad \text{(S10)}$$

Figure S1 shows the idea behind the exponentially smoothed model. In the growth rate model, we give importance to events that happened at a lag of $shift$ days. Thus, on day $t$, we assign 0 weights to data from $t - shift + 1$ to $t$. In exponentially smoothed model, we assign an exponentially reducing but non-zero weights to data for days $t - shift + 1$ to $t$.

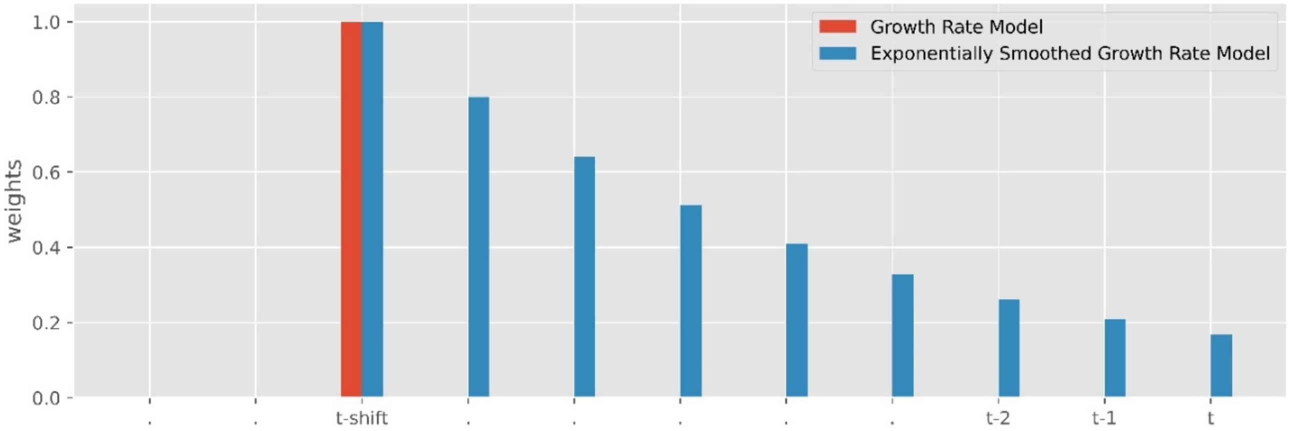


Figure S1. Weights given to different data points for two models – growth rate model in Equation S9 (red) and exponentially smoothed growth rate model (blue) in Equation S10. Exponentially smoothed growth rate model consider all the data points in recent history to day $t$ instead of considering the events only on day $t - shift$. Weights decrease as we move closer to day $t$ to incorporate the delay in observing the effect of events in the recent future.

*Control Function Model*: We also consider a control function approach to check the robustness of the mask parameter from Equation S8. Countries have had different experiences with air borne diseases due to multiple outbreaks in the past e.g. Severe Acute Respiratory Syndrome (SARS), Middle East Respiratory Syndrome Coronavirus (MERS-CoV) and H1N1 Influenza (Swine Flu). Countries with severe outbreak of these air-borne virus were quick to adopt to wearing face masks in public. This could potentially confound with the effect of mask discussed in this paper. So, we use a control function approach to isolate the effect of masks. Since the growth rate in COVID-19 is independent of number of deaths per thousand people from SARS, MERS and H1N1, it may affect the percentage of population with mask wearing $mask_{jt}$ but does not affect the growth rate. This allows us to use deaths from previous diseases as a control function. Total deaths per thousand people for different countries is given in Figure S2.

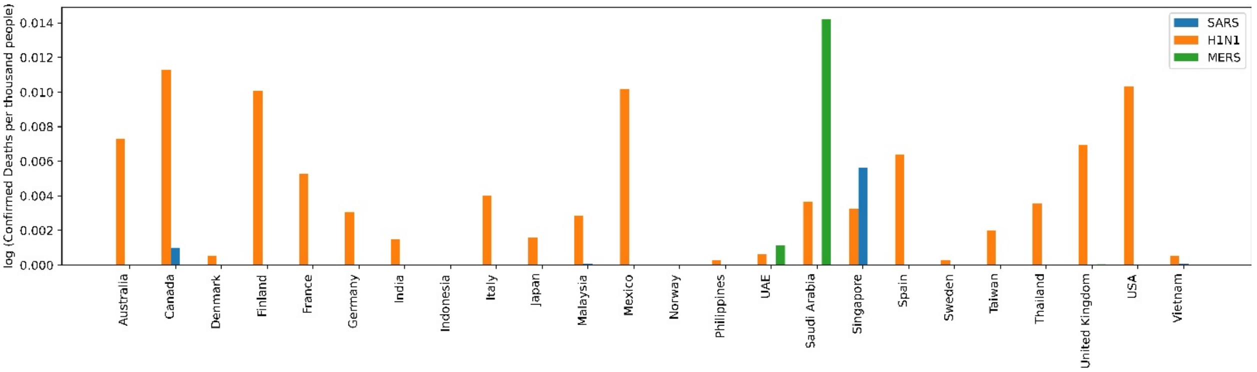


Figure S2. Logarithm of number of confirmed deaths with SARS, H1N1 and MERS-CoV per thousand people across 24 countries considered in this study.

In the control function model, we first predict average mask wearing in country $j$ $\widehat{mask}_j$ using the number of deaths from SARS, H1N1 and MERS in country $j$ as covariates in ordinary least square linear regression estimation. We use $d_{j,dis}$ where $dis \in \{SARS, H1N1, MERS\}$ as predictor variable where $d$ is the number of deaths per thousand people in country $j$. Specifically we use, $d_{j,dis} = 1\ |d_{j,dis} > median(D_{dis})$ for deaths per thousand people in country $j$. The model to predict $mask_{j,t}$ is shown in Equation S12. After estimating $\widehat{mask}_{j,t}$, we use the error $mask_{j,t} - \widehat{mask}_j$ in Equation S9 as a covariate. Control function model is shown in Equation S13.

$$\widehat{mask_J} = d_{j,SARS} + d_{jH1N1} + d_{j,MERS} + \epsilon_m \qquad \text{(S11)}$$

$$g_{j,t+shift} = \theta_0 + \theta_m.mask_{j,t} + \sum_{p \in P} \theta_p policy_{j,t,p} + \sum_{m \in M} \theta_m mobility_{j,t,m} + \theta_e.testing_{j,t} + \sum_{w \in W} \theta_w week_{j,t,w} + \theta_r.trend_{j,t} + \sum_{j \in J} \theta_j countries_j + \theta_{md}(mask_{j,t} - \widehat{mask_J}) + \epsilon_t \qquad \text{(S12)}$$

Wearing face masks in public is common in many Asian countries, as compared to countries in Europe or America [3, 4]. One of the reasons is due to recent experience with air borne diseases. Another reason could be air pollution or a culture of wearing face masks. We do not account for the different trends in wearing face masks among countries due to pollution or culture. However, we believe the country fixed effects could capture the country wise trends in wearing face masks.

### S2. Data Collection and Processing

We model the effect of wearing face masks, change in social mobility and government enforced Non-Pharmaceutical Interventions (NPIs) in the growth rate of infection. We select the countries with publicly available dataset for wearing face masks and community mobility. We collect data from February 21, 2020 to July 8, 2020. In this Section, we discuss the different datasets used in our analysis to isolate the effect of masks, social mobility and NPIs in the spread of the contagious SARS-CoV-2 (COVID-19 virus).

#### S2.1 Masks

We collect mask data from surveys conducted by YouGov [5]. YouGov is an international internet based market research company which specializes in opinion polls through online methods. YouGov used online surveys as their COVID-19 behavior change tracker. They conducted surveys periodically in some countries of the world to estimate the propensity of the percentage of people that wear face masks when they go out in public spaces. These surveys were conducted every week. Figure S3 shows the raw survey numbers from YouGov. As the surveys were conducted periodically and not every day, we use monotonic cubic splines to estimate the percentage of population that wear face masks in public spaces (Figure S4). Note that the online survey does not include data for type (quality) of masks or how people wear masks (insufficient quality or incorrect method of covering face masks e.g. touching the surface, not covering nose or mouth -- might not be effective in controlling the spread of virus). Thus, our estimation for effect of masks in this work would be an estimation of the behavior of wearing masks. In our analysis, we normalize the number for mask wearing such that $0 \leq mask_{jt} \leq 1, \ \forall j, \forall t$.

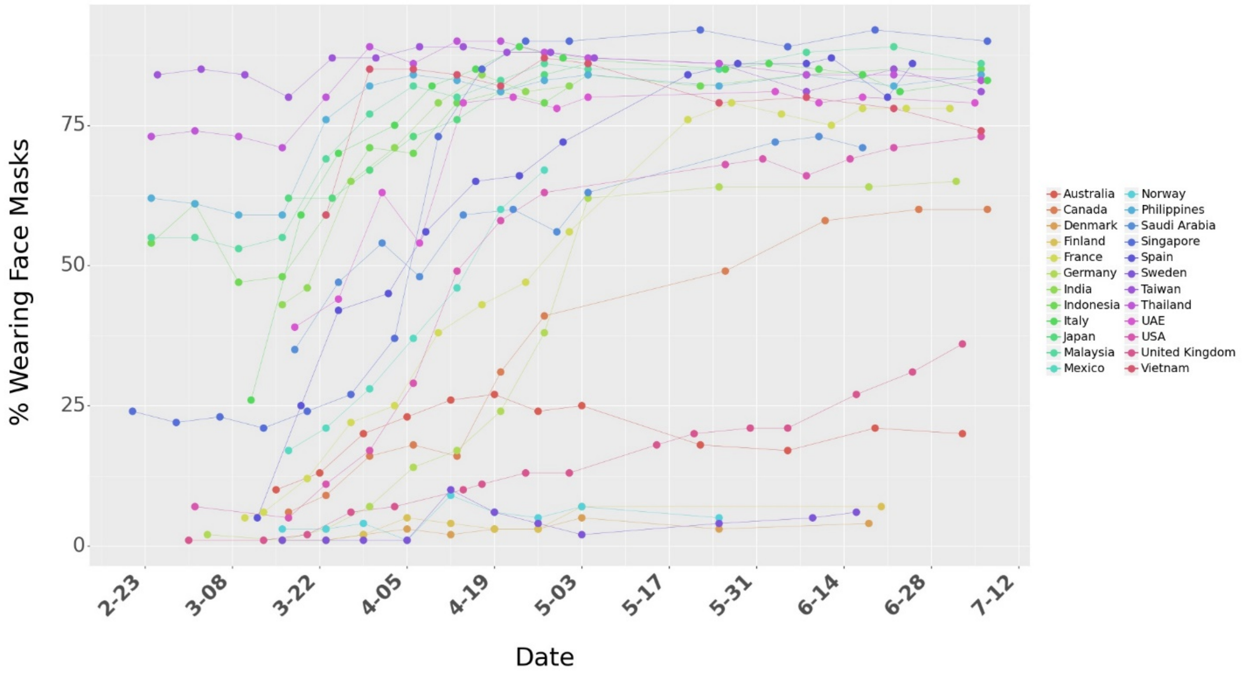


Figure S3. Raw data for surveys on percentage of people who say they wear a face mask when in public spaces. The dots represent the raw numbers from the survey data from YouGov. The lines are shown for better visualization.

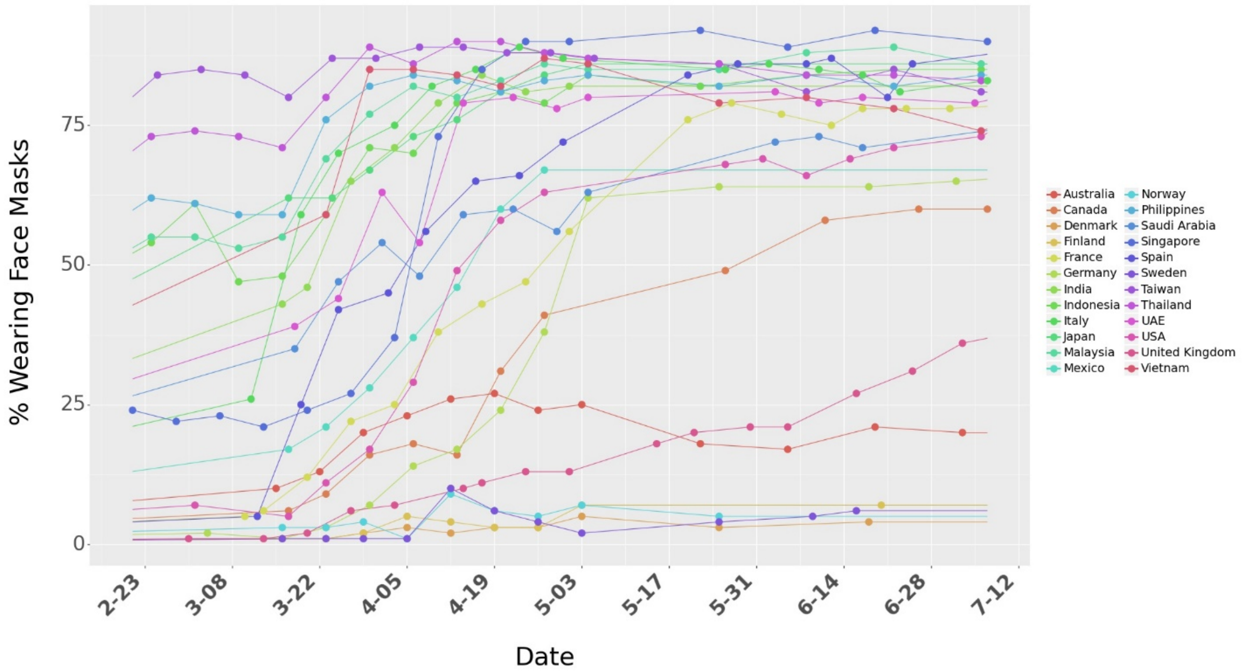


Figure S4. Survey data on percentage of people who say they wear a face mask when in public spaces. We use linear interpolation to consider mask numbers for days between surveys days. The dots represent the raw numbers from surveys.

### S2.2 Active Cases

We use the timeline for total confirmed cases and total recovered cases from Johns Hopkins Coronavirus Research Center [6] to find the daily active cases across different countries. We use the daily active cases to calculate the outcome variable of our model -- growth rate. Cumulative confirmed cases, Cumulative recovered cases and daily active cases for different countries is shown in Figure S5. We use a 7-day moving for daily cumulative confirmed cases and cumulative recovered cases.

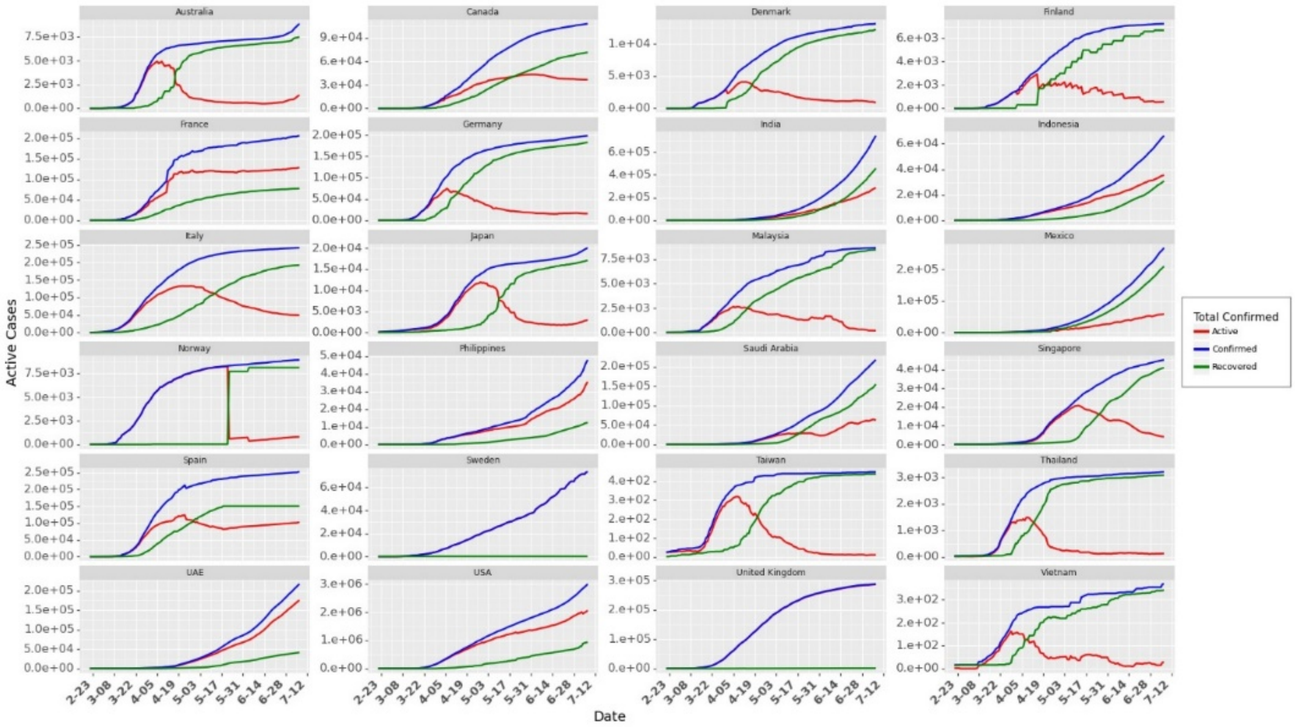


Figure S5. Cumulative confirmed cases, cumulative recovered cases and daily active cases across 24 countries. We observe data reporting issues in Norway, Sweden and United Kingdom. For these countries, we use cumulative confirmed cases for calculating the growth rates.

#### S2.3 Growth Rate

We use daily active cases to estimate the daily growth rate for these countries. Growth Rate (Equation S8) can be very volatile at the start of the pandemic due to the low number of cases in the early stages. For example, a unit increase in $I_{jt}$ will record a growth rate of 0.4 when $I_{jt} = 2$ as compared to a growth rate of 0.0004 when $I_{jt} = 1000$ (growth rate is calculated as the first difference in log of active cases in consecutive days). Similarly, during the later stages of the pandemic (at least when the first wave is slowed down for some countries), growth rate could be affected by multiple other factors such as awareness or changed individual behavior. To avoid these issues, we use the data for first 60 days for a country (after we start collecting data for a country following the '$th$').

Unlike Hsiang et al. [1], we use data for 60 days and do not restrict to the initial phase when the cases rise exponentially. In §Robustness check, we discuss the performance of the model (and changes in model parameter estimates) as we add more/less data in the model from 24 countries. To filter out the volatile growth rate during the start of the pandemic, we consider data for each country when the daily new cases cross a threshold $th$. We define this threshold, as the day when the 7 day average of daily new cases in a country crosses $th$ =20% of the peak case observed in that country (till July 8, 2020). We select $th$ based on maximum likelihood estimate of the growth rate model (mechanism for the selection of $th$ is discussed later in the Section on §Robustness Check). Figure S6 shows the daily new cases. Figure S7 shows the respective growth rate. Since we use data for a maximum of 60 days for a country, from the day its daily cases cross the threshold. Thus, our dataset contains an unbalanced panel data from 24 countries.

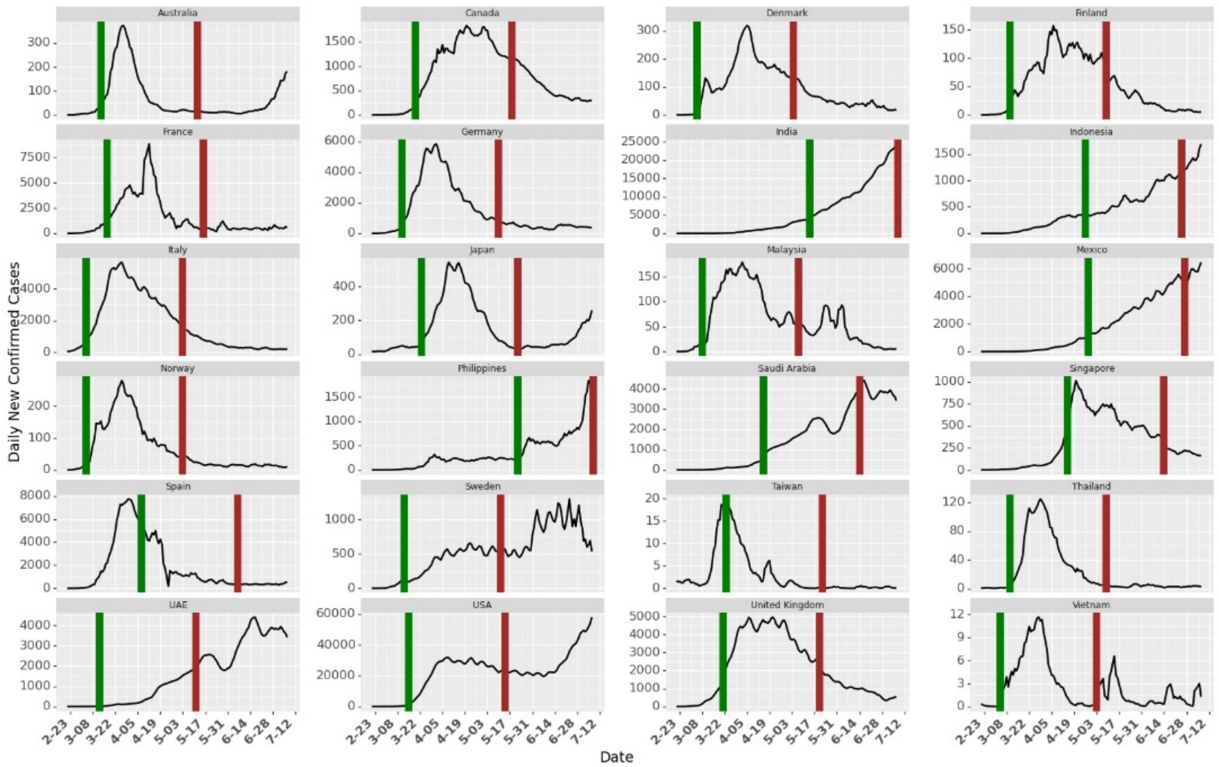


Figure S6. Daily New Cases. The green line marks the day when daily new cases in that country crossed the threshold. Brown line shows the end of 60 days of data collected for each country. For countries where cases are still increasing vis-à-vis India and Philippines, we collected fewer data points than 60 days.

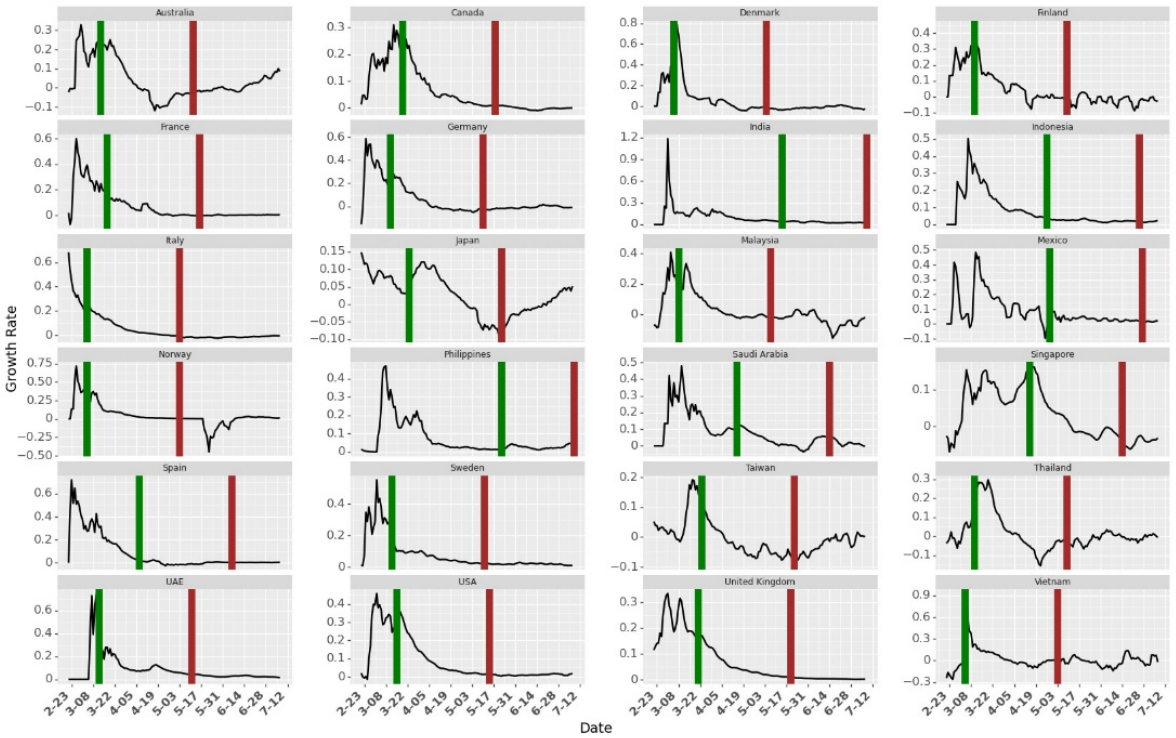


Figure S7. Growth rate across Countries. The green line marks the day when daily new cases in that country crossed the threshold. Brown line shows the end of 60 days of data collected for each country. We use cumulative confirmed cases to calculate growth rate for Norway, Sweden and United Kingdom. Collecting data after the green line allows us to filter initial noisy growth rate from that country.

### S2.4 Community Mobility

Google's COVID-19 Community Mobility Reports [7] provides information on how movement trends change over time across different types of locations in different countries. The mobility numbers are calculated based on the change in trend from the baseline (details in the report on how Google calculates the baseline). The report tracks movement trends over time by geography, across different categories of places such as retail and recreation, groceries and pharmacies, parks, transit stations, workplaces, and residential. Figure S8 shows the community mobility across different countries.

We observe high correlation between the social mobility numbers from Google across different types of locations. This may lead unstable parameter estimates due to multicollinearity in parameter estimation using ordinary least squares. Based on the correlations, we consider the mobility in Parks and Transit stations as our measure for mobility. We also confirm these two categories using a Lasso regression (more details in Section §Robustness Check in Section 4.5.3). The Lasso regression model pushes the coefficients of correlated variables (variable which do not add much information to the model) to 0 and gives non-zero weights to only two of the mobilities: Parks and Transit stations.

The correlation matrix between the mobility across different locations is shown in Table S1. The correlation matrix shows that transit stations is highly correlated with mobility in Retail and Recreation, Grocery and Pharmacy and Residential. Mobility in transit stations is negatively correlated with mobility in Residential as – fewer people travel indicates establishes that more people are staying home. Thus, mobility in transit stations is able to capture the information from mobility across all other locations except Parks. Henceforth, we include mobility in Parks and Transit stations as a measure of mobility (and as also selected by Lasso Regression model). In our analysis, we normalize the number for social mobility such that $0 \leq mobility_{j,t,m} \leq 1\ \forall j, \forall t, \forall M$.

Table S1. Correlation Matrix for Community Mobility in Different Locations

| | **Retail and Recreation** | **Grocery and Pharmacy** | **Parks** | **Transit Stations** | **Workplace** | **Residential** | **Driving** | **Walking** |
|---|---|---|---|---|---|---|---|---|
| **Retail and Recreation** | 1.00 | 0.85 | **0.53** | **0.90** | 0.75 | -0.85 | 0.81 | 0.82 |
| **Grocery and Pharmacy** | 0.85 | 1.00 | **0.45** | **0.78** | 0.67 | -0.74 | 0.69 | 0.67 |
| **Parks** | **0.53** | **0.45** | **1.00** | **0.42** | **0.16** | **-0.51** | **0.73** | **0.67** |
| **Transit Stations** | **0.90** | **0.78** | **0.42** | **1.00** | **0.84** | **-0.89** | **0.73** | **0.78** |
| **Workplace** | 0.75 | 0.67 | **0.16** | **0.84** | 1.00 | -0.87 | 0.53 | 0.58 |
| **Residential** | -0.85 | -0.74 | **-0.51** | **-0.89** | -0.87 | 1.00 | -0.74 | -0.75 |
| **Driving** | 0.81 | 0.69 | **0.73** | **0.73** | 0.53 | -0.74 | 1.00 | 0.91 |
| **Walking** | 0.82 | 0.67 | **0.67** | **0.78** | 0.58 | -0.75 | 0.91 | 1.00 |

Summary statistics for the community mobility is shown in Table S2.

Table S2. Summary Statistics on Mobility Trends from Google

| Mobility | Min | Mean | Max | Std Dev | 25th percentile | 75th percentile |
|---|---|---|---|---|---|---|
| **Retail and Recreation** | -96 % | -32.3 % | 23 % | 27.1 % | -53 % | -10 % |
| **Grocery and Pharmacy** | -94 % | -10.1 % | 51 % | 18.7 % | -19 % | 2 % |
| **Parks** | -91 % | 4.7 % | 517 % | 65.5 % | -38 % | 24 % |
| **Transit Stations** | -92 % | -38.8 % | 14 % | 23.9 % | -57 % | -20 % |
| **Workplaces** | -90 % | -27.7 % | 57 % | 23.8 % | -45 % | -7 % |
| **Residential** | -13 % | 12.8 % | 55 % | 10.4 % | 4 % | 19 % |

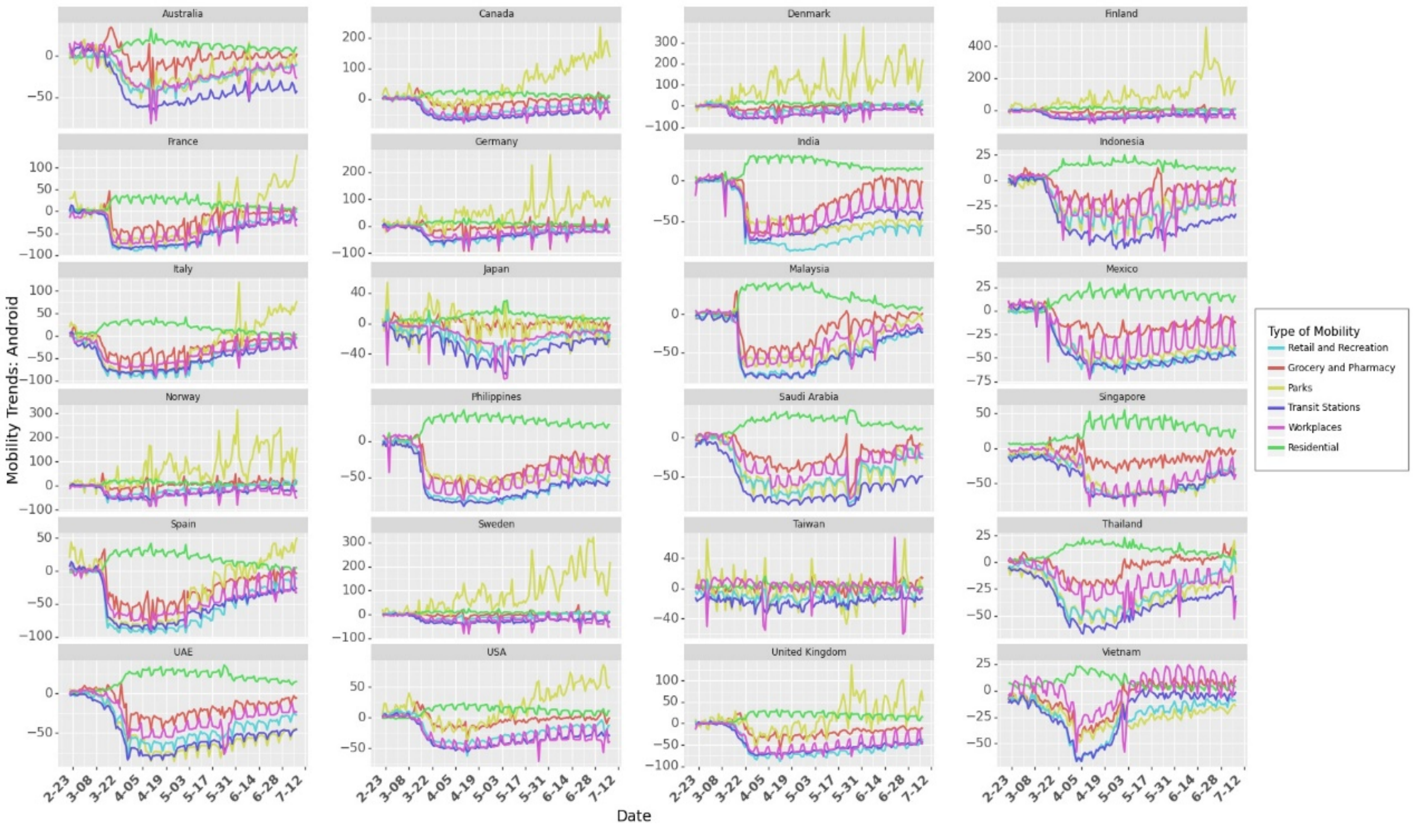


Figure S8. Community Mobility Trend from Google using Android Operating System.

Google community mobility reports uses data from users with android operating system. Also, they collect data from users who allow location sharing (Google does not disclose any personal information in Community Reports). To check the robustness of the model and our estimates on the effect of mobility, we also use mobility data from Apple Mobility Trend reports [8] to validate the results from Google Community Mobility reports. The Apple Mobility reports show a relative volume of directions requests per country/region, sub-region or city compared to a baseline volume on January 13th, 2020. Apple compares the relative volume for Driving, Transit and Walking in their dataset. However, we could not find the data on Transit for all the 24 countries considered in this paper. So we use timeline for Driving and Walking as a proxy for measuring social mobility. Table S3 provides the summary statistics on Apple Mobility Trends. The mobility trends for Driving and Walking across 24 countries is shown in Figure S9.

Table S3. Summary Statistics on Mobility Trends from Apple

| Mobility | Min | Mean | Max | Std Dev | 25th percentile | 75th percentile |
|---|---|---|---|---|---|---|
| **Driving** | -100 % | -28.2 % | 184 .9% | 48.5 % | -64.9 % | -6.2% |
| **Walking** | -100 % | -35.7 % | 94.4 % | 44.1 % | -72 % | 0.58 % |

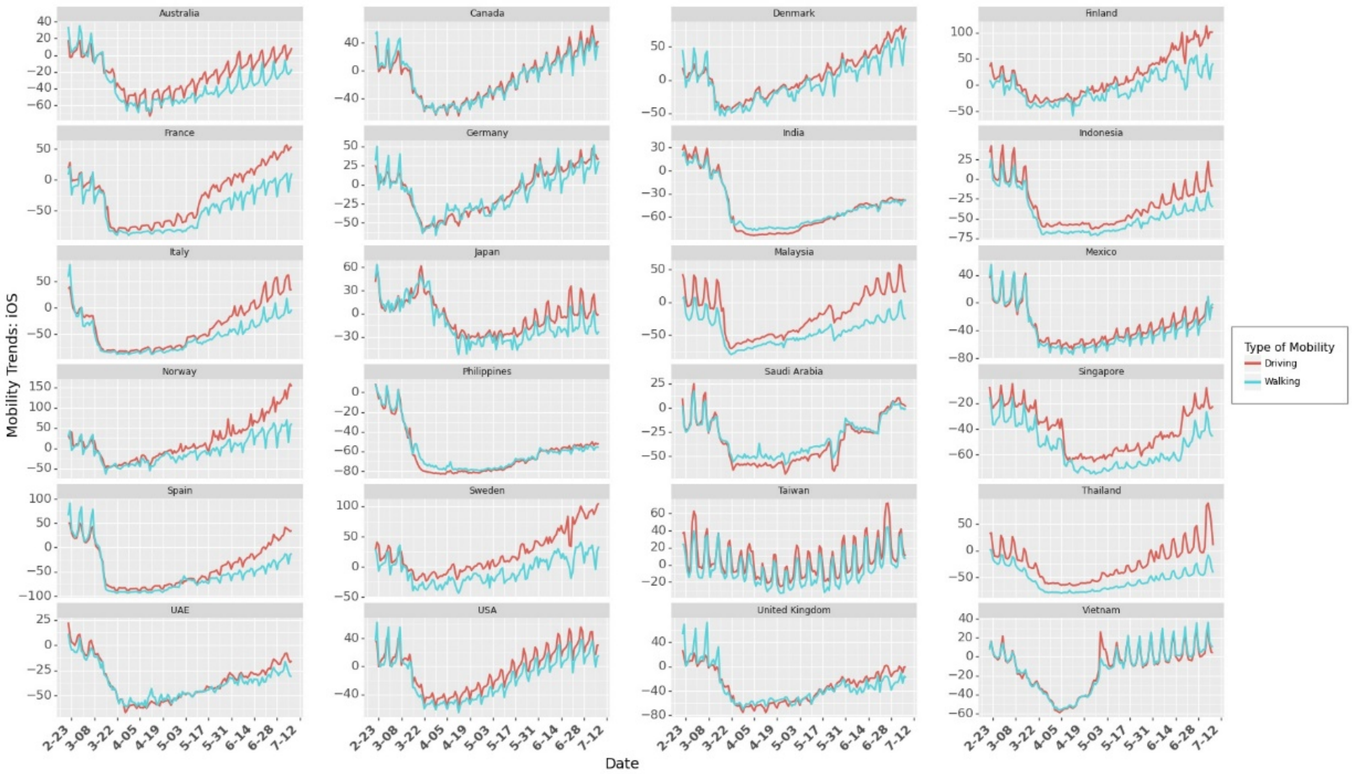


Figure S9. Community Mobility Trend from Apple using iOS

### S2.5 Non-Pharmaceutical Interventions (NPIs)

Governments (and its policies) play a critical role in fighting a pandemic. Vaccines may take a long time to be available, particularly for a new disease e.g. COVID-19. In an ongoing pandemic, we cannot depend only on the vaccines but need strong government interventions (institutional measures) to control the spread of the disease. During such times, governments must take various measures e.g. increasing testing infrastructure to control the spread of infections. These NPIs help in decreasing mobility (for example travel bans imposed restriction on travel across states/ regions/ countries while it also helped in restricting mass gatherings in places such as transit stations. Since COVID-19 spreads through person to person physical interaction (or prolonged proximity), governments introduced various policies (Non-Pharmaceutical Interventions or NPIs) for social distancing to minimize person-to-person interaction. They also introduced closures of places where people gather together at the same time e.g. schools or businesses. However, these government policies seriously affected businesses [9], leading to economic shutdowns which adversely affecting the poor community [10]. The effect of these shutdowns may also lead to prolonged economic hardships e.g. closure of some businesses and employment [11].

Since these policies directly affect the livelihood of a majority of the population across the world, it is important to investigate the impact they have on controlling the spread of the disease. As these policies were implemented at different times across different countries, it gives us an opportunity to explore the combined effect of these policies. Estimating the combined effect of these policies could help the governments in future (or current) pandemics to introduce policies that are effective and may not necessarily lead to complete lockdown unless extremely necessary. Please note that some of these policies were introduced at the same time, or some of the policies were implemented first and some of the policies were implemented always after some other policy were implemented, we do not claim any causal effect of the policy on the growth rate. It is difficult to isolate the effect of individual policies as the implementation of policies were not randomly sequenced across countries.

We use Coronanet dataset from Cheng at al [12]. They collected information on all the government policies introduced by different countries across the world. They categorized the policies into 19 different $policy\_types$. We use their categorization to build our model. The policies were implemented at different levels – National, Provincial and Municipal. In this work, we consider the policies implemented at National and Provincial level. From February 21, 2020 to July 8, 2020, we check if a policy $p$ was implemented in a country $j$ or not on day $t$. If the policy was implemented, we assign a value of 1 to $s_{j,t,p}$. If the policy was introduced at a provincial level (could be introduced by the central government or a respective state government), we increase $s_{j,t,p}$ by the population of the state. Equation S13 explains $s_{jtp}$ if a policy $p$ is employed at a provincial level where $\widehat{N_{js}}$ is the population of state $s$ in country $j$. After identifying $s_{j,t,p}$for all countries over the period of our analysis (considering all the entries in the dataset), we use normalization using maximum value in a country such that $s_{j,t,p} \in [0,1]\ \forall j,\ \forall t,\ \forall p$ as shown in Equation S13..

$$s_{j,t,p} = s_{j,t,p} + \frac{\widehat{N_{js}}}{\widehat{N_{js}} + N_j} \qquad \text{(S13)}$$

$$s_{j,t,p} = \frac{s_{j,t,p}}{\max_{t,p} s_{j,t,p}} \qquad \text{(S14)}$$

The dataset contains 5816 entries on policies (some of the policies were announcements/ recommendations/ new entry or an update to existing policy) at National and Provincial level. The dataset provides detailed information on the type of the data entry (e.g. policy type, description of the policy). The statistics on the types of policies is shown in Figure S10. Figure S10 also provides a count of entries of each policy type. The data set contains 20 $policy\_types$. Figure S10 shows how many countries implemented (light grey bars) a particular $policy\_type$. It also shows how many countries implemented a particular $policy\_type$ at national or provincial level (light blue bars and dark blue bars respectively). The dark grey bars show the total number of entries (divided by 100 for visualization) for all $policy\_types$.

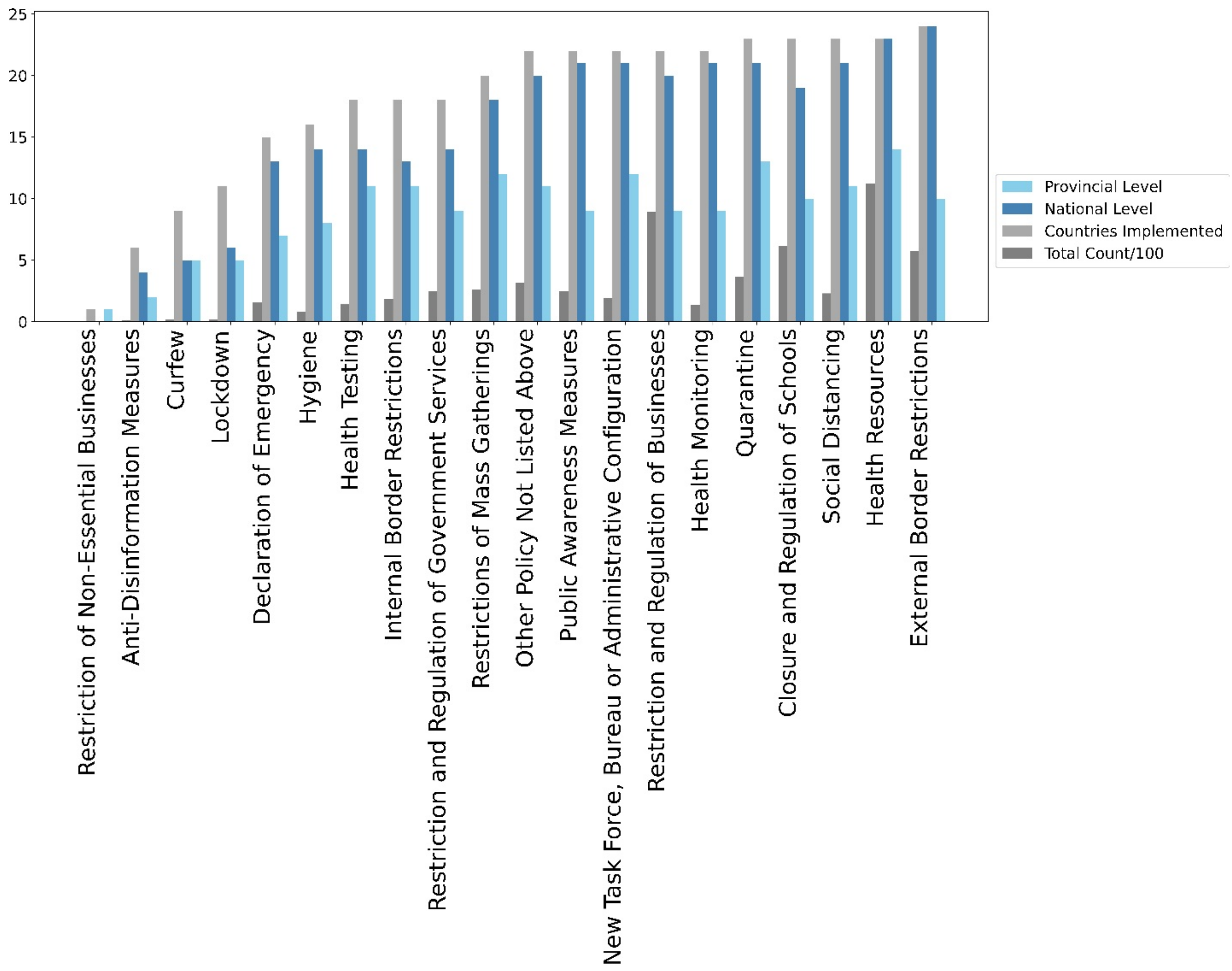


Figure S10. Policy Implementation at National and Provincial Levels Across Different Countries

We use the text description of the policy to identify if an entry was an update, recommendation or actual implementation. If the entry was an announcement or an update for a policy with start date and an end date, we give a weightage of 0 to that entry in the dataset because if there is policy update, it could recount that policy. Note that the policies could have been implemented differently across different countries, even if they were categorized in the same policy type. For example, a country may impose Social distancing rules from 4 pm – 8 pm while another country may impose it from 6 am – 6 pm. It may differ across different states in country. However, for the purpose of this research, we do not consider the variations in implementations of policies.

As we have survey numbers for wearing face masks at a national level, we a consider policy type that were implemented across majority of the countries. Therefore, we do not consider Anti-disinformation measures, Curfew and Lockdowns. Curfew and Lockdowns are similar to Quarantine and Restrictions of Mass Gatherings (which lead to closure of places of mass gatherings) so we can ignore them for the purpose of this research. Moreover, Curfew and Lockdowns affect the community mobility, which can be accounted for by trend in social mobility from Google Community Mobility Reports (discussed in the previous Section).

We also do not consider Hygiene Announcements and New Task Force policy as these were administrative announcements and did not have much effect on the growth rate of the infection. Some policies did not have a start date and end date. We calculate the cumulative number of times announcements were made for such categories. Health Testing, Health Monitoring and Health Resources are administrative announcements, so we combine them in to one Health Resources policy. As we use linear models, a linear combination (addition of three policies) does not affect our analysis. It further reduces the number of parameters to be estimated. Similarly, we combined Restrictions and Regulations of Businesses and Restriction and Regulation of Government Services. Health resources can also be used as proxy for increased awareness among governments and citizens. So, we do not consider "Public Awareness Measures" announcements to avoid multicollinearity in the set of predictor variables.

Table S4 shows the correlation between different government policies implemented across countries. The correlation value between pairs of any two NPIs is not high (>0.7 as observed with community mobility across different locations in Google Community Mobility Reports), so we include all the following 8 government policies in our model in Equation S9. We also include the Social Mobility in Parks and Transit Stations to check its correlation with NPIs. Changes (reduction during the early stages of pandemic) in social mobility were induced by the introduction of NPIs. However, social mobility is a combination of institutional measures e.g. NPIs and individual measures e.g. social mobility. Correlation between social mobility and any NPIs as shown in Table S4 is not high (>0.7) so we do not reject any further NPIs.

Table S4. Correlation Between NPIs

| | Health Resources | Restriction and Regulation | Closure and Regulation of Schools | External Border Restrictions | Quarantine | Restrictions of Mass Gatherings | Social Distancing | Internal Border Restrictions | Mobility Parks | Mobility Transit Stations |
|---|---|---|---|---|---|---|---|---|---|---|
| Health Resources | 1.00 | 0.32 | 0.04 | 0.56 | 0.34 | 0.26 | 0.35 | 0.49 | -0.26 | -0.26 |
| Restriction and Regulation of Businesses | 0.32 | 1.00 | 0.55 | 0.46 | 0.35 | 0.58 | 0.44 | 0.43 | -0.01 | -0.44 |
| Closure and Regulation of Schools | 0.04 | 0.55 | 1.00 | 0.16 | 0.25 | 0.52 | 0.15 | 0.40 | 0.02 | -0.40 |
| External Border Restrictions | 0.56 | 0.46 | 0.16 | 1.00 | 0.28 | 0.38 | 0.47 | 0.26 | -0.19 | -0.54 |
| Quarantine | 0.34 | 0.35 | 0.25 | 0.28 | 1.00 | 0.34 | 0.32 | 0.26 | 0.14 | 0.02 |
| Restrictions of Mass Gatherings | 0.26 | 0.58 | 0.52 | 0.38 | 0.34 | 1.00 | 0.35 | 0.37 | -0.01 | -0.38 |
| Social Distancing | 0.35 | 0.44 | 0.15 | 0.47 | 0.32 | 0.35 | 1.00 | 0.24 | -0.14 | -0.41 |
| Internal Border Restrictions | 0.49 | 0.43 | 0.40 | 0.26 | 0.26 | 0.37 | 0.24 | 1.00 | -0.21 | -0.26 |
| Mobility Parks | -0.26 | -0.01 | 0.02 | -0.19 | 0.14 | -0.01 | -0.14 | -0.21 | 1.00 | 0.51 |
| Mobility Transit Stations | -0.26 | -0.44 | -0.40 | -0.54 | 0.02 | -0.38 | -0.41 | -0.26 | 0.51 | 1.00 |

Policy implementation across countries for policies considered in this work (as described above) is shown in Figure S11 and implementation of policies across different countries is shown in Figure S12. Note that we normalize $s_{j,t,p}$ such that $s_{j,t,p} \in [0,1]\ \forall j,\ \forall t,\ \forall p$.

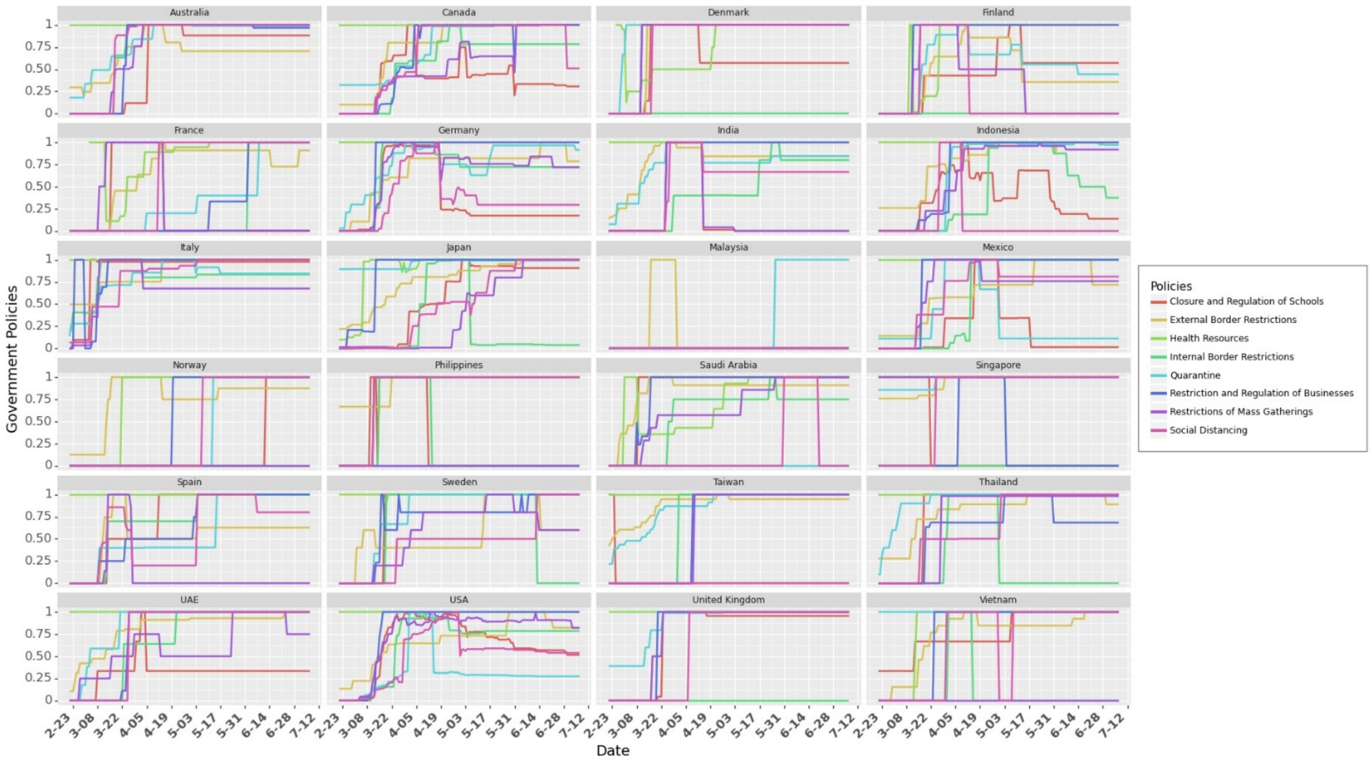


Figure S11. Policy Implementation Across Different Countries. This graph shows how some countries e.g. Taiwan and Malaysia employed only few of the policies. It also shows that countries e.g. USA and Mexico employed region specific policies (at provincial level).

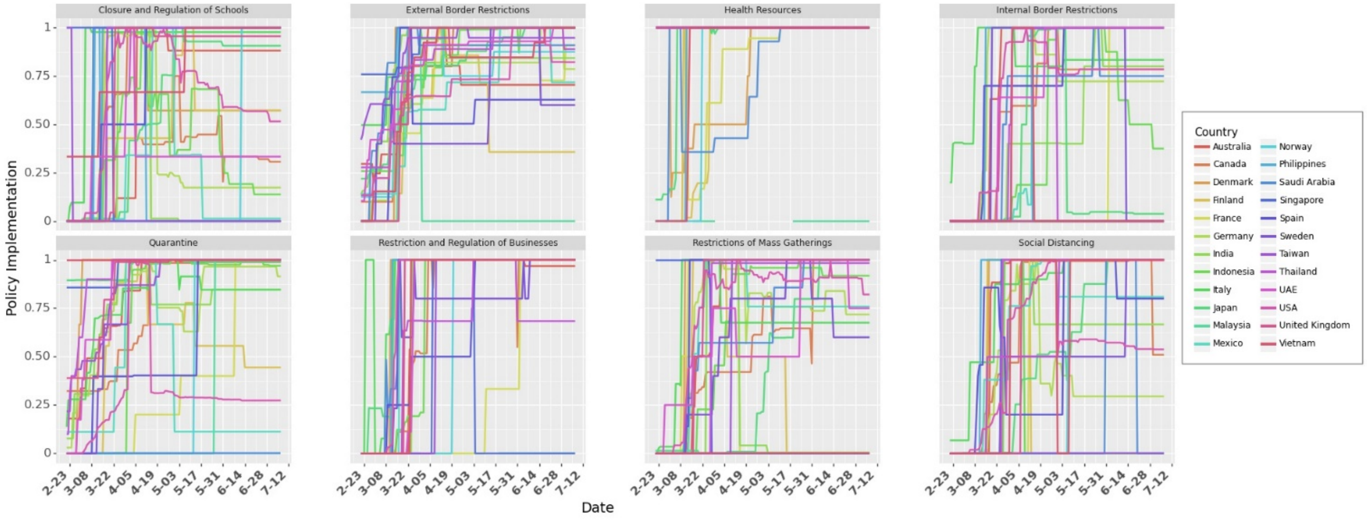


Figure S12. Country wise Implementation of Different Policies. Figure shows how majority of the government policies were introduced at similar times across different countries. This makes it difficult to estimate the causal impact of each NPI on the spread of COVID-19.

### S2.6 Lag in Observation of the Effects of Control Variables

Studies have reported a delay in observation of effects of policies on the events on a given day. This delay could be due to several reasons. One of the most prominent reasons is the incubation period (time between getting infected and onset of symptoms/knowing that individual is confirmed for COVID-19). During the incubation period, an individual may be asymptomatic. Incubation period is estimated to be 4 days to 14 days [2]. Another reason could be the testing time – time it takes to get the confirmation of results. Due to limited healthcare professionals, there could be a long queue to get tested to get the results from testing centers.

To model this delay, we use as lag variable, $shift$. We use cross validation method to find the lag with the best fit for the data. We test $shift \in [0,14]$ and observe that the model performs best at a *shift* of 9 days. We discuss this further in Section §Robustness check.

**S2.7 Testing**

Testing is critical in identifying the infectious individuals. Once identified, these individuals can be quarantined or isolated from public so that they do not spread to susceptible individuals. While people can get tested when they start showing symptoms, evidence reports that even asymptomatic individuals can spread the virus (50% of cases can be attributed to asymptomatic cases [13]) Since they do not show any symptoms, people around them (e.g. asymptomatic young adult living with family) are less cautious and may get infected through them. It is critical to identify asymptomatic individuals as they can spread the virus unknowingly. This can be done by increased testing and contact tracing the individual who have come in contact with those who tested positive for the COVID-19. Testing can be crucial in identifying COVID-19 positive individuals so that they can be quarantined (hospital or home isolation) or treated early when symptoms starts showing. In our analysis, we normalize the number for social mobility such that $0 \leq testing_{j,t} \leq 1 \; \forall j, \forall t$.

As testing increases, the probability that more confirmed positive cases will be identified. This will lead to increase in empirical growth rate over time as more confirmed cases will be reported. This shows a change in testing pattern over time which could lead to bias (or underestimating the effect of NPIs and masks). To counter this time sensitive bias, we use testing data to account for increased testing over time. Figure S13 shows the data on total tests (per thousand people) in a country from ourworldindata.org [14].

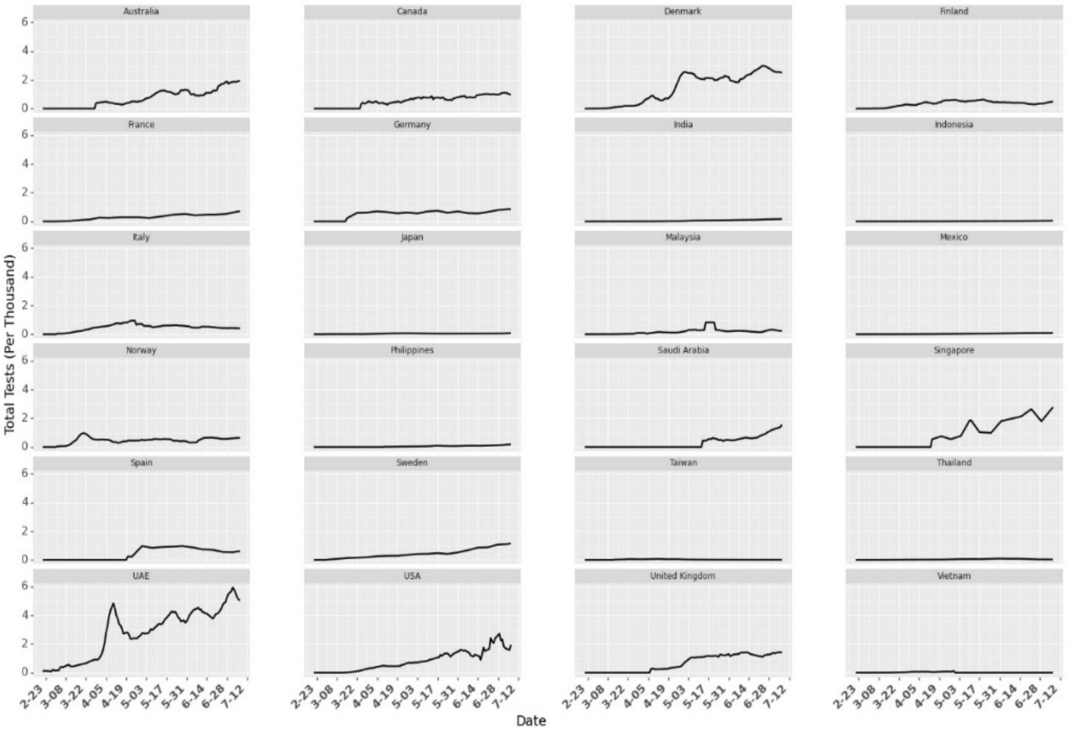


Figure S13. Tests per Thousand in different countries over time. The numbers indicate the total number of tests conducted per thousand people in a country but the data set does not provide information on how many individuals were tested for COVID-19.

### S2.8 Google Trends

As the number of cases start increasing, awareness increased in public (e.g. washing hands more often). We use Google Trends [15] to account for the increase in active awareness over time (Figure S14). Google Trends numbers indicate the search interest of a topic over time as a proportion of all other searches at the same time. In our analysis, we normalize the number for Google Trends such that $0 \leq trend_{j,t} \leq 1 \; \forall j, \forall t$.

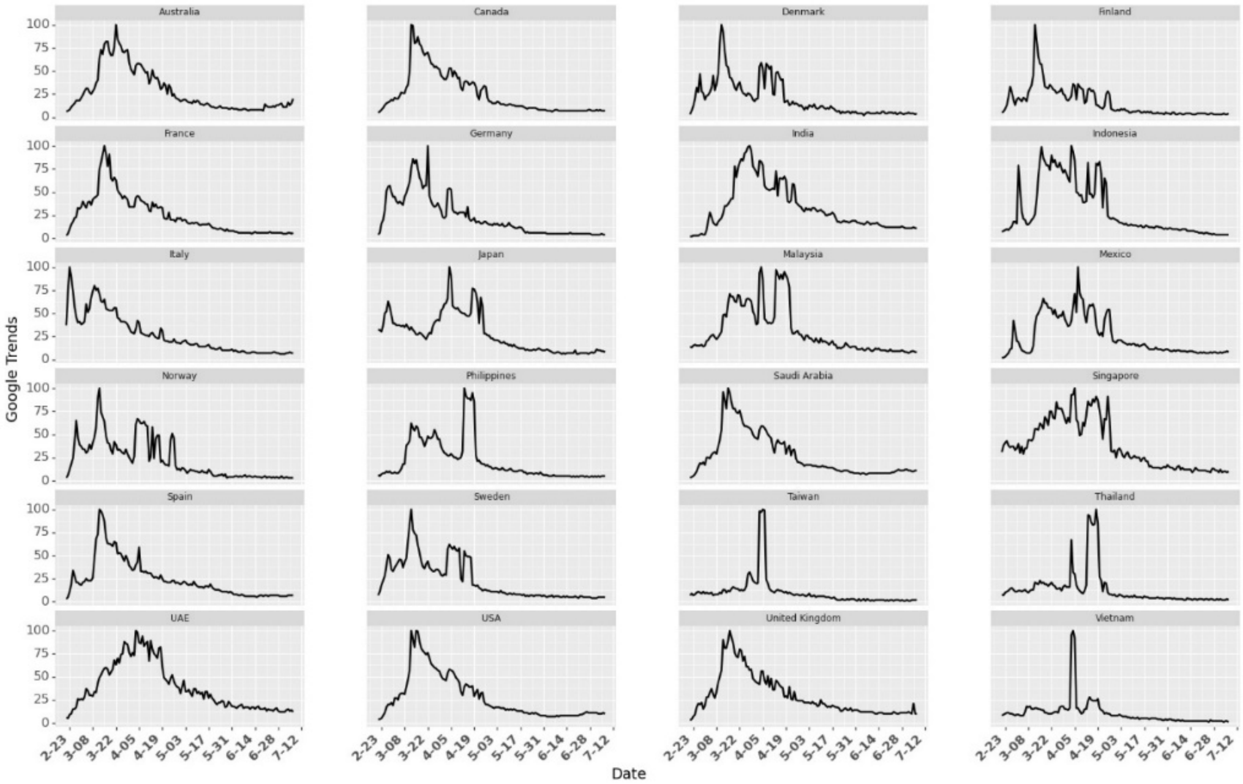


Figure S14. Google Trends for the Search term *coronavirus*

### S2.9 Week Fixed Effects

Handling of COVID-19 changes over time. It includes increase in public awareness or better understanding of the virus as more studies and research comes to public attention. Not only do citizens understand how to be more careful (or more informed), healthcare providers also learn more about the disease for more efficient treatment of COVID-19 patients (e.g. creating new wards for COVID-19 patients, treating them by wearing Personal Protection Kits, PPE). It also involves improved infrastructure e.g. testing, converting existing medical facilities to dedicated COVID—19 centers. To account for all the time sensitive fixed effects (other than the controls we discuss before), we use fixed effects for weeks (from the day that country reaches $th$ in our analysis).

Figure S14 shows how the growth rate changes across different weeks (Figure S15 provides same information across different countries). Even after removing the initial noisy data, we observe highest variance in the growth rates during week 1 in most countries (Figure S14 and Figure S15). We use one hot vector to denote week ($week_{j,t,w} = 1$) if day $t$ lies in week $t$ for country $j$. Note that due to different starting times for each country (Figure S6), a day may come under different week for different country. For example, days in week 1 for Vietnam are earlier in the calendar as compared to the days in week 1 in

India. Note that the growth rate is higher during the initial weeks and slows down with time. To capture this effect, we use fixed effects for weeks. Results in Section §Result show that the magnitude of coefficient for week 0 is higher than the magnitude of the coefficient for week 1 and so on.

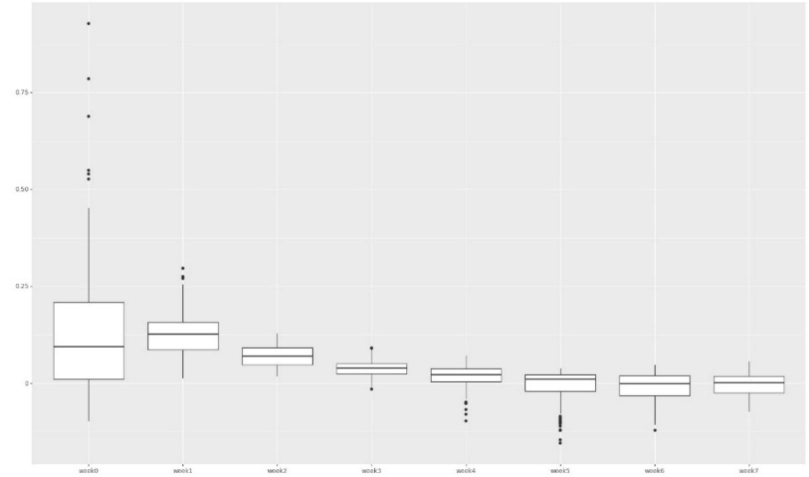

Figure S14. Box plot for growth rate in different weeks across different countries

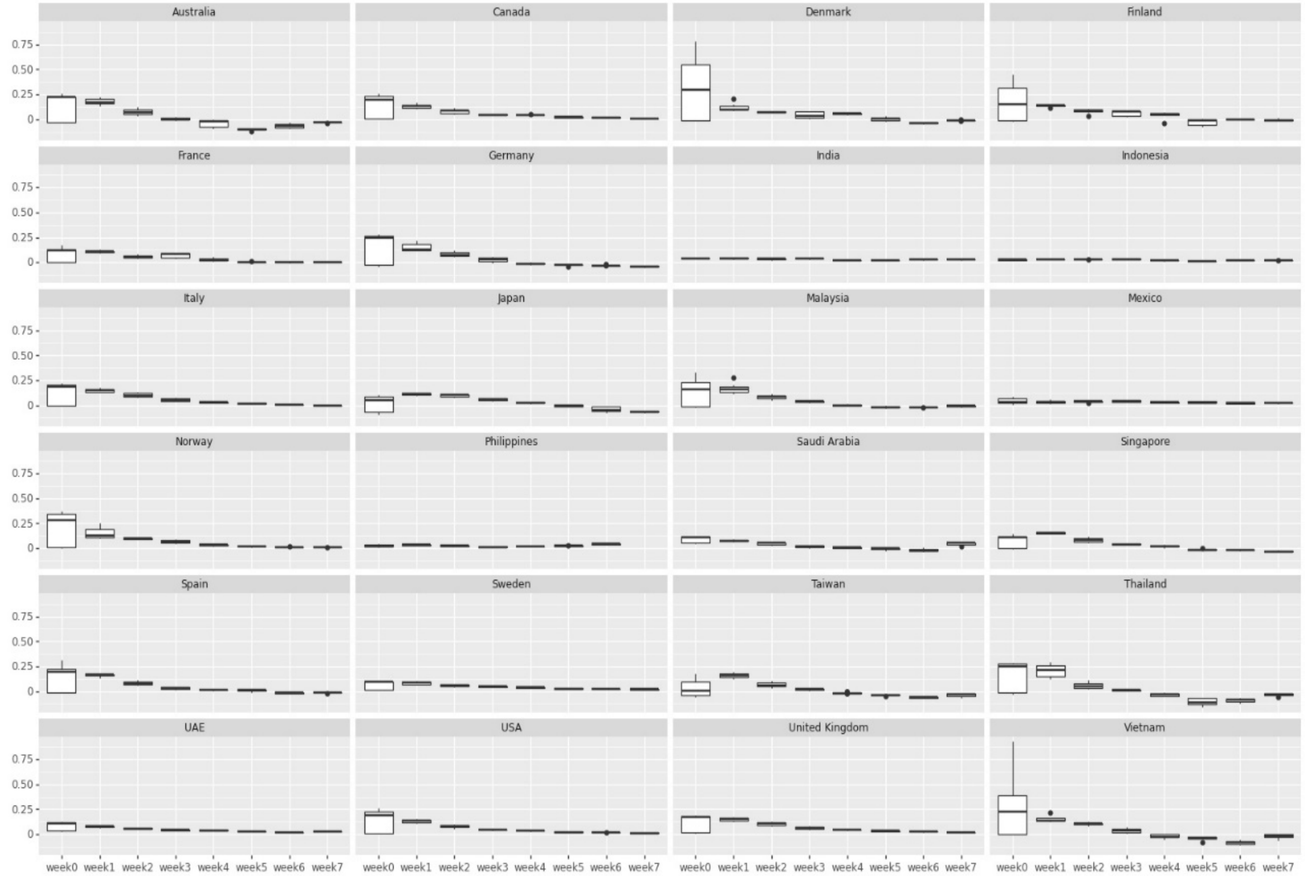


Figure S15. Box plot for growth rate in different weeks across different countries

#### S2.10 Country Fixed Effects

We observe heterogeneity across countries with many aspects in handling COVID-19. Multiple factors affect the spread and handling of a disease in a country. Heterogeneity may be observed at different levels. For example, heterogeneity at government level includes difference in reporting cases, testing infrastructure, strictness is reducing social mobility and implementation of NPIs. Population wise heterogeneity includes population density in a country or percentage of population living in high density urban regions or poor neighborhoods with shared sanitation facilities. It may also include cities with international airports or international travelers (particularly from countries hard hit with COVID-19 in early 2020 e.g. China and Iran). It also includes heterogeneity at the level of education (awareness about COVID-19, responsibility in understanding the severity of precautions), poverty (health insurance, ability to purchase sanitizers or high quality masks), basic health care facilities (drinking water, sanitation, shared places) or family structure (number of young adults in a family or size of the family residing in a residential complex). To control for all this heterogeneity among countries which may lead to country level effects in the growth rate of COVD-19, we use country fixed effects.

Note that since we use a constant in our model, we consider fixed effects for 23 countries and we consider last country (Vietnam) as our base country (with 0 fixed country effect). This ensures that parameter estimates are stable.

### S3. Results

We use growth rate model in our analysis to study the effect of Masks, Social Mobility and Non-Pharmaceutical Interventions (NPIs). Since we do not have real numbers on how many people wear masks (or wear masks that could be effective), we use different transformations of the mask numbers from the surveys. We use growth rate model with masks transformed as $\ln(1 + mask_{jt})$ as our focal model. We use threshold $th = 0.2$ and $shift = 9$ days. Details on selection of $th$ and $shift$ is provided in Section §Robustness Check. Model statistics are given in Table S5. Parameter estimates for model with different transformations is shown in Table S6.

Table S5. Model statistics

| | |
|---|---|
| R-squared: | 0.738 |
| Adj. R-squared: | 0.729 |
| F-statistic: | 88.03 |
| Prob (F-statistic): | 0 |
| Log-Likelihood: | 2400.8 |
| AIC: | -4712 |
| BIC: | -4475 |
| No. Observations: | 1422 |
| Df Residuals: | 1377 |
| Df Model: | 44 |

Table S6. Parameter Estimates for Growth Rate Model

| var | coefficient | std error | t-value | p-value | lower limit | upper limit |
|---|---|---|---|---|---|---|
| const | 0.1999 | 0.017 | 11.745 | 0 | 0.167 | 0.233 |
| log Mask | -0.1047 | 0.024 | -4.385 | 0 | -0.152 | -0.058 |
| Mobility Parks | -0.0296 | 0.006 | -4.677 | 0 | -0.042 | -0.017 |
| Mobility Transit Stations | 0.1109 | 0.013 | 8.398 | 0 | 0.085 | 0.137 |
| week0 | 0.0981 | 0.009 | 11.025 | 0 | 0.081 | 0.116 |

| | | | | | | |
|---|---|---|---|---|---|---|
| week1 | 0.0589 | 0.009 | 6.872 | 0 | 0.042 | 0.076 |
| week2 | 0.0411 | 0.008 | 5.146 | 0 | 0.025 | 0.057 |
| week3 | 0.0324 | 0.007 | 4.335 | 0 | 0.018 | 0.047 |
| week4 | 0.018 | 0.007 | 2.575 | 0.01 | 0.004 | 0.032 |
| week5 | 0.0039 | 0.007 | 0.592 | 0.554 | -0.009 | 0.017 |
| week6 | -0.0013 | 0.006 | -0.209 | 0.834 | -0.014 | 0.011 |
| week7 | 0.0021 | 0.006 | 0.343 | 0.732 | -0.01 | 0.014 |
| Testing | -0.0121 | 0.006 | -1.938 | 0.053 | -0.024 | 0 |
| Trend | -0.0455 | 0.008 | -5.933 | 0 | -0.061 | -0.03 |
| Health Resources | -0.034 | 0.012 | -2.881 | 0.004 | -0.057 | -0.011 |
| Restriction and Regulation of Businesses | -0.0049 | 0.005 | -1.03 | 0.303 | -0.014 | 0.004 |
| Closure and Regulation of Schools | -0.0153 | 0.006 | -2.436 | 0.015 | -0.028 | -0.003 |
| External Border Restrictions | -0.0315 | 0.008 | -3.807 | 0 | -0.048 | -0.015 |
| Quarantine | -0.0321 | 0.01 | -3.194 | 0.001 | -0.052 | -0.012 |
| Restrictions of Mass Gatherings | -0.0066 | 0.007 | -1.007 | 0.314 | -0.019 | 0.006 |
| Social Distancing | 0.0038 | 0.006 | 0.618 | 0.536 | -0.008 | 0.016 |
| Internal Border Restrictions | -0.01 | 0.006 | -1.639 | 0.101 | -0.022 | 0.002 |
| Australia | -0.0338 | 0.013 | -2.531 | 0.011 | -0.06 | -0.008 |
| Canada | 0.0205 | 0.012 | 1.645 | 0.1 | -0.004 | 0.045 |
| Denmark | 0.0167 | 0.014 | 1.23 | 0.219 | -0.01 | 0.043 |
| Finland | -0.02 | 0.016 | -1.27 | 0.204 | -0.051 | 0.011 |
| France | -0.032 | 0.014 | -2.209 | 0.027 | -0.06 | -0.004 |
| Germany | -0.0101 | 0.014 | -0.739 | 0.46 | -0.037 | 0.017 |
| India | -0.0038 | 0.013 | -0.295 | 0.768 | -0.029 | 0.022 |
| Indonesia | 0.0353 | 0.013 | 2.71 | 0.007 | 0.01 | 0.061 |
| Italy | 0.0481 | 0.011 | 4.308 | 0 | 0.026 | 0.07 |
| Japan | -0.0034 | 0.01 | -0.348 | 0.728 | -0.022 | 0.016 |
| Malaysia | -0.0397 | 0.013 | -2.944 | 0.003 | -0.066 | -0.013 |
| Mexico | 0.0002 | 0.017 | 0.014 | 0.989 | -0.032 | 0.033 |
| Norway | -0.0373 | 0.016 | -2.284 | 0.023 | -0.069 | -0.005 |
| Philippines | -0.0315 | 0.017 | -1.88 | 0.06 | -0.064 | 0.001 |
| UAE | 0.1119 | 0.025 | 4.418 | 0 | 0.062 | 0.162 |
| Saudi Arabia | 0.015 | 0.016 | 0.949 | 0.343 | -0.016 | 0.046 |
| Singapore | 0.0461 | 0.015 | 3.124 | 0.002 | 0.017 | 0.075 |
| Spain | 0.0073 | 0.014 | 0.543 | 0.588 | -0.019 | 0.034 |
| Sweden | -0.0248 | 0.014 | -1.776 | 0.076 | -0.052 | 0.003 |
| Taiwan | -0.0294 | 0.013 | -2.322 | 0.02 | -0.054 | -0.005 |
| Thailand | 0.0209 | 0.012 | 1.81 | 0.071 | -0.002 | 0.044 |
| United Kingdom | 0.0215 | 0.014 | 1.547 | 0.122 | -0.006 | 0.049 |
| USA | 0.0252 | 0.013 | 1.872 | 0.061 | -0.001 | 0.052 |

Figure S16 shows the complete results for the parameter estimation under different transformations for masks. Results for parameter estimation in Table S6 show consistency in the estimates for social mobility and NPIs. They also show consistency on the parameter estimates for fixed effects (week and countries). Fixed effect of week is able to capture the trend in awareness or infrastructure change over time. In the beginning of the pandemic in a country, the growth rates were higher. This is corroborated with positive and statistically significant values for the fixed effects of weeks (decreases as week increase). The coefficients for masks seem different across the different transformations but due to its transformation, it has to be interpreted differently (as we discuss next in the interpretation of results). However, we cannot claim causality from these results as the NPIs were not randomly introduced in different countries. Nonetheless, we can estimate the combined effect of different mobilities and NPIs.

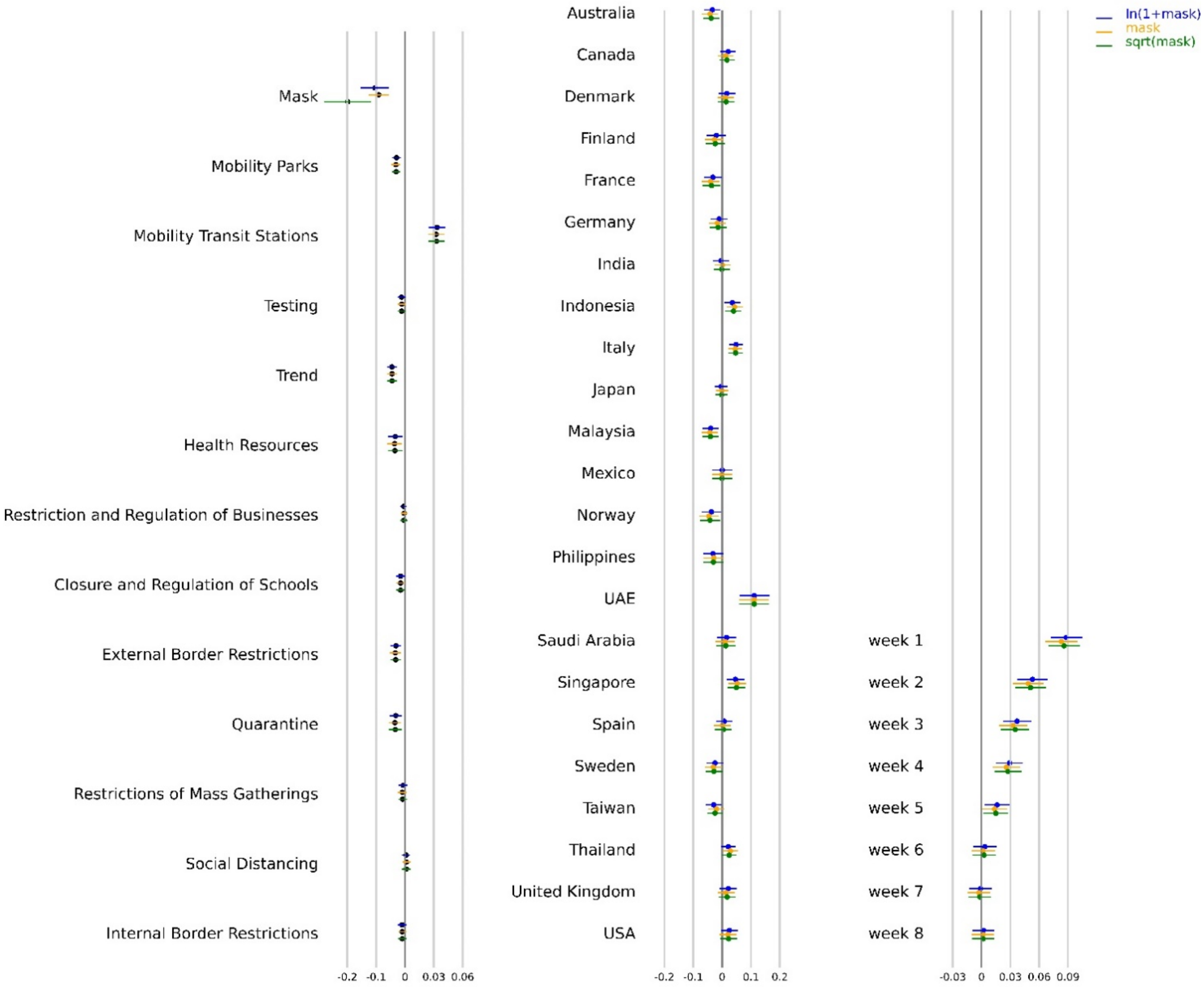


Figure S16. Parameter estimates for models with different transformations for masks. We use $th = 0.2$ and $shift$= 9 days.

### S3.1 Krinsky-Robb method

In the rest of our analysis, we use Krinsky-Robb method to estimate confidence intervals for combined effect of masks, social mobility and NPIs. We also use this method to obtain confidence bounds across the predictions of growth rate and active cases in a country. Krinsky-Robb method is a Monte Carlo simulation method used to draw samples from multivariate normal distribution. We use ordinary least square method to estimate the coefficients $\theta$ in Equation S8. Ordinary least square method for multiple linear regression assumes multivariate normal distribution of $\theta$. Krinsky-Robb method takes advantage of this assumption to sample random draws for $\theta$ using Cholesky decomposition and standard normal variates. Steps in Krinsky-Robb method are:

1. Find Cholesky decomposition matrix $C$ for the covariance matrix of $\sum_{\theta}$.
2. Draw $|\theta|$ x $n$ random samples from standard normal distribution (|x| is the cardinality of $\theta$).
3. $\theta_{samples} = \hat{\theta} + \sum_{\theta} . \text{ x } |\theta| \text{ x } n$
4. Calculate confidence interval based on $\theta_{samples}$

We use this method to get confidence interval bounds for the sum of the coefficients of mobility and NPIs to get the combined effect. We also use this method to predict confidence intervals of growth rate and daily active cases under different scenarios as we discuss next. First, we discuss the model performance and then discuss the interpretation of the coefficients in Table S6.

#### S3.2 Model Performance

We can use the coefficients from our model to predict the growth rate for different countries. Figure S17 shows the actual growth rate (green dots) with predicted growth rate (blue line) with its confidence interval (blue shade). We use Krinsky-Robb method to estimate the confidence interval bounds around the prediction. Results show that the model is accurately able to predict the growth rate of daily infections across different countries. Green and Brown vertical lines indicate the 60 days period for which data was collected for that country.

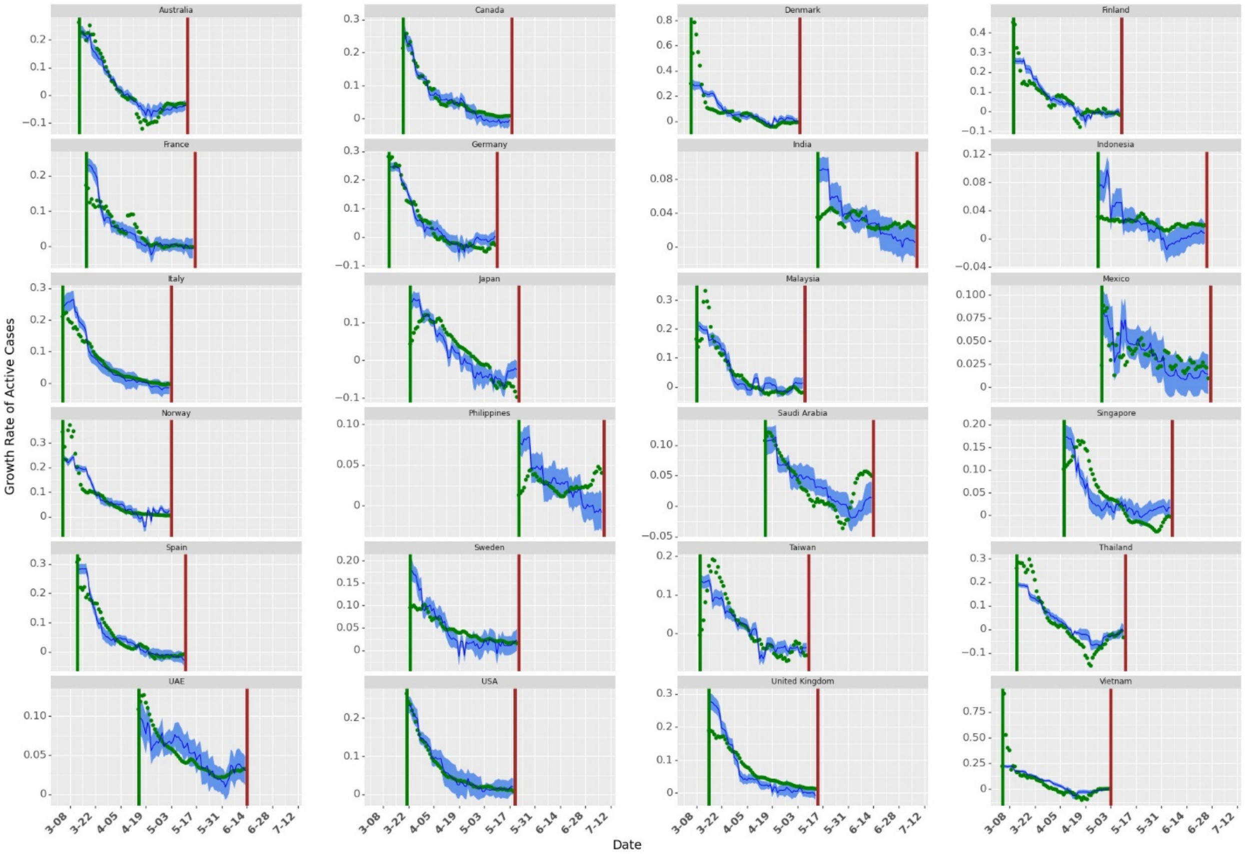

Figure S17. Growth Rate Predictions for different countries using current data. The green dots show the actual growth rates across countries. The dark blue line shows the mean of growth rate prediction (for 10000 samples in Krinksky-Robb method). The blue shaded area shows the confidence bounds around the mean prediction.

Since growth rate is a forward looking model, we can also use growth rates to estimate active infectious population by $I_{j,t} = I_{j,t-1} \mathrm{x}\, g_{j,t}$. Results for daily active cases are shown in Figure S18. Note that we estimate active cases using an exponential model. Thus, as the number of days in the prediction model increases, confidence interval bounds around predictions increase. However, the mean prediction for active cases closely approximates the actual active cases for different countries.

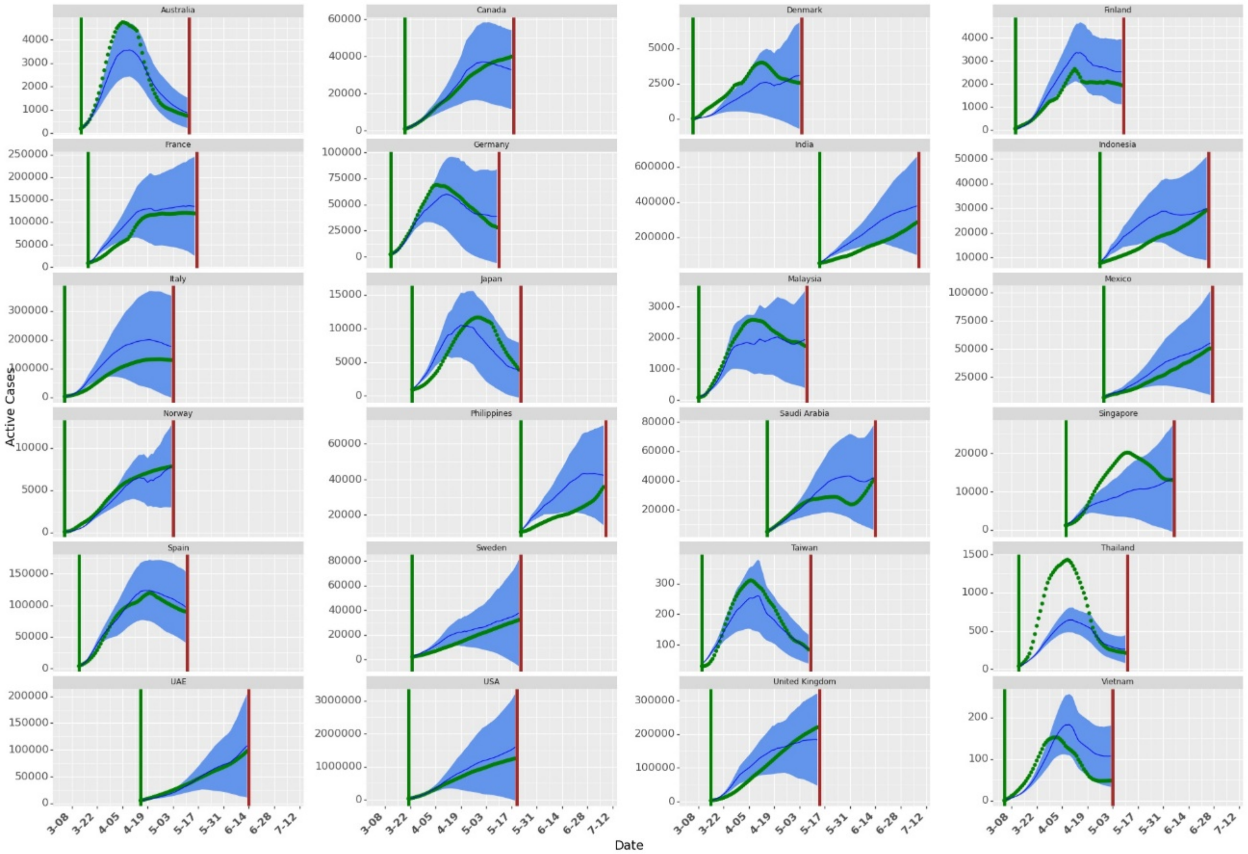


Figure S18. Simulating Daily Active Cases using Forward Looking Growth Rate Model. The green dots show the actual daily active cases across countries. The dark blue line shows the mean of daily active cases prediction (for 10000 samples in Krinksky-Robb method). The blue shaded area shows the confidence bounds around the mean prediction. As the days increase, the confidence bounds increase due to the multiplicative method in forward looking growth rate models.

#### S3.3 Effect of Masks, Social Mobility and NPIs

We model growth rate as the first difference of log of active daily infectious confirmed cases as shown in Equation S9. Thus, the exponential of the coefficients in Table S12 (other than mask as we discuss next) estimates % drop in active cases on day $t$ (as compared to active cases on day $t-1$). Using the coefficients in Table S13, we can estimate the combined effect of masks, social mobility and NPIs by using Krinsky-Robb method.

##### S3.3.1 Mask

Negative and statistically significant coefficient for masks show that increased mask wearing behavior may lead to decrease in the growth rate of COVID-19. As we use different transformations, the coefficient for masks should be interpreted differently. When masks are transformed as $\ln(1 + mask)$, a coefficient of $\theta_m$ shows that if 100% of the population wears masks, it would lead to a daily drop of $1 - e^{\theta_m(\ln(1+1)-\ln(1+0))}$ % in the growth rate as compared with the scenario when no one wears face mask. For raw mask numbers, the effect of masks can be interpreted directly as $1 - \mathrm{e}^{\theta_m}$ % drop in daily total infectious cases when everyone wears masks as compared to no one wearing masks. When masks are transformed as $\sqrt{(1 + mask)}$, coefficients should be interpreted as -- a coefficient of $\theta_m$ shows that if 100% of the population wears masks, it would lead to a daily drop of $1 - e^{\theta_m(\sqrt{(1+1)}-\sqrt{(1+0)})}$ % in the growth rate as compared with the scenario when no one wears face mask. Similarly, we can estimate the bounds for the effect of coefficients of masks. The estimate for decrease in daily growth rate when one percent additional population wears face masks in public spaces (under different transformations) is given in Table S7.

Table S7. Daily Drop in Growth Rate When Additional 100% People Wear Face Masks

| **Masks** | **Lower Limit of Daily Drop** | **Daily Drop** | **Upper Limit of Daily Drop** |
|---|---|---|---|
| $\ln(1 + mask)$ | 3.8% | 6.9% | 10.1% |
| $mask$ | 5.7% | 9.1% | 12.5% |
| $\sqrt{(1 + mask)}$ | 4.9% | 8.2% | 11.4% |

The effects of not wearing masks in each country is shown in Figure S19. Note that the results are not significantly different for Denmark, Finland, Norway and Sweden as these countries already had very low numbers for mass wearing in public spaces. Similarly, the effect is much stronger for countries e.g. Japan, Thailand and Vietnam, which have higher percentages of people wearing face masks.

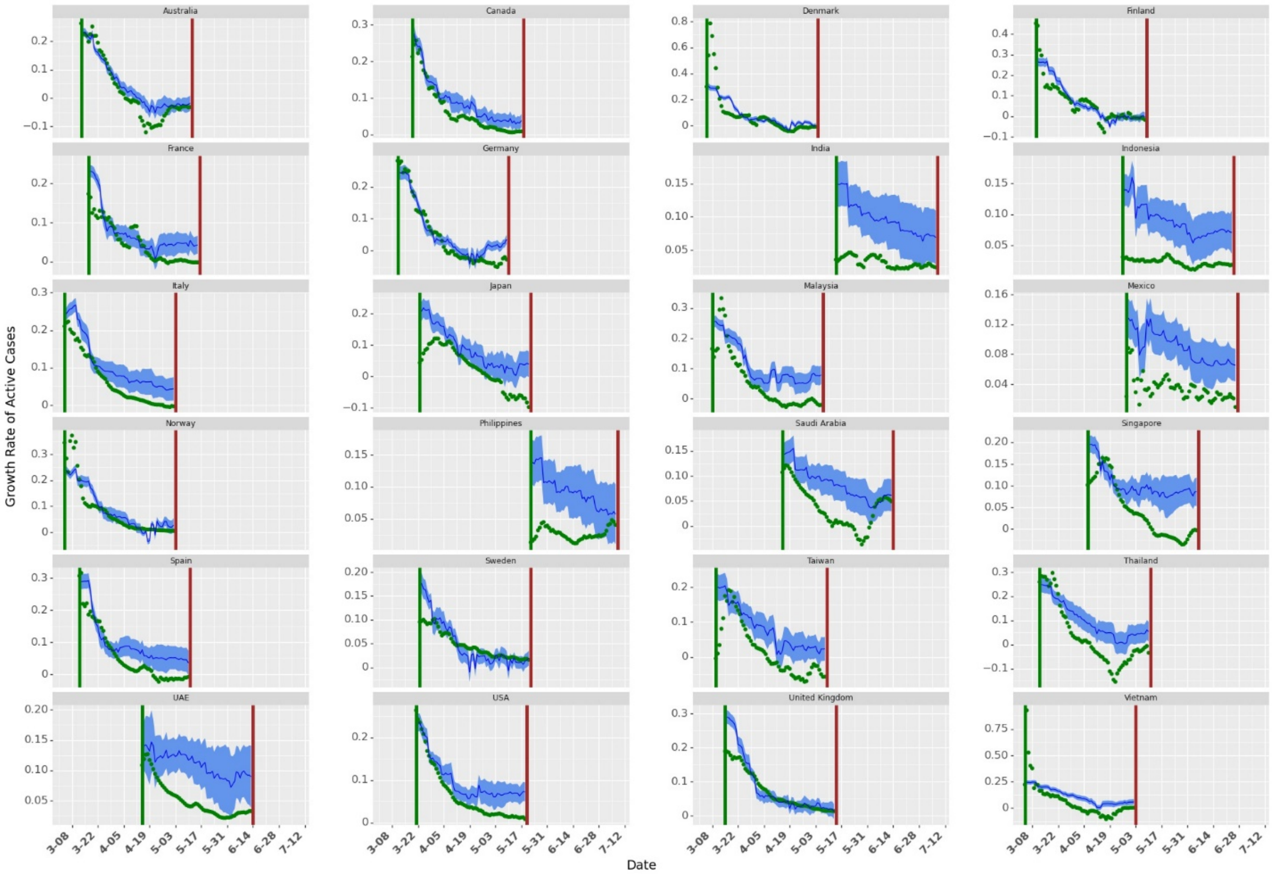


Figure S19. Growth Rate with no Mask Wearing. The green dots show the actual growth rates across countries. The dark blue line shows the mean of growth rate prediction (for 10000 samples in Krinksky-Robb method). The blue shaded area shows the confidence bounds around the mean prediction. We predict the growth rate when the masks are transformed as $\ln(1 + mask)$.

Similar to Figure S18, we can predict daily active cases with zero percent mask wearing as shown in Figure S20. The results show that masks lead to significant reduction in total cases as without these measures, the number of cases could exponentially increase over time (more discussion later on §Country wise effect),

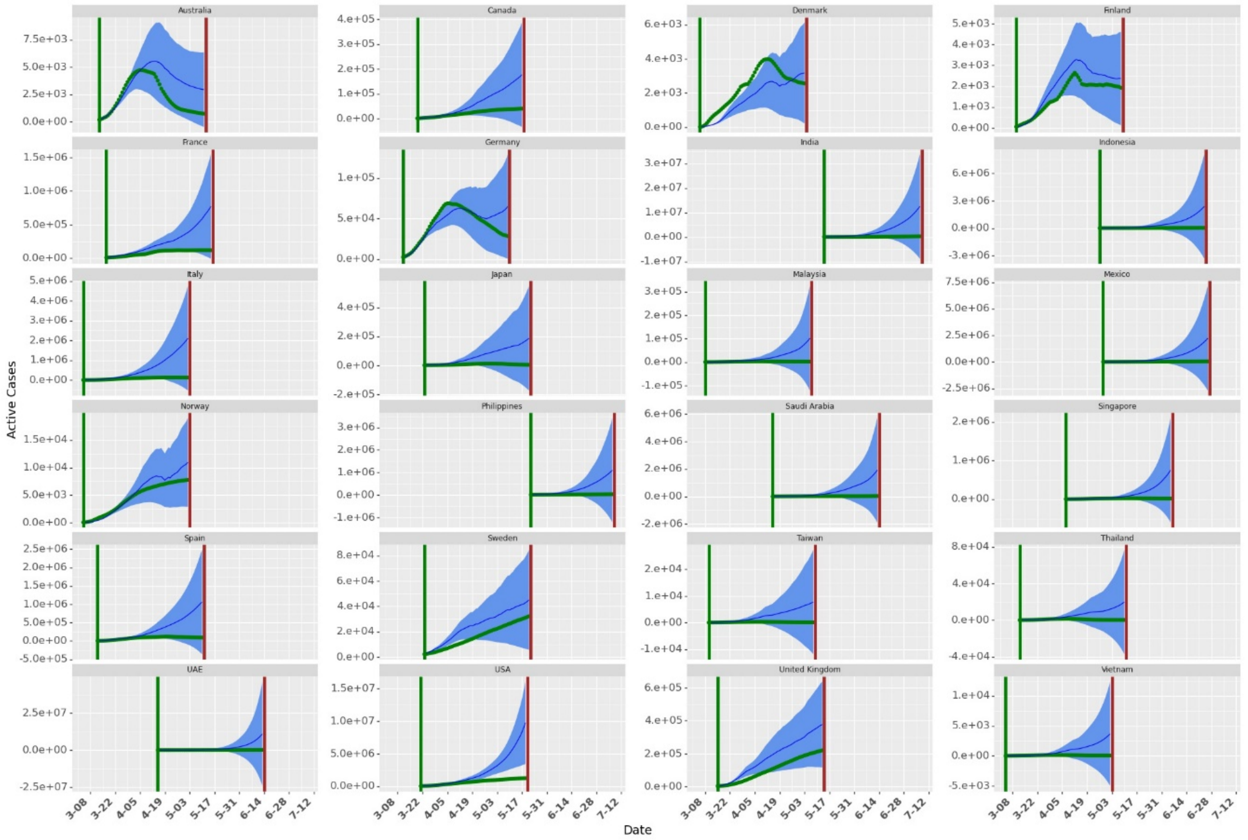


Figure S20. Simulation with no Mask Wearing. The green dots show the actual growth rates across countries. The dark blue line shows the mean of active cases prediction (for 10000 samples in Krinksky-Robb method). The blue shaded area shows the confidence bounds around the mean prediction.

We build five simulation models to further understand the impact of masks. In the first simulation model, we consider a hypothetical country with a constant value for all the covariates in the Equation S8. In the second simulation model, we check the change in active cases at the end of 60 days when mask wearing in a country is $m\%$ where $m \in [0,10,20,..,100]$. In the third model we check the change in active cases at the end of 60 days if the current levels of mask wearing is multiplied by a factor of $x \in [0,0.2,0.4,\ldots,2]$. In the fourth model, we present results for active cases at the end of 60 days when mask wearing percentage increases by $a\%$ as compared to the current levels in that country where $a \in [1,2,..10]$. In the fifth model, we exchange the mask wearing numbers between countries with minimum and maximum average mask wearing through the period of our analysis.

Simulation Model 1 helps in isolating the effect of masks as in this analysis, we do not consider any other covariates (as if no individual or institutional measures were taken apart from wearing masks in public areas). We construct data for a hypothetical country with these numbers to quantify the effect of masks in our analysis. In Simulation Model 2-4, we study country-wise association of masks with growth rate. In these models, we do not change numbers of any other covariates other than masks. These results show the potential change in active cases in that country for different percentage of people wearing masks in public. Simulation model 5 is used to build an approximate counter factual model for masks by exchanging mask wearing in countries with minimum and maximum average mask wearing during the period of our analysis. We discuss the results of these simulation models next.

*Simulation Model 1: Average country*

We simulate a hypothetical country with no wearing ($mask_{jt} = 0$), no active awareness ( $trend_{jt} = 0$), no testing( $testing_{jt} = 0$) and no government implemented NPIs ($s_{pjt} = 0$). We predict the active cases at the end of 60 days at different levels of mask wearing. We consider the average country with 0 country fixed effects for prediction. Daily active cases for this average country at different levels of mask wearing is shown in Figure S21. We assume that the average country has 100 cases on day 0 of simulation. Results show that increasing the mask number can help in flattening the curve (even when social mobility and NPIs remain unchanged). As the percentage of people wearing face masks increases, the daily active goes down as compared to no mask wearing. Results also show that social mobility along with NPI can also play significant role in flattening the curve (the daily active curve flatten even with no mask wearing, albeit slower). Cases starts rising again after initial flattening for most cases as social mobility increases and NPIs are relaxed (Figure S8). The results imply that if masks are mandated and its use widespread, complete lockdowns may be eased to help alleviate the associated economic hardships.

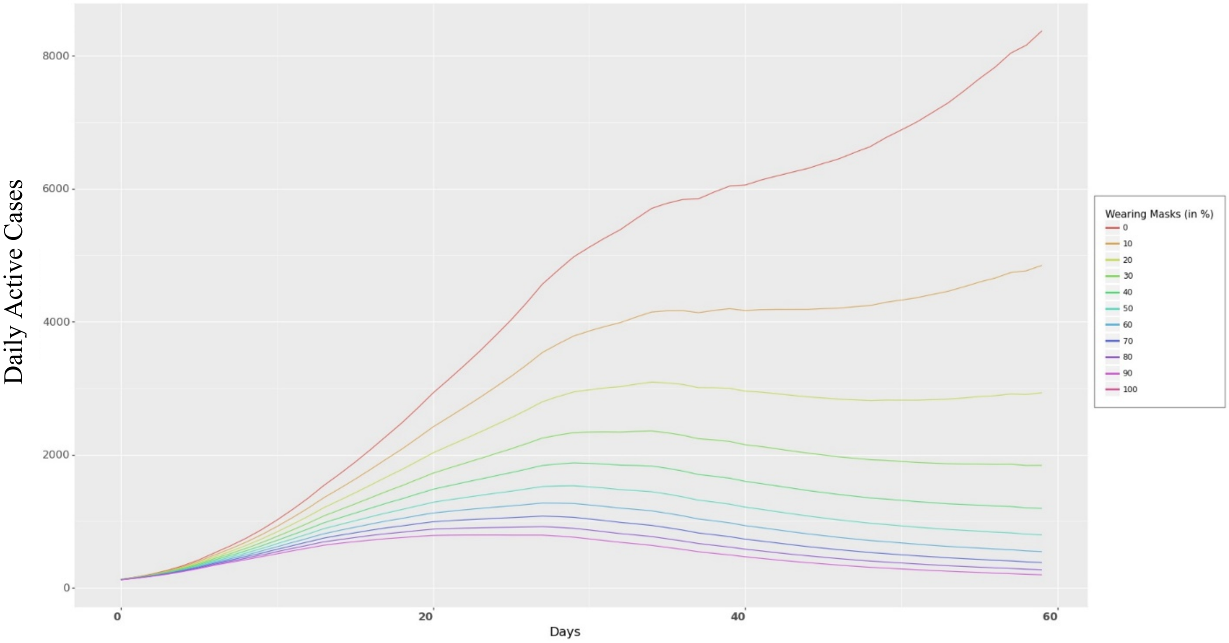


Figure S21. Daily Active Cases for the Average Country. As more people wear masks, the faster the curve can be flattened. Note that the graph shows the mean prediction under different levels of mask wearing. Also, it shows the daily active cases. Flattening of a curve has been synonymous with daily new cases, but if we use $I_t - I_{t-1}$, we can approximate daily new cases.

*Simulation Model 2: Changing mask levels*

In this simulation, we predict the number of active cases in each country by changing the levels of mask wearing. Figure S22 shows the ratio of active cases at the end of 60 days under different levels of mask wearing as compared to active cases at the current levels of masks. As the mask levels increase, the ratio of active cases to the true active cases at the end of 60 days decreases (Figure S22).

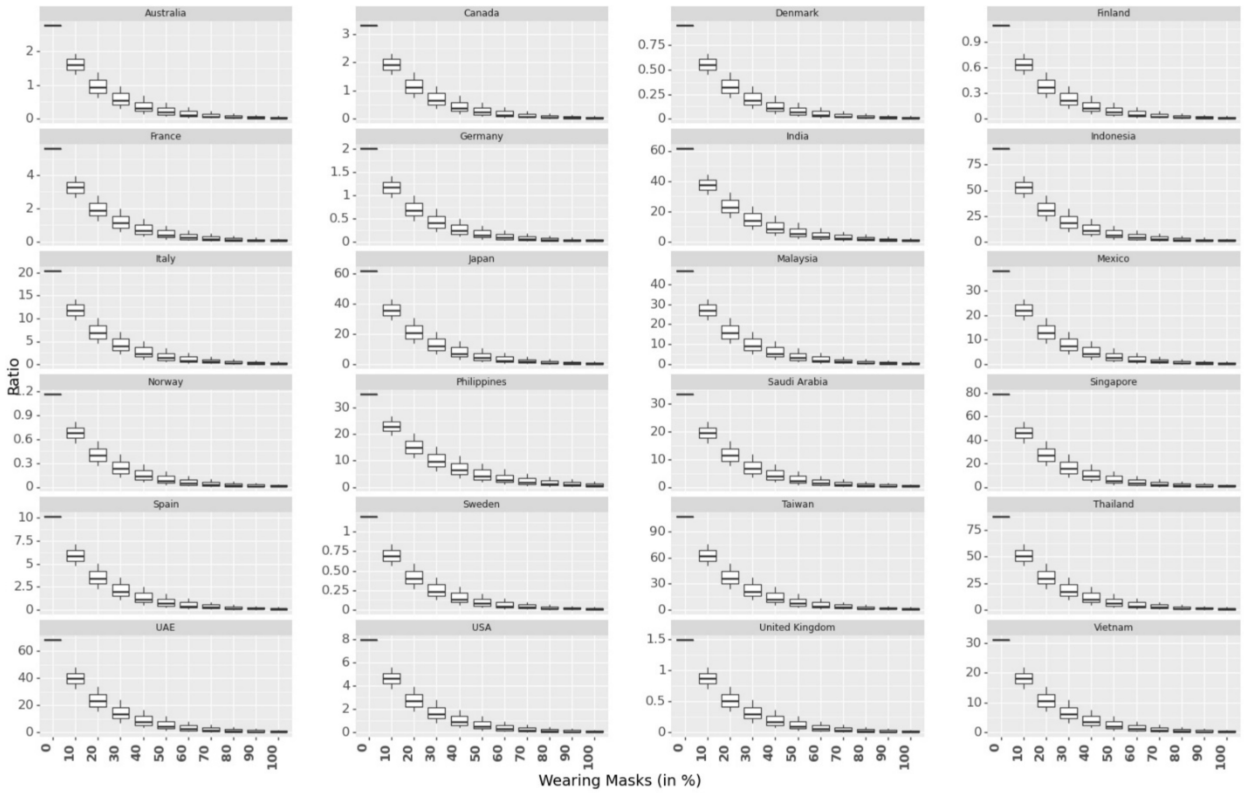

Figure S22. Box Plot on Ratio of Predicted Active Cases to Actual Cases at the End of 60 Days Under Different Levels of Mask Wearing.

*Simulation Model 3: Multiplying a constant to current mask levels*

In this simulation, we multiply the current mask wearing levels with a constant multiplication factor (0.2, 0.4, ..,, 2) to predict active cases at the end of 60 days as compared to actual scenario across 24 countries (Figure S23). Similar to Figure S22, we observe as we increase mask levels, ratio decreases significantly. However, the effect is different across different countries.

Similar to results in Figure S22, when the mask levels are much lower than the current levels (e.g. countries like Thailand, Vietnam, Singapore), the ratio of active cases at the end of 60 days (as compared to current levels of mask wearing) is much higher as compared to countries with lower current mask rates (e.g. Sweden, Norway).

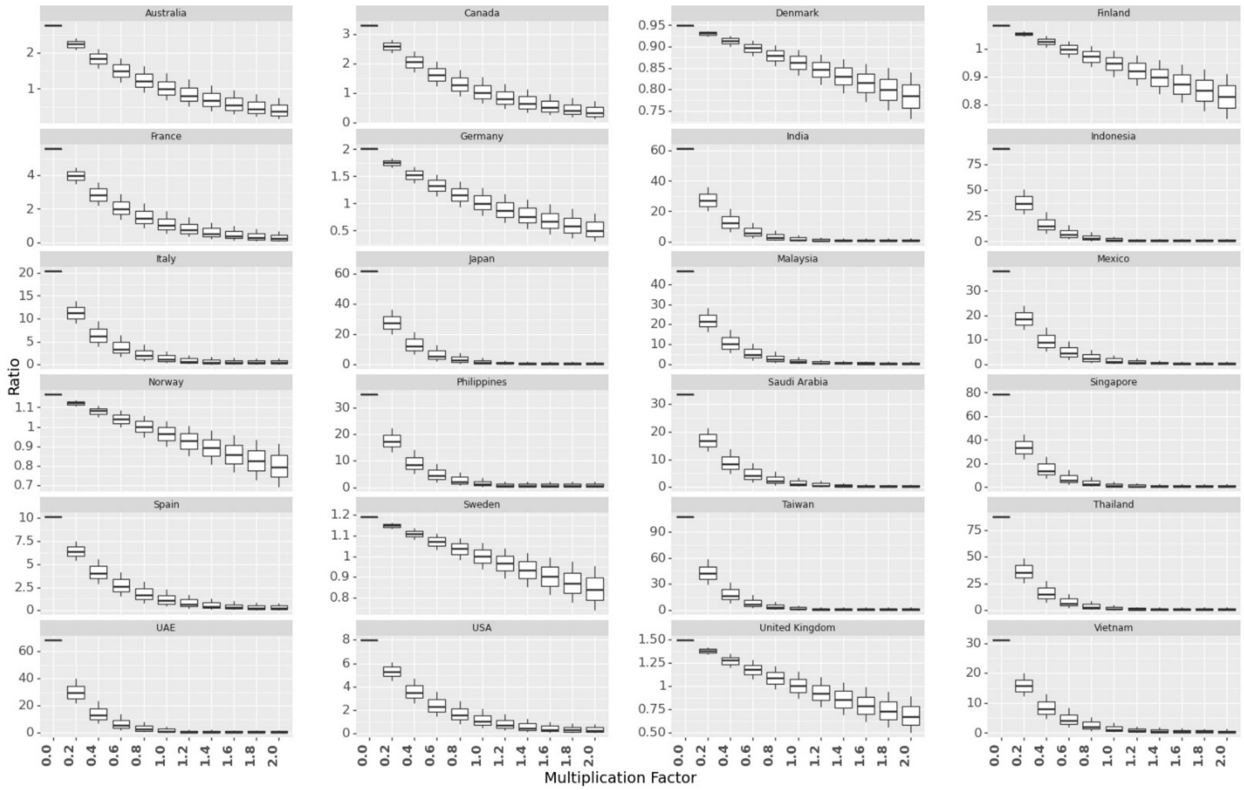


Figure S23. Box Plot on Ratio of Predicted Active Cases to Actual Cases at the End of 60 Days Under Different Levels of Mask Wearing obtained by multiplying the current levels of mask wearing with a constant.

*Simulation Model 4: Adding a constant to current mask levels*

In this simulation, we predict the ratio of active cases at the end of 60 days when mask wearing in a country is increased by different percentage points (0%, 1%, 2%, …, 9 %). Figure S24 plots the ratio of active cases at the end of 60 days with simulation for increased mask wearing to the actual active cases. This could help the government in forming policies that if $a$ % of more people follow the guidelines of wearing face masks, which NPIs could be relaxed while still controlling the spread of the virus. Enforcing mask wearing policy could be particularly useful in countries with low mask wearing.

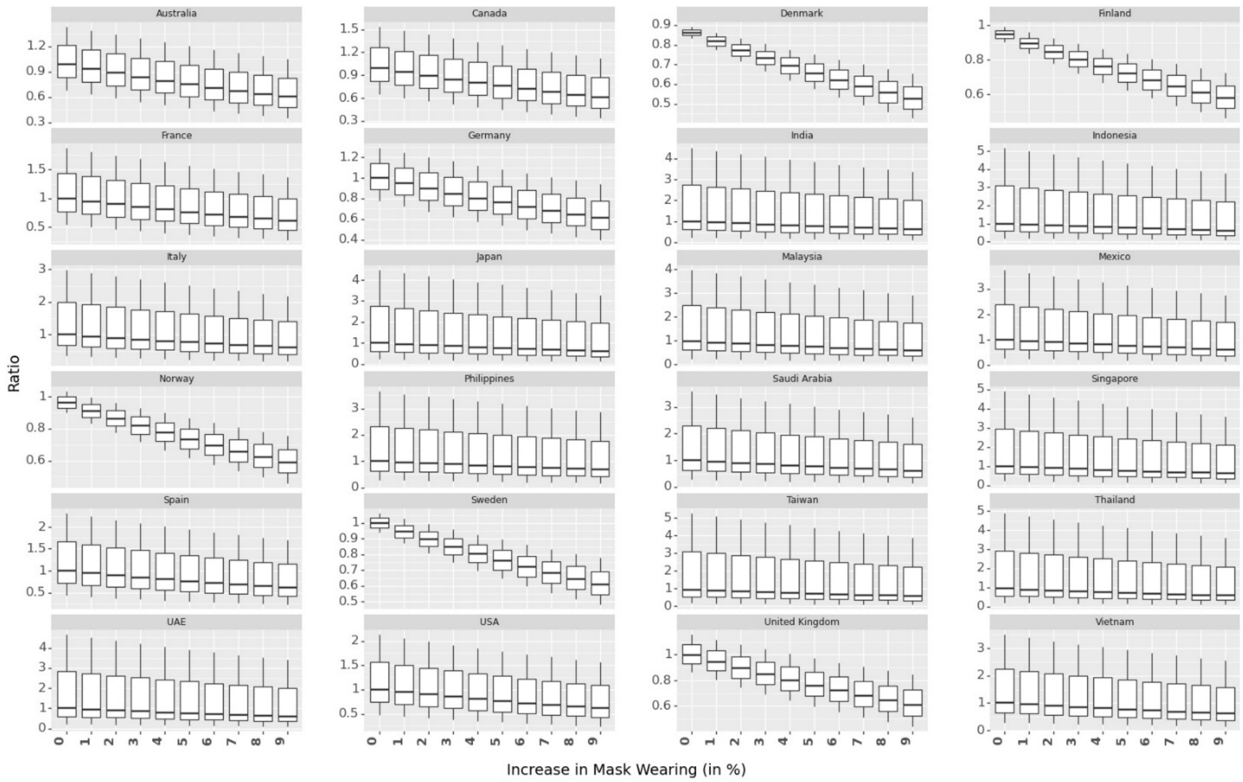

Figure S24. Box Plot on Ratio of Predicted Active Cases to Actual Cases at the End of 60 Days Under Different Levels of Mask Wearing obtained by increasing the current levels of mask wearing by different percentage points.

*Simulation Model 5: Exchanging the current mask levels*

Countries have observable heterogeneity in their culture of mask wearing (Figure S4). Mask wearing has been more common in Asian countries as compared to Scandinavian countries. In this simulation, we exchange the mask wearing numbers between 8 countries (4 Asian with highest average mask wearing among 24 countries and 4 Scandinavian countries with lowest average mask wearing among 24 countries). The ratio of active cases under new mask wearing as compared to actual active cases at the end of 60 days is shown in Table S7. Results show that Scandinavian countries could have reduced their confirmed cases significantly if they had enforced people to wear face masks in public. We find that the Scandinavian countries could have reduced the active cases by up to 50 times in 60 days if the citizens were wearing masks at the levels of Asian Countries.

Table S7. Ratio of Active Cases at the End of 60 Days after Exchanging Mask Wearing Numbers

| | **country** | **exchanged with** | **LL** | **M** | **UL** |
|---|---|---|---|---|---|
| max | Malaysia | Denmark | 23.86516 | **25.13209** | 26.44875 |
| max | Philippines | Finland | 12.00857 | **13.95786** | 16.19232 |
| max | Taiwan | Norway | 35.53529 | **39.52427** | 43.90114 |
| max | Thailand | Sweden | 33.90308 | **36.76336** | 39.82361 |
| min | Denmark | Malaysia | 0.009502 | **0.041856** | 0.180894 |
| min | Finland | Philippines | 0.00727 | **0.035283** | 0.167807 |
| min | Norway | Taiwan | 0.00432 | **0.025148** | 0.143133 |
| min | Sweden | Thailand | 0.004408 | **0.025332** | 0.142346 |

We present the combined effect of social mobility and NPIs. We provide a combined effect for social mobility and NPIs as it is difficult to estimate the causal analysis for individual variables. We use Krinsky-Robb method to estimate the combined effect of social mobility and NPIs. After drawing samples of coefficients of social mobility and NPIs using Krinsky-Robb method, we add the random samples draw and present the mean and confidence interval bounds of these samples as the combined effect and confidence interval bounds of that combined effect.

#### S3.3.2 Social Mobility

Parameter coefficients in Table S6 show that growth rate increases as mobility increases. This is because if people travel more or move to places with potential of public gatherings, infected individuals can spread the virus to the susceptible population. Governments therefore imposed strict restrictions to reduce mobility. We report the effect of mobility as negative of the coefficients in Table S13. Thus, we report the effect of mobility if the mobility numbers were 0 (no mobility change). Results in Figure S25 indicates that 0 change in mobility trends (no change in individual mobility trend indicates if people move around as they were before COVID-19) is associated with a daily increase in growth rate by 8.1% (5.6% - 10.6%) as compared to no mobility. Note that decrease in mobility can also be attributed to NPIs, no causality can be claimed on the effect of increase in mobility on growth rate.

The effect of full mobility across different countries is shown in Figure S25. It shows that even with mask numbers remaining unchanged and NPIs being implemented as they were implemented in that country, increasing mobility can lead to a significant increase in growth rate. Similar to Figure S20, we can predict daily active cases with full mobility as shown in Figure S26.

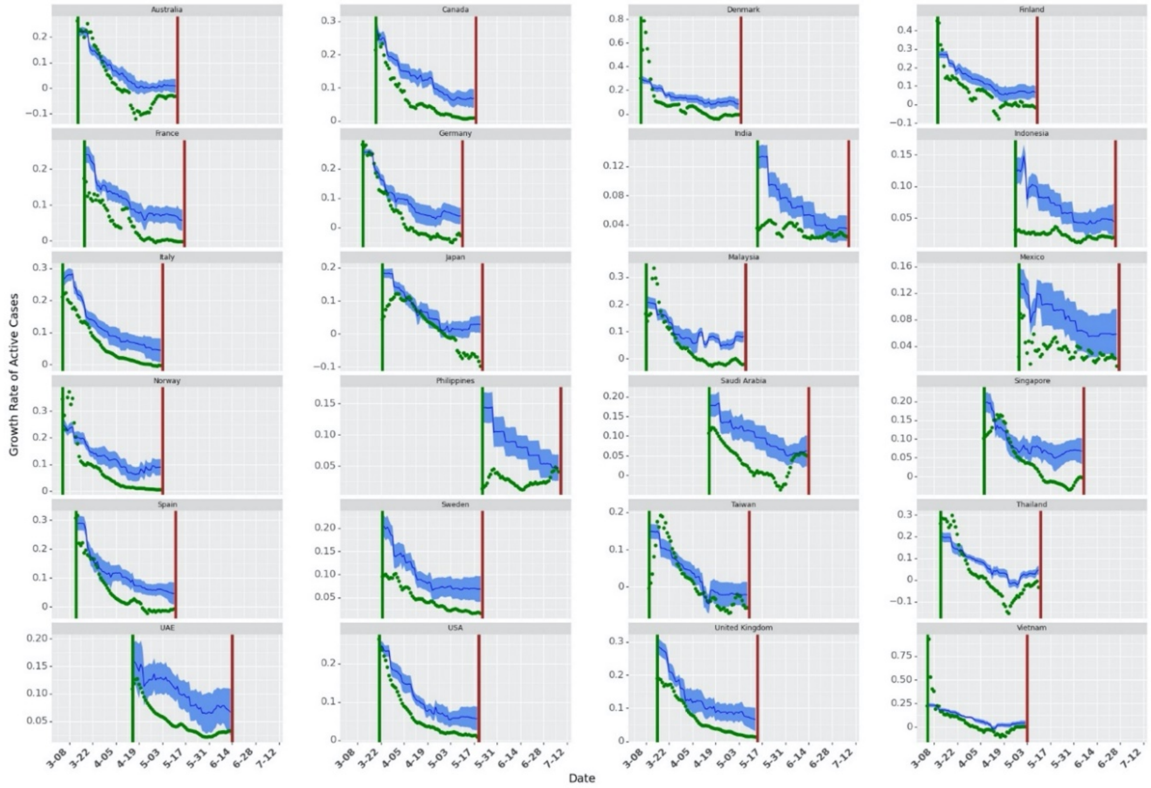


Figure S25. Growth Rate with Full Mobility. The green dots show the actual growth rates across countries. The dark blue line shows the mean of growth rate prediction (for 10000 samples in Krinksky-Robb method). The blue shaded area shows the confidence bounds around the mean prediction.

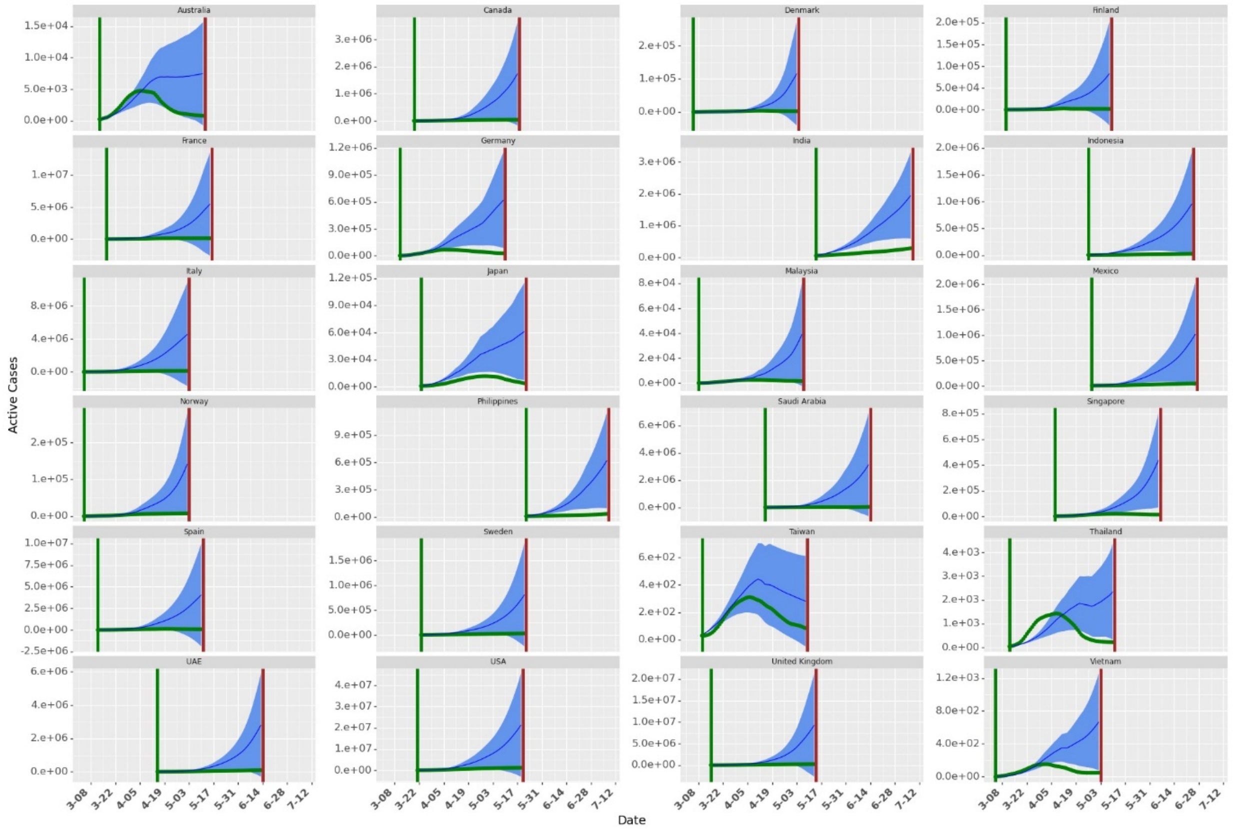

Figure S26. Simulation with no change in Mobility (as compared to pre-COVID-19 mobility). The green dots show the actual daily active cases across countries. The dark blue line shows the mean of active daily cases prediction (for 10000 samples in Krinksky-Robb method). The blue shaded area shows the confidence bounds around the mean prediction.

Similar to exchanging the mask wearing numbers among the countries with lowest and highest mask wearing percentages, we simulate for the active cases at the end of 60 days by exchanging social mobility among countries with highest and lowest social mobility. The results are shown in Table S8.

Table S8. Ratio of Active Cases at the End of 60 Days after Exchanging Social Mobility Numbers

| Social Mobility | Country | Exchanged with | Lower Limit for 95% confidence Interval | Ratio | Upper Limit for 95% confidence Interval |
|---|---|---|---|---|---|
| High | Philippines | Taiwan | 1.81473 | 6.940316 | 26.09045 |
| High | Denmark | United Kingdom | 2.495734 | 2.620681 | 2.750162 |
| High | Sweden | India | 1.163527 | 1.270406 | 1.385541 |
| High | Norway | Spain | 0.998357 | 1.100658 | 1.211927 |
| Low | Spain | Norway | 0.363724 | 0.947547 | 2.438379 |
| Low | India | Sweden | 0.026063 | 0.123776 | 0.576219 |
| Low | United Kingdom | Denmark | 0.314838 | 0.384042 | 0.467267 |
| Low | Taiwan | Philippines | 0.016745 | 0.098078 | 0.561585 |

#### S3.3.3 Non-Pharmaceutical Interventions (NPIs)

Negative and statistically significant estimate for the combined effect of NPIs show that NPIs helped in controlling the spread of virus. Results in Table S6 indicates that if mask wearing and mobility remains unchanged, implementing NPIs is associated to a daily drop in infectious cases by 13% (9.2% - 16.2%). Predicted growth rate and daily active cases with no NPIs is shown in Figure S27 and S28 respectively.

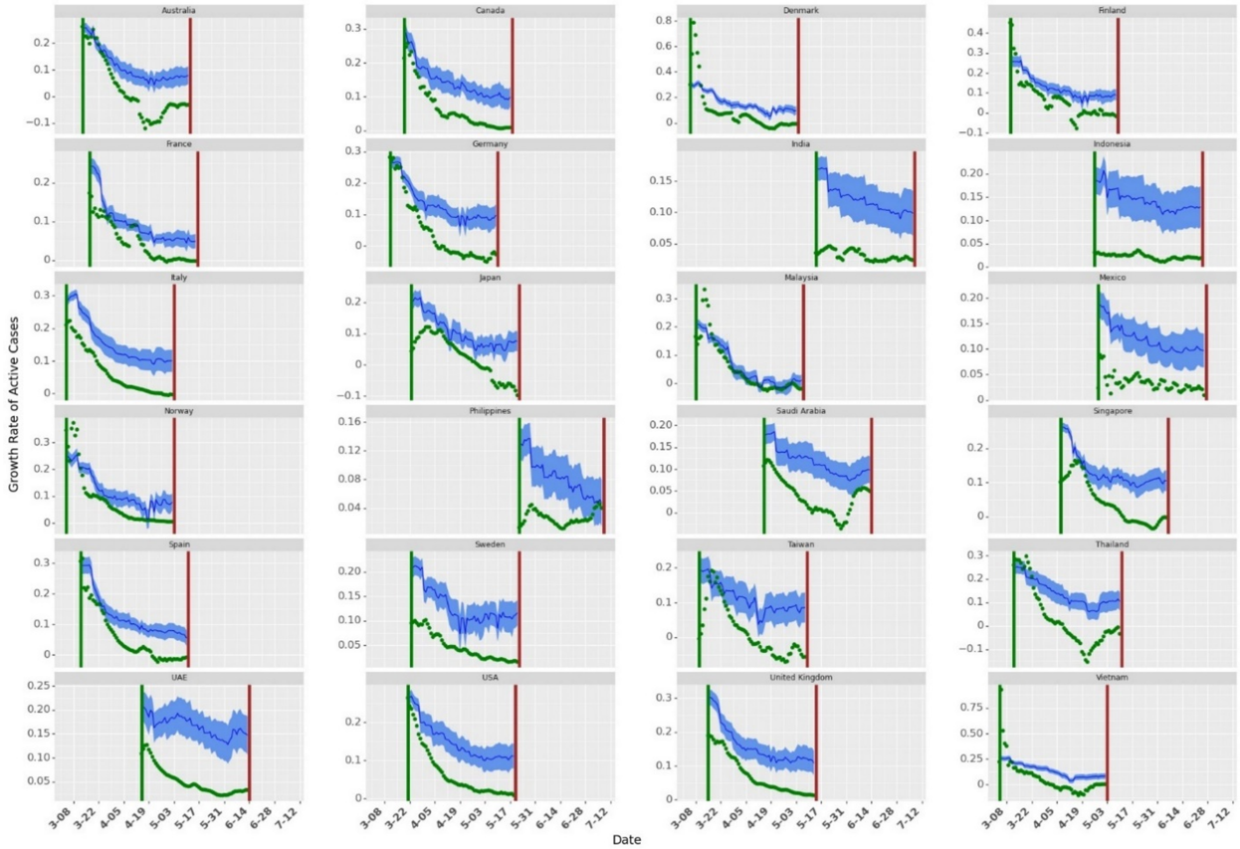


Figure S27. Growth Rate with no NPIs. The green dots show the actual growth rates across countries. The dark blue line shows the mean of growth rate prediction (for 10000 samples in Krinksky-Robb method). The blue shaded area shows the confidence bounds around the mean prediction.

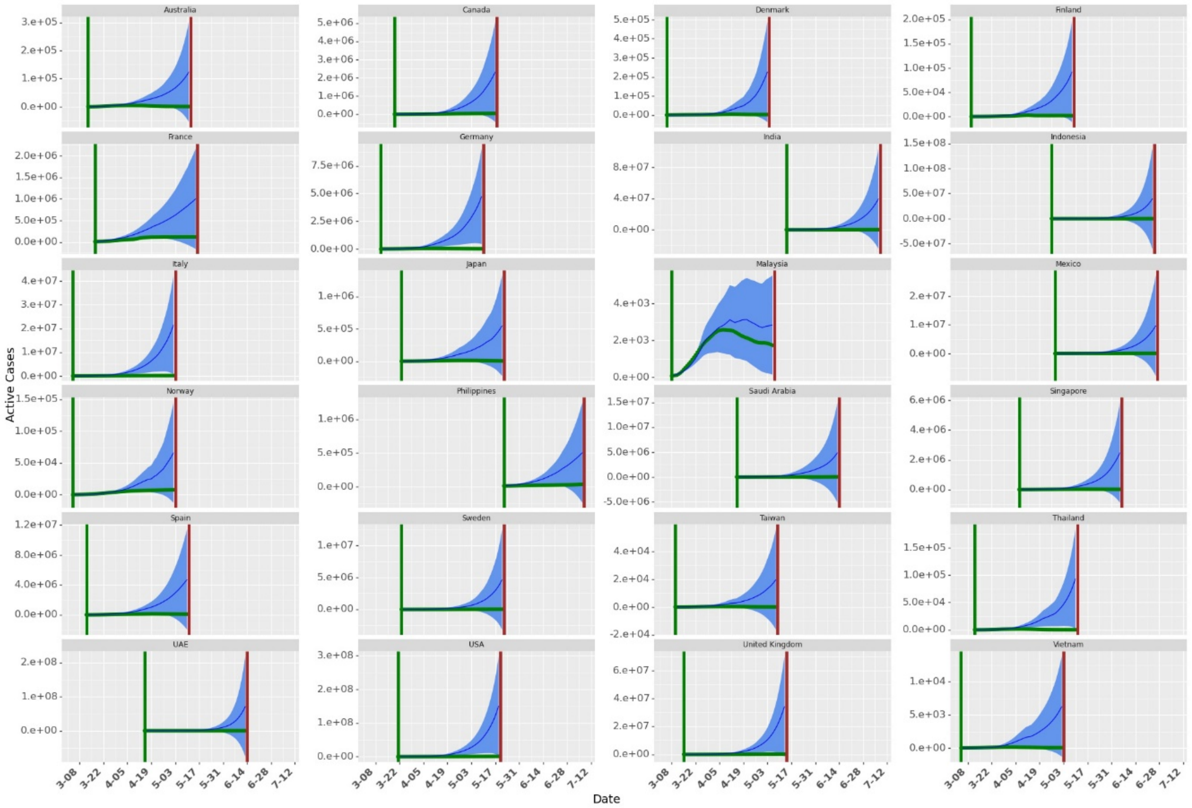


Figure S28. Simulation with no NPIs implemented. The green dots show the actual growth rates across countries. The dark blue line shows the mean of active cases prediction (for 10000 samples in Krinksky-Robb method). The blue shaded area shows the confidence bounds around the mean prediction.

Similar to exchanging the mask wearing numbers among the countries with lowest and highest mask wearing percentages, we simulate for the active cases at the end of 60 days by exchanging NPI numbers among countries with highest and lowest number of NPIs introduced across the country. The results are shown in Table S8.

Table S9. Ratio of Active Cases at the End of 60 Days after Exchanging Social Mobility Numbers

| NPI | Country | Exchanged with | Lower Limit for 95% confidence Interval | Ratio | Upper Limit for 95% confidence Interval |
|---|---|---|---|---|---|
| High | Italy | Finland | 2.551666 | 8.34897 | 26.90575 |
| High | Australia | Norway | 10.81348 | 17.51927 | 28.20859 |
| High | Thailand | Malaysia | 40.38261 | 209.2452 | 1061.602 |
| High | Singapore | France | 10.7138 | 59.20919 | 320.1261 |
| Low | Finland | Italy | 0.12681 | 0.13582 | 0.145342 |
| Low | Norway | Australia | 0.084334 | 0.092976 | 0.102375 |
| Low | Malaysia | Thailand | 0.001354 | 0.006118 | 0.027119 |
| Low | France | Singapore | 0.00753 | 0.015919 | 0.033334 |

#### S3.3.4 Combined Effect of Mask, Social Mobility and NPIs

Similar to presenting the combined effect of mobility and NPIs, we use the Krinsky-Robb method to present the effects for the combined effect of masks, social mobility and NPIs in daily drop in growth rate in Figure S29. Figure S29 also show the robustness of the model across different values of $shift$. It also shows the Mean Absolute Percentage Error for 10 – fold cross validation used to get the $shift$ that best fit the data. We observe best data fit for a lag of 9 days. Grey vertical lines indicate a shift of day 7 and day 11. The combined effect of masks, social mobility and NPIs is estimated to be a 28.1% (24.2%-32%) drop in daily growth rate.

Results show that the effect of masks remain consistent across different transformations and different $shift$. We also observe consistency across different transformations of mask numbers. Furthermore, the total combined effect of masks, social mobility and NPIs remain consistent as we change $shift$.

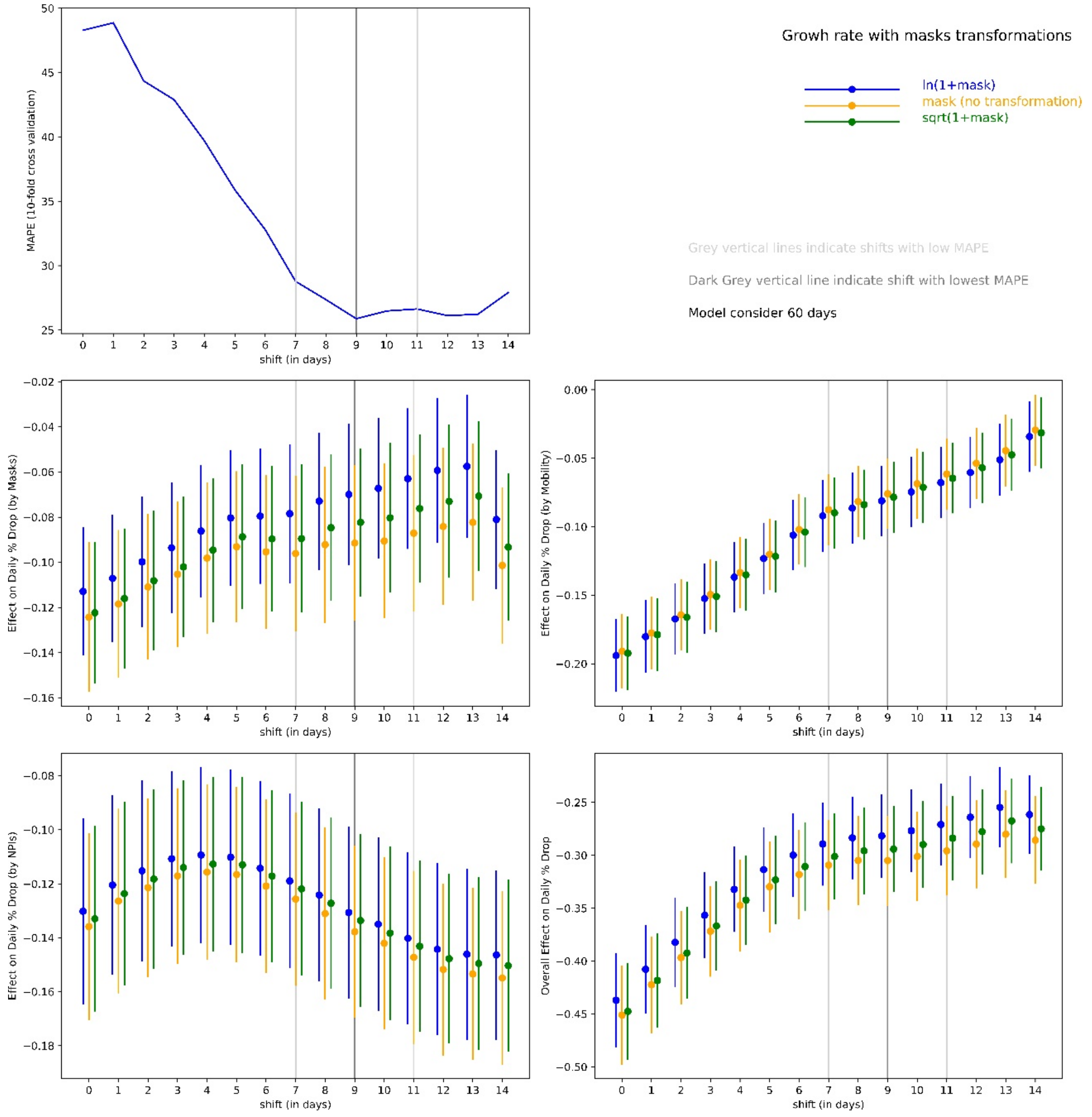


Table S29. Combined effect of Mask, mobility and NPIs in daily % drop in infectious cases

The results show that masks, social mobility and NPIs lead to significant reduction in total cases as without these measures, the number of cases could exponentially increase over time.

#### S3.4 Testing and Google Trends

Negative and statistically significant coefficient for testing and Google Trends indicate a drop in daily growth rate as these numbers increase. As testing increases, it may show increased daily confirmed cases (as more people get tested and it can discover asymptomatic cases). However, our model uses a lag of 9 days. Thus the cases may not be affected by testing immediately. Thus, a negative coefficient with a lag shows that testing helps in achieving a daily drop in growth rate in active infectious cases. Similar to testing, as our model uses a lag of 9 days, increased google trends indicate increased awareness among the citizens regarding COVID-19 which may lead to more caution against COVID-19.

### S4. Robustness Checks

#### S4.1 Selection of $th$ and $shift$

To filter the initial volatile growth rate, we use a threshold in the model (one for each country). We start collecting data for a country from the day respective countries reach their threshold. We define threshold as -- the day after which the 7-day average of daily new cases were $th$ % of the peak daily new cases observed in that country. Decreasing $th$ will add noise to the model due to high volatility in early the growth rates. However, if the threshold is high, we miss out on important data, particularly during the initial phase when the growth in infections is exponential.

Along with $th$, we also use $shift$ in the growth rate model to capture the delay in effect of mask, Non-Pharmaceutical Interventions (NPIs) and mobility. First we find the value of $th$ and then use that $th$ to find optimal $shift$ for our analysis. We calculate log likelihood for growth rate model under different $th \in (0.01, 0.02, \ldots, 0.3)$. For each $th$, we run the model for different $shift$. Using multiple $lag$ ensures that the model performance is consisted with different values of $shift$. It also ensures that we do not select $th$ that performs well by chance. We select $th$ based on maximum average log likelihood for different $shift$. We start with threshold value of 0.01 and starts increasing. Figure S30 shows that the average log-likelihood for does not change much after $th = 0.18$. We use a $th = 0.20$ in the rest of the paper. We use sensitivity test to check the consistency of the model.

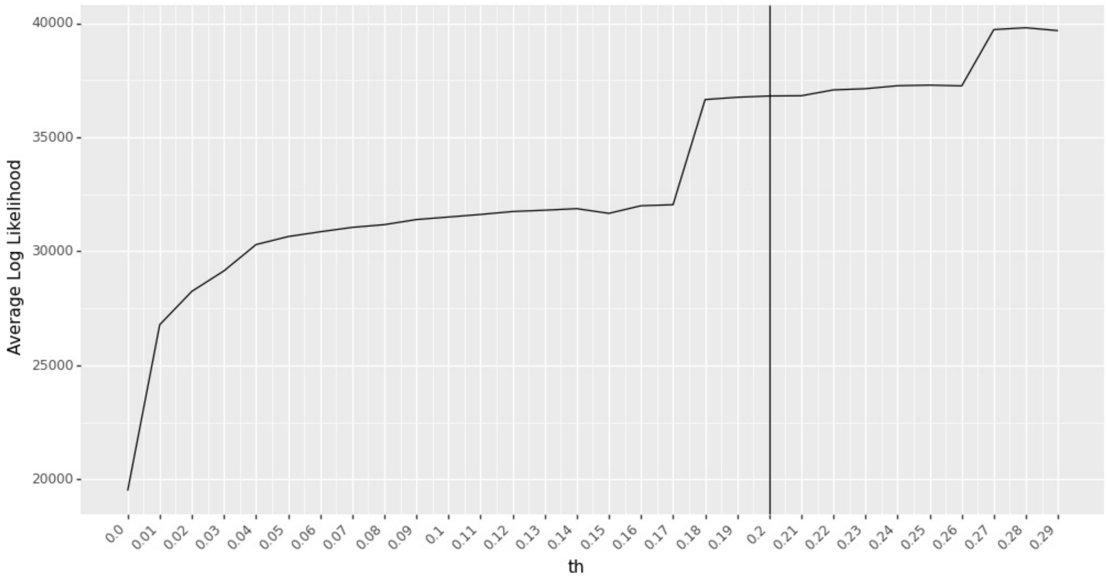


Figure S30. Average Log-likelihood Values for Different $th$. The model performs the best (based on maximum likelihood function) when $th = 0.28$. However, the performance change is not significant after $th = 0.18$. So we use $th = 0.2$ in our analysis in this research.

In the growth rate model, we use $shift$ to estimate the parameter coefficients as shown in Equation S9. After we select $th$, we use cross validation to find $lag$ that best fits the data. We use 10-fold cross-validation with Mean Percentage Error (MAPE) as a metric for out of sample data points to select $shift$ with the best fit. The average MAPE (for 10-fold cross validation) for different lags is shown in Figure S31. $shift = 9$ days shows the best fit with minimum value for average MAPE for 10-fold cross validation.

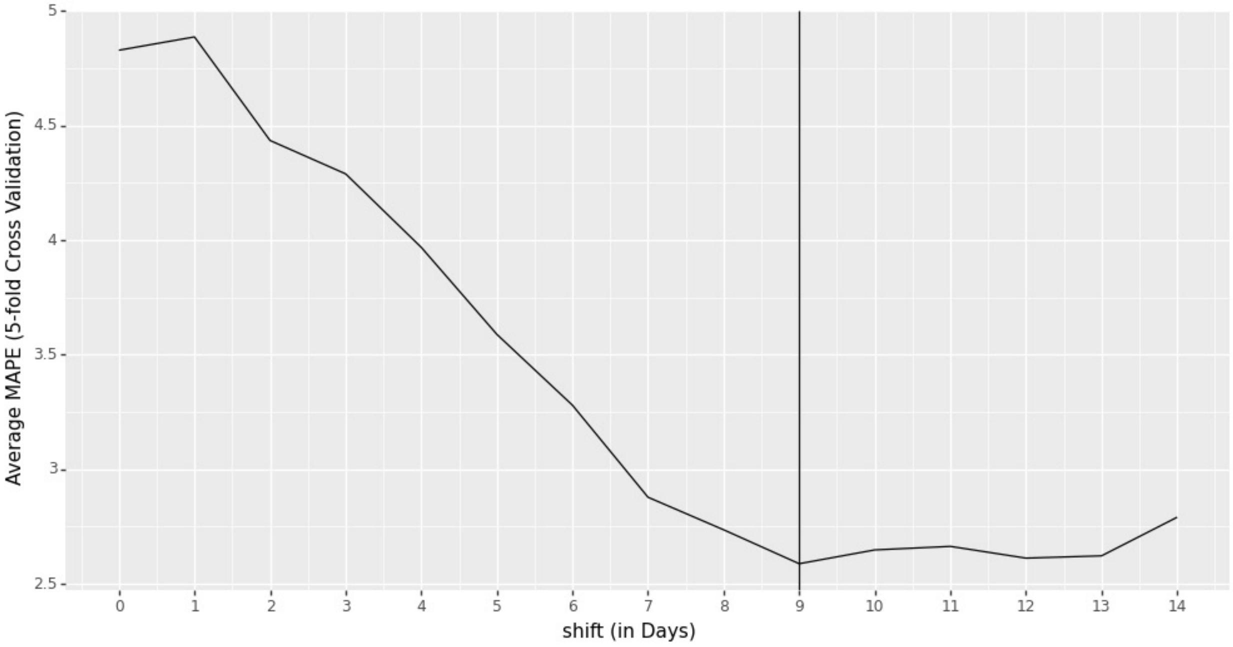


Figure S31. Average MAPE from 10-fold Cross Validation Different $lag$. Minimum value for Mean Absolute Percentage Error (MAPE) is obtained at a $lag = 9\ days$.

### S4.2 Model Estimation Sensitivity to $th$ and $shift$

We use sensitivity test to check the consistency of the parameter estimates for different values of $th$ and $shift$. The performance of the model remains consistent on changing the values of $th$ from 0.18 to 0.22 as shown from parameter estimates in Figure S32 (we use a $shift$ of 9 days). The performance of the model remains consistent on changing the values of $shift$ from 7 days to 9 days as shown from parameter estimates in Figure S33 (we use $th = 0.2$). We transform masks as $\ln(1 + mask_{jt})$.

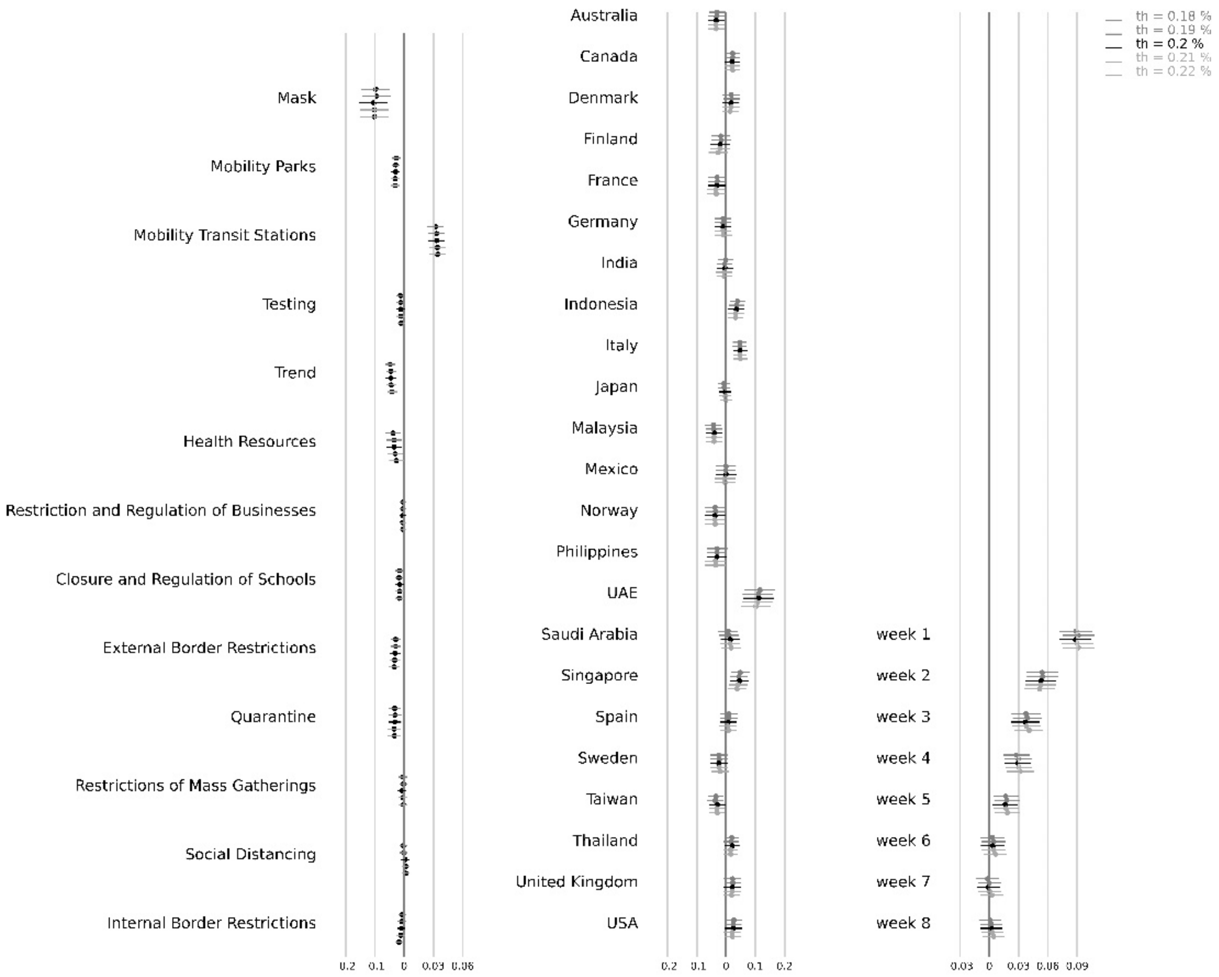


Figure S32. Parameter Estimates for Growth Rate Model for Different $th$. Horizontal lines represent the upper and lower confidence interval bound for the parameter estimates. We show the results for a lag of 9 days with $th \in [0.18, 0.22]$. The results indicate that the model is robust to different lags as the parameter estimates show consistency. Vietnam has been kept as the base country (the fixed country effect is 0).

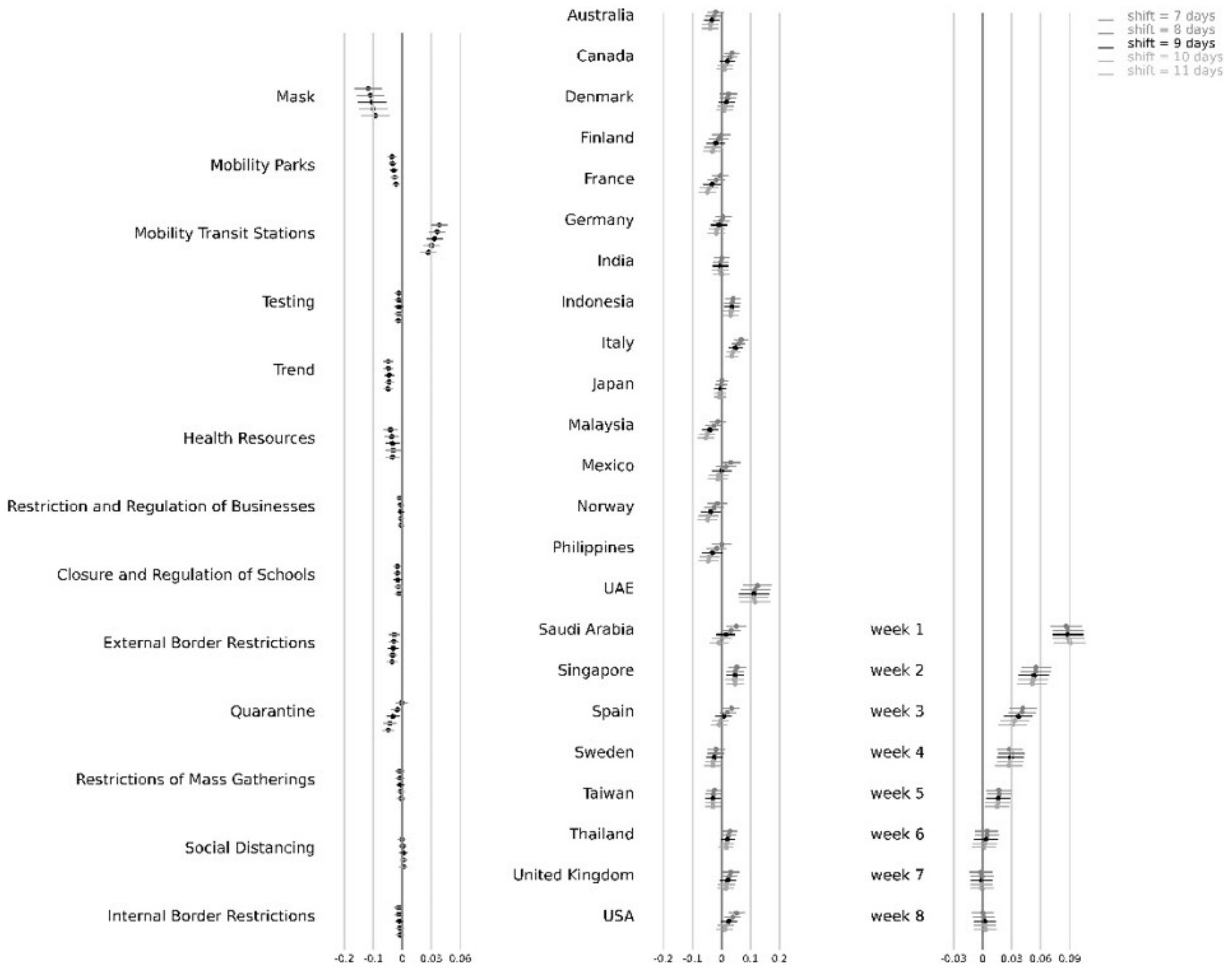


Figure S33. Parameter Estimates for Growth Rate Model for Different $shift$. Horizontal lines represent the upper and lower confidence interval bound for the parameter estimates. We show the results for a lag of 7 days to 11 days with $th = 0.2$. The results indicate that the model is robust to different $shift$ as the parameter estimates show consistency. Vietnam has been kept as the base country (the fixed country effect is 0).

#### S4.3 Handling Data Error in Active Cases

In Figure S5, we observe data reporting issue in Norway, Sweden and United Kingdom. We observe that the recovered cases are reported very late in Norway, not reported in Sweden and recorded very in United Kingdom. We use data from Johns Hopkins Resource Center. In our analysis, we use total confirmed cases to find active cases for these three countries. However, this may lead to bias in the results. To check the bias, we run the model without these three countries. The parameter estimates after excluding these countries is shown in Figure S34. In Figure S18, we show the parameter coefficients when $shift = 9$ days and $th = 0.2$.

The results show that the model parameter estimates are robust to exclusion of Norway, Sweden and United Kingdom from the model. However, the model slightly overestimates the coefficient of masks after excluding Norway, Sweden and United Kingdom (as compared to the growth model with data from all the 24 countries). All the three countries are in Europe where wearing face masks is not as common as Asian countries. Moreover, Norway and Sweden (along with Denmark and Finland) have the lowest percentage of people who wear face masks in public in our data set.

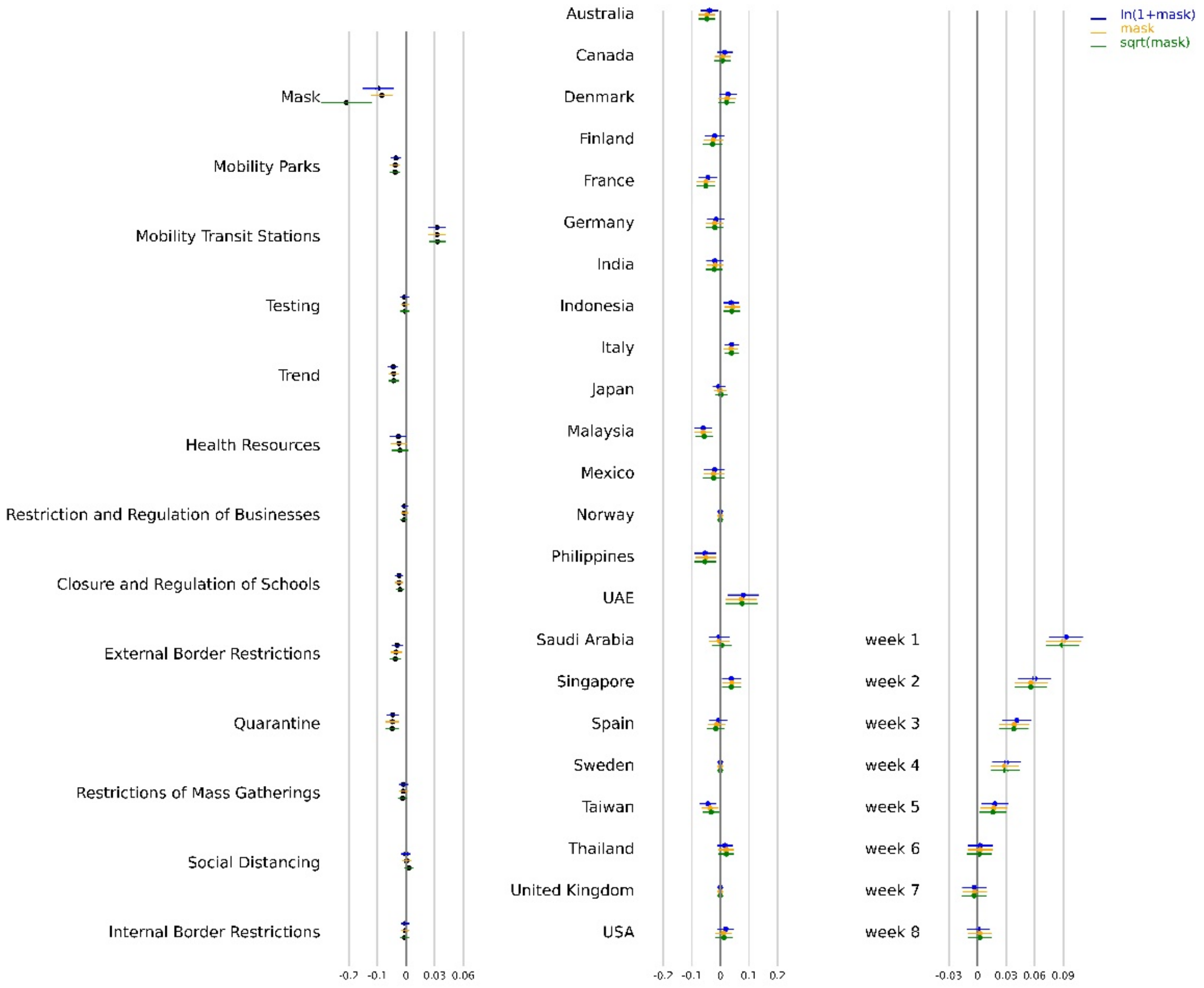


Figure S34. Parameter Estimates for Growth Rate Model after excluding Norway, Sweden and United Kingdom from the analysis. Horizontal lines represent the upper and lower confidence interval bound for the parameter estimates. We show the results for different transformations of $mask_{jt}$. The results indicate that the model is robust to different transformations as the parameter estimates show consistency. Vietnam has been kept as the base country (the fixed country effect is 0)

### S4.4 Robustness Check for Mobility

In our analysis, we used Google's community mobility report as a measure of social mobility. We used Google's Community reports numbers as a measure of social mobility as android operating devices are more common than iOS, particularly in Asian countries [14]. Also, Apple's Community Mobility Reports record data only when an individual opens Apple Maps. To check the robustness of the combined effect of masks, social mobility and NPIs, we also consider the apple's community mobility numbers as a measure of social mobility. Apple released data for change in trend for driving and walking for all the 24 countries considered in this work. The combined effect after substituting with apple's mobility report is shown in Figure S35. Consistency of the results show that the estimates for the model are robust.

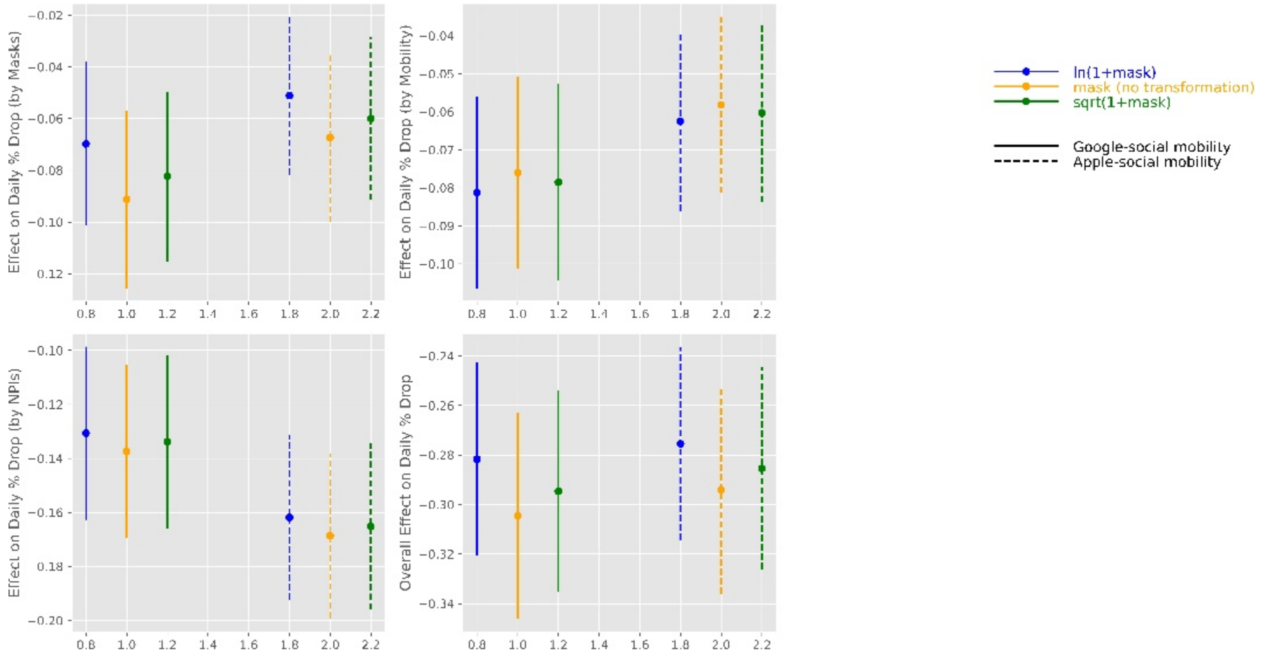


Figure S35. Combined effect of masks, social mobility and NPIs with mobility numbers from Google and Apple

#### S4.5 Alternative Specifications

We build two robustness model to check the consistency and reliability of the parameter estimates of the growth rate model. In the first robustness check, we use exponential smoothing as shown in Equation S9-S10. In the second model, we use a control function approach to identify the impact of masks on the spread of COVID-19. We discuss it in details in this Section.

##### S4.5.1 Model 1: Exponentially Smoothed Variates for Growth Rate

In the first specification, we use exponential smoothing to estimate the parameter estimates for masks, NPIs and social mobility. In our base model in Equation S8, growth rate is defined as a function of masks, NPIs, social mobility, trend and testing at a lag of $lag$ days. On any day $t$, this model ignores the value of the variates from days $t - lag + 1$ to $t$ (discussed in Figure S1). In this model, we do not ignore variates between $t - lag$ and $t$ and use exponential smoothing average to check the robustness of our model. To check the consistency of the parameter estimates for different transformations of masks (top left), mobility (top right), NPIs (bottom left) and the combined effect of masks, social mobility and NPIs (bottom right) is shown in Figure S36.

We also show the parameter estimates from the growth model for comparison. The results show that the parameter estimates for both the models are close and consistent, thus showing the robustness of the results in Table S6.

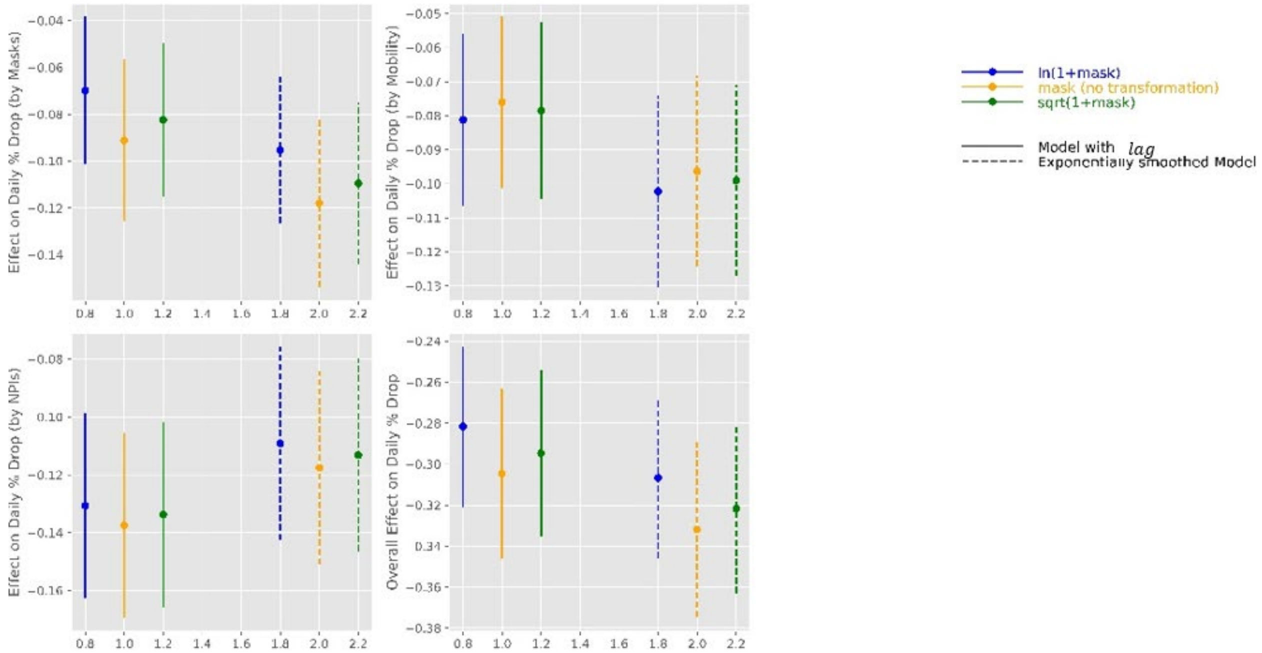

Figure S36. Effect on daily % drop in growth rate by Masks, Mobility, NPIs and the Combined effect of masks, social mobility and NPIs with $shift = 9$ days for exponentially smoothed model.

In the next model, we use a control function approach to check the robustness of our estimates in Table S6. Control function approach considers an error variable based on an exogenous variable which is not correlated with response variable but is correlated to an instrumental variable. We use number of deaths per thousand people for SARS, H1N1 and MERS CoV ad our instrumental variables.

#### S4.5.2 Model 2: Control function Approach to Growth Rate

In the control function approach, we first predict the average value of $mask_{j,t}$ by using the number of deaths per thousand people in each country by SARS, MERS-CoV and H1N1. Results from predicting masks using disease per thousand people is shown in Table S10 and the parameter estimates are shown in Table S11.

Note that we consider all the available data set (from February 21, 2020 to July 8, 2020) to estimate the coefficients for SARS, H1N1 and MERS. We convert the numbers for deaths due to SARS, H1N1 and MERS into binary variable (1 if the number for a country is greater than the median).

Table S10. Results Statistics for Predicting $log(1 + mask)$ using SARS, H1N1 and MERS

| | |
|---|---|
| R-squared: | 0.056 |
| Adj. R-squared: | 0.055 |
| F-statistic: | 65.24 |
| Probability (F-statistic): | 0 |
| Log-Likelihood: | -8108 |
| AIC: | 16200 |
| BIC: | 16250 |
| No. Observations: | 3312 |
| Degree of Freedom: Residuals: | 3308 |
| Degree of Freedom: Model: | 3 |

Table S11. Results from Predicting $log(1 + mask)$ using SARS, H1N1 and MERS

| coef | **std** | **err** | **t** | **P>\|t\|** | **[0.025** | **0.975]** |
|---|---|---|---|---|---|---|
| Sars | 0.5013 | 0.179 | 2.797 | 0.005 | 0.15 | 0.853 |
| H1N1 | -1.3904 | 0.101 | -13.78 | 0 | -1.588 | -1.193 |
| Mers | 0.1556 | 0.114 | 1.363 | 0.173 | -0.068 | 0.38 |
| const | 6.9685 | 0.071 | 98.706 | 0 | 6.83 | 7.107 |

We use $mask_{jt}$ along with the residuals from prediction model, $e = mask_{jt} - \widehat{mask_j}$, as control function in Equation S9. The results for the combined effect of masks, social mobility and NPIs is shown in Figure S37. We use a $shift$ of 9 days. We also show the combined effect of masks, social mobility and NPIs without control function (focal model in this paper) to show that the combined effect of masks, social mobility and NPIs estimated in Figure 21 are not appreciably different.

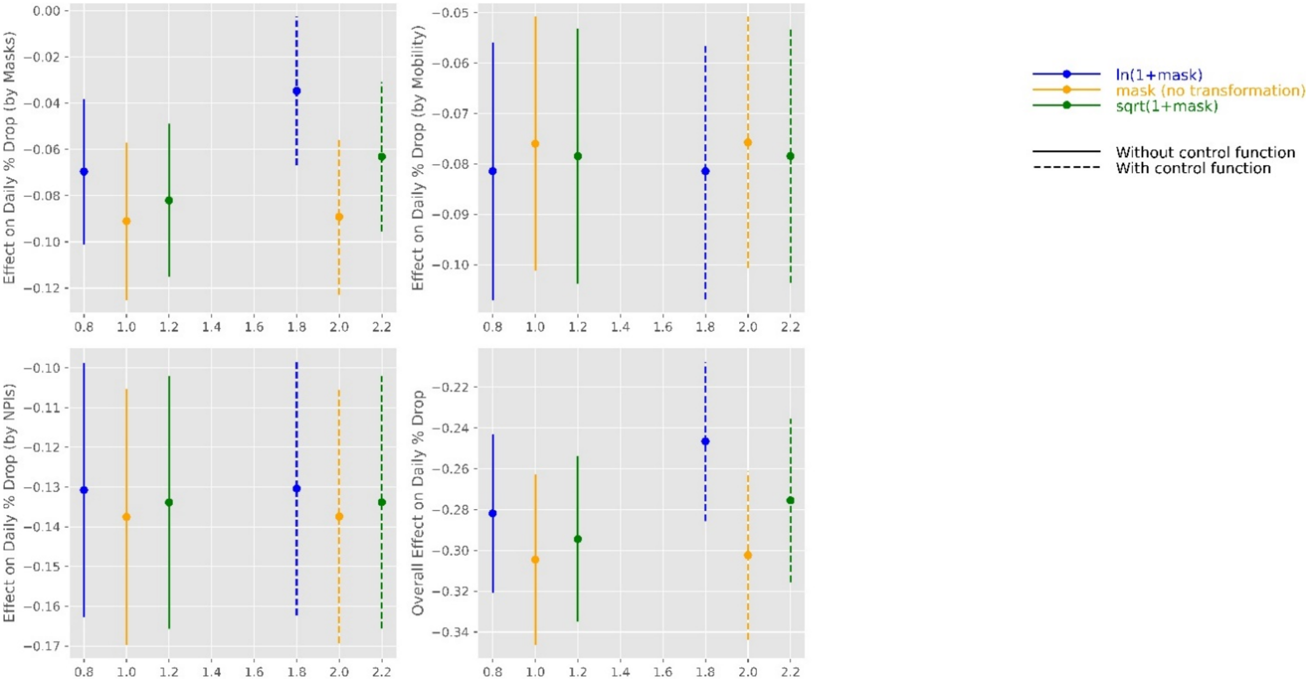


Figure S37. Combined effect of Mask, Social Mobility and NPIs with and without control functions. We use growth rate model with a $shift$ of 9 days.

#### S4.5.3 Lasso Regression

Governments across the world introduced NPIs to enforce social distancing through policies like quarantine, restriction on mass gatherings or closure of schools and businesses. NPIs led to decreased social mobility. For example, there were no major gathering in railway or bus stations as rails and buses were closed down. In our analysis, NPIs and social mobility across different location types are correlated. This may lead to multicollinearity that may lead to unstable coefficients. As a robustness check, we use penalized linear regression (Lasso regression) to shrink the coefficients of highly correlated variates.

Lasso regression can also handle multicollinearity in the data as it shrinks the coefficients to 0 using L1-norm. Lasso regression pushes the coefficients of insignificant variables to 0, thereby introducing sparsity in the model. Lasso regression pushes the coefficients for all the indicators of social mobility except mobility in parks and transit stations. As we observe the correlation between different indicators of social mobility in Table S2, Lasso regression provides a validation for the selection of two (out of 6) indicators of mobility. Coefficients for the growth model are shown in Figure S38. Lasso regression model is given in Equation S14 where $n$ is sample size, $\beta$ is a vector of coefficients and $Y$ is outcome variable. We use 5-fold cross-validation to find $\lambda$ that best fits the out of sample test data.

$$\beta = \arg\min_{\beta \in R^p} \left\{ \frac{(Y - X\beta)^2}{n} + \lambda ||\beta||_1 \right\} \qquad (S14)$$

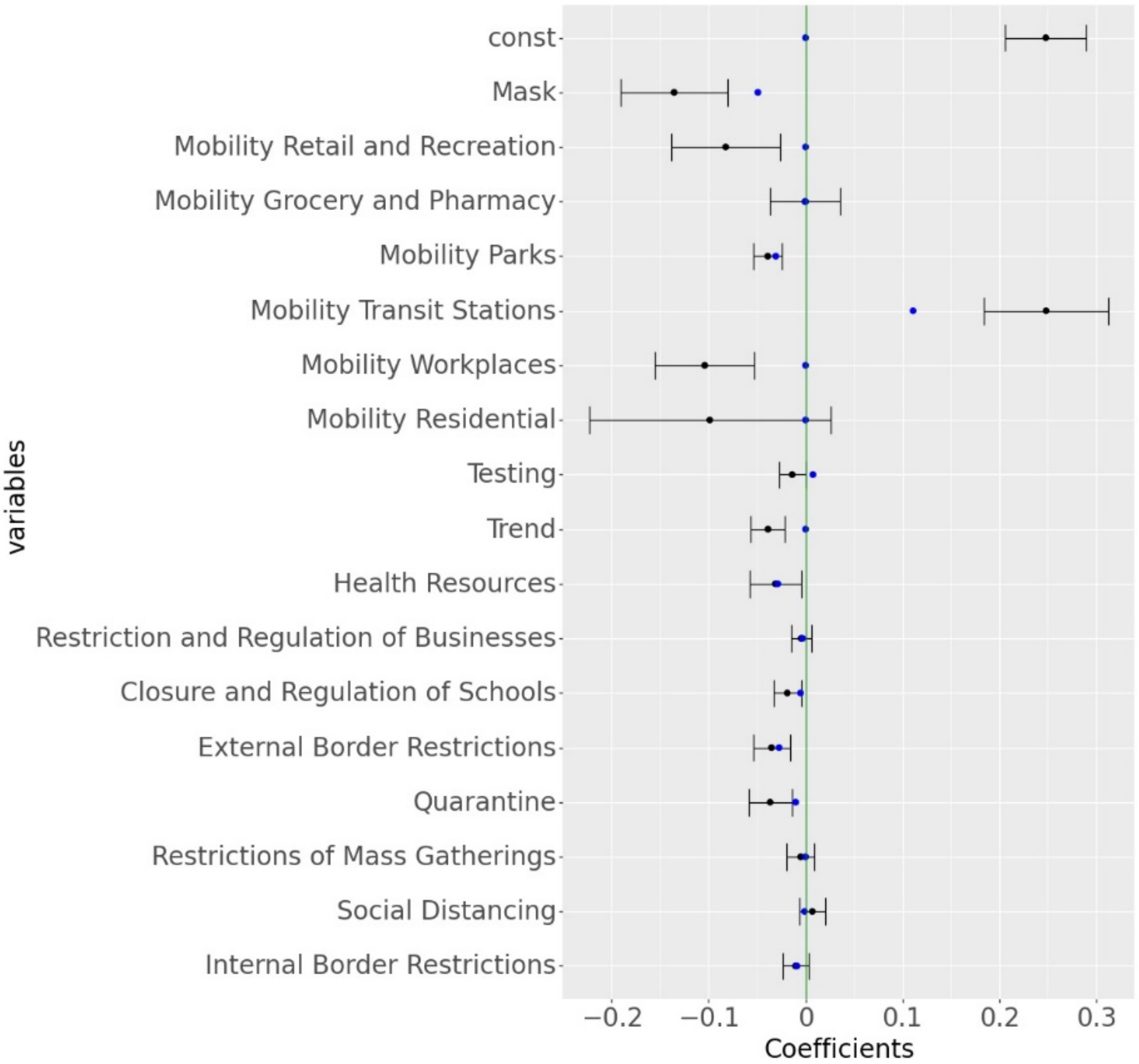


Figure S38. Parameter Estimation from Ordinary Least Square and Lasso Regression Model for growth rate. The blue dot represents the coefficients estimated from the Linear Regression model. The error bars represent the upper and lower confidence interval for the coefficients obtained from Ordinary Least Squares. The blue dot represents the coefficients estimated from the Lasso Regression model.

### S4.6 Selecting Period of Analysis in the Analysis

To filter out initial volatile growth rates during the start of the pandemic, we use a threshold $th$ as discussed before. We collect data for up to 60 days for a country, from the day it reaches $th$ percent of peak daily cases in that country. However, the model estimates could be biased and fit to the given set of data points. To estimate the robustness of the model, we estimate the model parameters by collecting data for up to $D$ days from the day that country reaches threshold $th$. The results for the combined effect of mask, social mobility and NPIs for different $D \in [35,45,55,65,75,85]$ is shown in Figure S39.

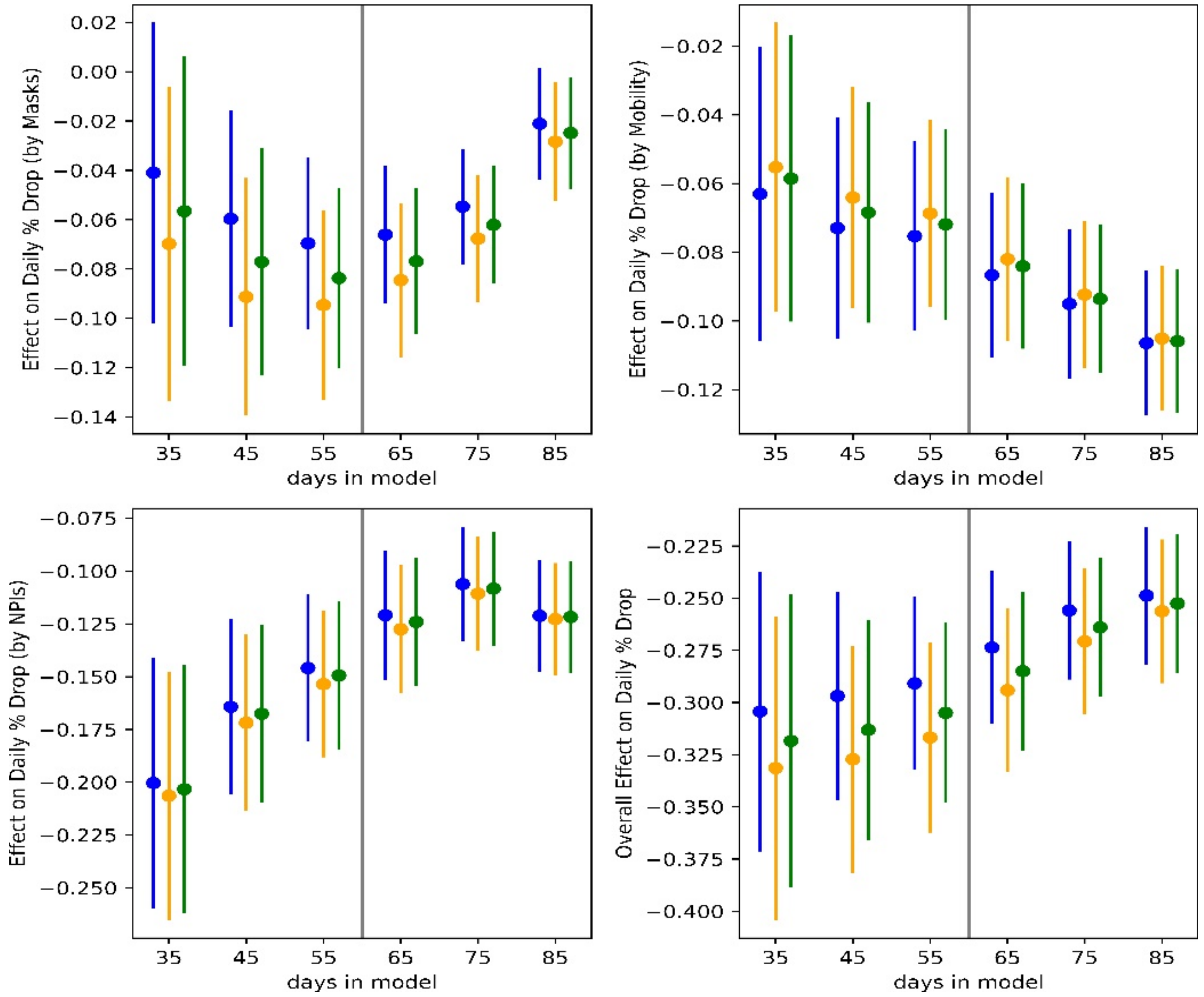


Figure S39. Combined effect of Mask, Social Mobility and NPIs when data is collected for different number of days. We consider a shift of 9 days for these results (Note that a shift of 9 days was the best fit for a model that used data for 60 days. The results show consistency within the bounds of the combined effects.

### S4.7 Interpolating Mask Survey Numbers Between Survey Days

We use survey data released by the Institute of Global Health Innovation (IGHI) at Imperial College London and YouGov4 for reported mask-wearing across multiple countries. The data present global insights on people's reported behavior in response to COVID-19. The dataset provides the percentage of population in each country who report to wear a mask in public places. Because these surveys were conducted at an interval of several days, we used linear interpolation to estimate the percentage of the population that would wear masks in public spaces for days when the data were unavailable (Figure S4). To check the robustness of estimates from the model, here we use a quadratic interpolation method to estimate the percentage of population that would wear masks in public spaces for days between surveys. The estimate for stated mask wearing using quadratic interpolation is shown in Figure S40.

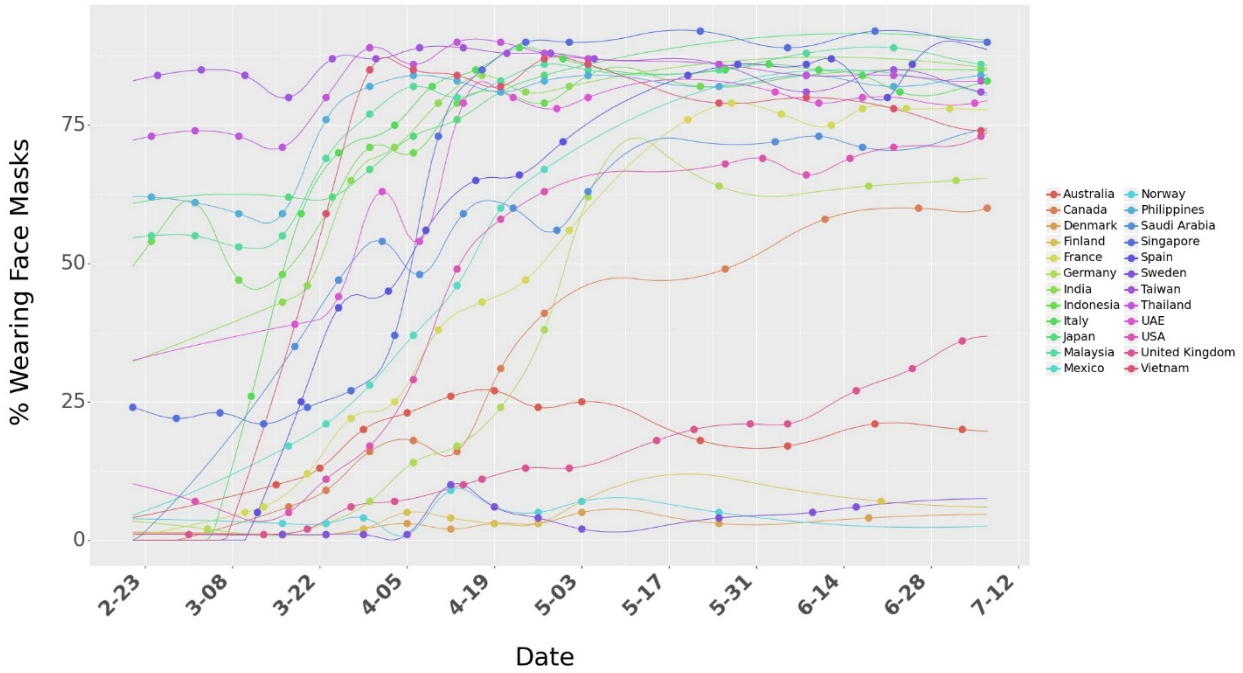

Figure S40. Survey data on percentage of people who say they wear a face mask when in public spaces. We use quadratic interpolation to consider mask numbers for days between surveys days. The dots represent the raw numbers from surveys.

The results for the association of mask, social mobility, NPIs and the combined effect of masks, social mobility and NPIs on growth rate for quadratic interpolation is shown in Figure S41. We use a $shift$ of 9 days and transformed masks as our focal model ($\ln(1 + mask)$). We also show similar for linear interpolation (focal model in this paper) to show that the parameter estimates are not appreciably different.

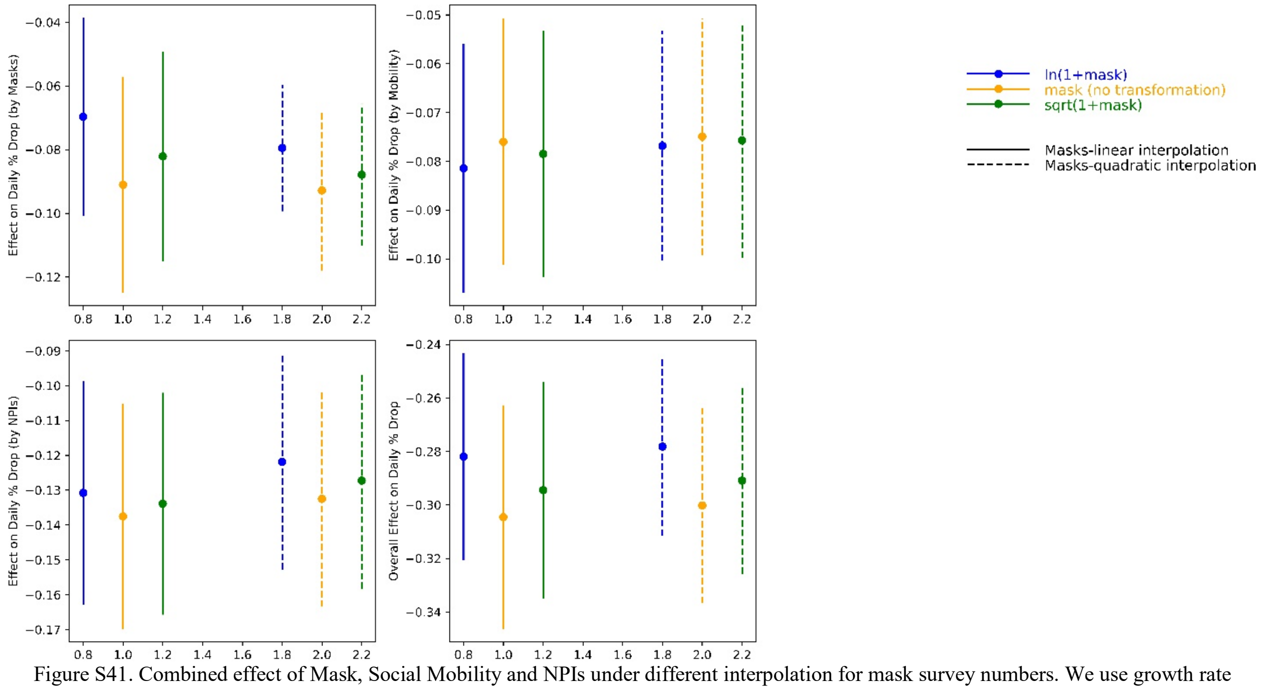

Figure S41. Combined effect of Mask, Social Mobility and NPIs under different interpolation for mask survey numbers. We use growth rate model with a $shift$ of 9 days and consider data for 60 days.

**Data and Code Availability**

All codes have been written in python 3.7 programming language. All data and codes are available at open sourced Github repository at: https://github.com/ashutoshnayakIE/COVID-masks. Data is stored in python's numpy format. However, the raw data can be procured from the sources mentioned in the references below (link for raw data set is also provided in the Github repository).
